# Diabetic retinopathy feature utilisation in 54 convolutional neural networks: a heatmap analysis

**DOI:** 10.64898/2026.06.19.26356040

**Authors:** Timothy I. Murphy, James A. Armitage

## Abstract

**Purpose:** To investigate how artificial intelligence (AI) systems detect referable diabetic retinopathy (DR) from retinal photographs by analysing heatmap patterns and determining their overlap with DR features.

**Methods:** Fifty-four AI systems were developed using 27 backbone architectures, with each implemented as both binary-referable and multi-class grading models based on the International Clinical Diabetic Retinopathy (ICDR) grading scale. Models were trained on DDR, BRSET and Kaggle datasets. After training, each model analysed 749 images with DR feature annotations, with Grad-CAM, Integrated Gradients, LIME and SHAP heatmaps generated and compared to pixel-level annotations of microaneurysms, haemorrhages, exudates, cotton wool spots, venous beading, intraretinal microvascular abnormalities and neovascularisation.

**Results:** All models achieved acceptable predictive performance (AUROC >0.8 for most architectures). Heatmap analysis revealed consistent attention to the central and temporal macular region. Exudates and cotton wool spots were highlighted most frequently by the heatmaps, with venous beading and neovascularisation at the disc showing poor overall coverage for binary referable classifiers. Models grading per the ICDR scale demonstrated higher coverage for all features. Substantial variability was observed between architectures, suggesting different feature detection capabilities. Interestingly, the heatmap analysis indicated that the models may be using different features to the ICDR grading scale definitions.

**Conclusion:** AI models do not uniformly correlate with all DR features when detecting referable DR, limiting their predictive performance in unusual presentations. Heatmap aggregation analysis provides a scalable method for analysing model behaviour, allowing strengths and weaknesses to be assessed. These findings may help improve clinician’s trust and acceptance of AI.

## Introduction

Diabetes is a significant global health concern, affecting an estimated 589 million adults aged 20-79 in 2025, approximately 11% of the population (International Diabetes Federation 2025 p. 42). This prevalence is expected to increase 17% by 2050, predominantly in lower income countries (International Diabetes Federation 2025 p. 42). Diabetes can cause a variety of systemic complications including neuropathy, nephropathy and retinopathy, leading to lower-limb amputation, kidney damage and blindness, and is a significant global cause of morbidity and mortality (Lu et al. 2023).

Diabetic retinopathy (DR) is a common complication of diabetes and a leading cause of blindness (Flaxman et al. 2017). DR refers to damage to the retinal microvasculature and can present anywhere in the retina in various forms (Figure 1), hereby referred to as DR features:

1. Microaneurysms; small and localised areas of capillary wall weakness that cause outpouching of vessels.
2. Intraretinal haemorrhages; breakages in small vessels feeding the retinal layers, resulting in bleeds.
3. Exudates; proteins and lipid accumulations in the retina that remain after leakage of plasma from weakened blood vessels.
4. Cotton wool spots; diffuse edged white lesions of the retinal nerve fibre layers caused by localised ischaemia as a result of blood vessel insufficiency.
5. Venous beading (VB); altered shape of veins as a result of vessel wall damage, resulting in the veins taking on a lumpy or sausage-like appearance.
6. Intraretinal microvascular abnormalities (IRMA); modification of existing capillary networks to shunt blood around areas of capillary blockage.
7. Neovascularisation; development of new blood vessels in response to signals from oxygen starved tissues. These vessels are fragile and leak or break easily.
8. Vitreous or pre-retinal haemorrhages; New blood vessels grow off the face of the retina and if they break, result in large volume bleeds that leak into the space between the retina and vitreous humour or into the vitreous itself.

**Figure 1:**
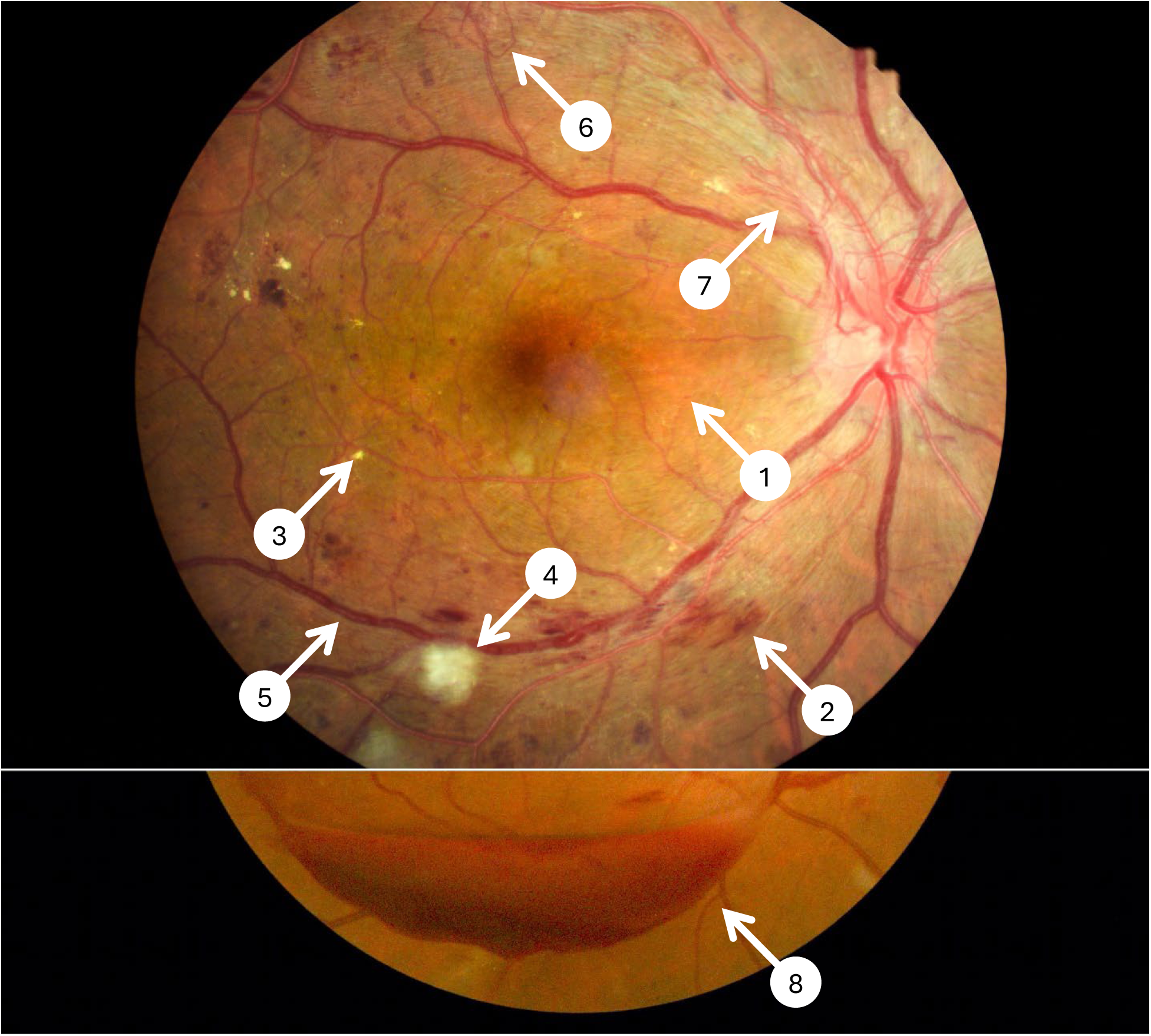
Examples of diabetic retinopathy features visible in a retinal photograph. Images adapted from Li et al. (2019). 1: microaneurysm. 2: intraretinal haemorrhage. 3: exudate. 4: cotton wool spot. 5: venous beading. 6: intraretinal microvascular abnormality. 7: neovascularisation. 8: pre-retinal haemorrhage. Contrast and colour saturation have been increased for clarity.

Retinal neovascularisation from DR, known as proliferative DR, can lead to fibrosis (scarring), retinal detachment, and irreversible blindness (Fong et al. 1999). Diabetic macular oedema can also be caused by diabetes, however this is classified separately to the primary features of DR so will not be discussed here.

Multiple grading scales exist for diabetic retinopathy, with the International Clinical Diabetic Retinopathy Disease Severity Scale (ICDR) (Wilkinson et al. 2003) commonly used in clinical practice and artificial intelligence (AI) research (Table 1). Using this grading scale, DR is assessed and graded based on the presence and number of key DR features, with each grade indicating the risk of progression to proliferative DR and risk of vision loss.

**Table 1:** International Clinical Diabetic Retinopathy Disease Severity Scale (Wilkinson et al. 2003). DR: diabetic retinopathy. NPDR: non-proliferative DR. IRMA: intraretinal microvascular abnormalities.

| **Disease Severity** | **Clinical Findings** |
| --- | --- |
| **No apparent retinopathy** | No abnormalities |
| **Mild NPDR** | Microaneurysms only |
| **Moderate NPDR** | More than just microaneurysms but less than severe NPDR |
| **Severe NPDR** | Any of the following, without signs of proliferative DR:<br>• >20 intraretinal haemorrhages in all 4 quadrants; or<br>• Venous beading in 2+ quadrants; or<br>• IRMA in 1+ quadrant |
| **Proliferative DR** | Neovascularisation or vitreous/pre-retinal haemorrhage |

This information is used clinically to determine when patients should be reviewed and/or referred to ophthalmology. Thus, misclassification of disease severity can lead to sub-optimal patient management relative to their risk of progression to sight threatening disease. Of note is that the ICDR scale makes no mention of exudates or cotton wools spots, despite them being recognised features of the disease, as they were deemed to have low risk of progression to proliferative DR (Early Treatment Diabetic Retinopathy Study Research Group 1991).

Significant work has been undertaken to develop AI-driven systems to detect DR from retinal photographs (Grauslund 2022, Grzybowski et al. 2020). Many of these systems are designed as screening or triage tools, considering Moderate NPDR or worse as referable; requiring referral to an eye specialist for assessment or treatment; or non-referable where grading is Mild NPDR or no DR (Abràmoff et al. 2013, van der Heijden et al. 2018). Literature suggests that these systems are at least comparable to human graders, and these systems are now being used in clinical practice and screening programs (Brant et al. 2025, van der Heijden et al. 2018).

While the performance of these AI systems is not disputed, it is not clear exactly how AI systems develop these predictions. Modern AI systems utilise neural networks, resulting in a “black box” system where input data is converted into disease prediction with little insight into how these data are processed (Murphy et al. 2023). This opacity is largely driven by the complexity of the system; images are converted into pixels, then further into individual colour channels, before undergoing tens or even hundreds of millions of mathematical operations. This complexity and departure from clinical explainability has created a barrier to widespread adoption of AI in the clinic, as clinicians are suspicious of disease predictions that they may not be able to explain or verify with other clinical testing (Murphy et al. 2023, Wang, Kaushal & Khullar 2020). This suspicion is well placed as regulators and national professional associations all clearly ascribe risk and responsibility for using AI with the clinician, who remains medicolegally liable in the event of misdiagnosis by an AI system (Australian Health Practitioner Regulation Agency 2024, Optometry Australia 2024). The Australian Health Practitioner Regulation Agency publishes unambiguous advice on this topic; “…Regardless of what technology is used in providing healthcare the practitioner remains responsible for delivering safe and quality care and for ensuring their own practice meets the professional obligations set out in their Code of Conduct. Practitioners must apply human judgment to any output of AI. TGA approval of a tool does not change a practitioner’s responsibility to apply human oversight and judgment to their use of AI …” (Australian Health Practitioner Regulation Agency 2024). Techniques such as occlusion-based explanations (Zeiler & Fergus 2014) and Grad-CAM (Selvaraju et al. 2017) have been developed to create heatmaps which can be overlayed on the image to estimate which features had the highest influence on the prediction, however these estimates are done per image which prevents a deeper understanding of strengths, weaknesses and biases of the AI system.

As AI has no knowledge of DR features or the specifics of the grading scale, several questions arise; do heatmaps correspond with relevant DR features used for grading? Are all relevant DR features used when an image is analysed, or are there some features which AI does not seem to utilise? Does AI place more emphasis on specific retinal areas or anatomical landmarks? By analysing heatmap outputs from multiple AI systems, this study aims to address these questions for AI that predicts referable diabetic retinopathy from retinal photos, with a goal of developing interpretable AI outputs that can provide clinicians with the data and assurances they need to comply with regulatory guidelines governing the use of AI tools.

## Materials and methods

This study created various neural networks, using existing backbones, to grade retinal photographs as referable or not referable DR. In keeping with previous work (Abràmoff et al. 2013, Murphy et al. 2025, van der Heijden et al. 2018), referable DR was defined as moderate non-proliferative diabetic retinopathy (NPDR) or worse on the ICDR grading scale (Wilkinson et al. 2003).

### Models

To maximise the generalisability of the findings, models were created from 27 publicly-available Keras backbones (Table 2) using TensorFlow. Models were created using two different techniques:

1. Predict whether an image shows referable disease (“refer” models); or
2. Grade images based on the ICDR grading scale and consider predictions of moderate NPDR or worse as referable (“grade” models).

**Table 2:** Neural network backbones used to create diabetic retinopathy prediction models.

| Model | Authors | Input shape | Parameters (millions) |
| --- | --- | --- | --- |
| **ConvNeXtTiny** | Liu et al. (2022) | 224×224 | 28.6 |
| **ConvNeXtSmall** | Liu et al. (2022) | 224×224 | 50.2 |
| **ConvNeXtBase** | Liu et al. (2022) | 224×224 | 88.5 |
| **ConvNeXtLarge** | Liu et al. (2022) | 224×224 | 197.7 |
| **ConvNeXtXLarge** | Liu et al. (2022) | 224×224 | 350.1 |
| **DenseNet121** | Huang et al. (2017) | 224×224 | 8.1 |
| **DenseNet169** | Huang et al. (2017) | 224×224 | 14.3 |
| **DenseNet201** | Huang et al. (2017) | 224×224 | 20.2 |
| **EfficientNetV2B0** | Tan & Le (2021) | 224×224 | 7.2 |
| **EfficientNetV2B1** | Tan & Le (2021) | 240×240 | 8.2 |
| **EfficientNetV2B2** | Tan & Le (2021) | 260×260 | 10.2 |
| **EfficientNetV2B3** | Tan & Le (2021) | 300×300 | 14.5 |
| **InceptionResNetV2** | Szegedy et al. (2017) | 299×299 | 55.9 |
| **InceptionV3** | Szegedy et al. (2016) | 299×299 | 23.9 |
| **MobileNet** | Howard et al. (2017) | 224×224 | 4.3 |
| **MobileNetV2** | Sandler et al. (2018) | 224×224 | 3.5 |
| **NASNetLarge** | Zoph et al. (2018) | 331×331 | 88.9 |
| **NASNetMobile** | Zoph et al. (2018) | 224×224 | 5.3 |
| **ResNet50** | He et al. (2016) | 224×224 | 25.6 |
| **ResNet101** | He et al. (2016) | 224×224 | 44.7 |
| **ResNet152** | He et al. (2016) | 224×224 | 60.4 |
| **ResNet50V2** | He et al. (2016) | 224×224 | 25.6 |
| **ResNet101V2** | He et al. (2016) | 224×224 | 44.7 |
| **ResNet152V2** | He et al. (2016) | 224×224 | 60.4 |
| **VGG16** | Simonyan & Zisserman (2015) | 224×224 | 138.4 |
| **VGG19** | Simonyan & Zisserman (2015) | 224×224 | 143.7 |
| **Xception** | Chollet (2017) | 299×299 | 22.9 |

Each backbone was used to create both refer and grade models, resulting in 54 models in total.

### Datasets

Images were sourced from DDR (Li et al. 2019), BRSET (Nakayama et al. 2024) and Kaggle (Warner 2019) datasets, using an 80:10:10 split for training, validation and test groups, respectively. To maintain an even class balance, 39,062 images were used to create refer models (19,531 in each referable/non-referable category) and 12,580 images used to create grade models (2,516 in each ICDR category). Data augmentation was undertaken on the training set, with each image undergoing three or more of the following techniques:

- Horizontal and/or vertical flip
- Contrast modulation
- Gaussian noise
- Rotation
- Zoom

Augmentation increased training sets 8-fold, resulting in 250,048 images for refer models and 80,512 for grade models. No augmentation was done for validation or test sets.

For heatmap analysis, a subset of the DDR dataset was used (Li et al. 2019), combining pixel-level annotations for microaneurysms, intraretinal haemorrhages, exudates and cotton wool spots included in the dataset with additional venous beading, IRMA and neovascularisation annotations from Murphy et al. (2024). Images have also been annotated with optic disc and fovea coordinates (Murphy et al. 2024). An additional 100 images from the DDR dataset were included from people diagnosed with diabetes but without apparent retinopathy. This “heatmap set” of 849 images had the following disease severity distribution:

- No apparent retinopathy: 100
- Mild NPDR: 79
- Moderate NPDR: 548
- Severe NPDR: 41
- Proliferative DR: 81

### Training

All models were trained on an Intel Core i9 14900F CPU (Intel Corporation, California, USA) workstation with 192GB of DDR5-5600 RAM (Corsair Gaming, California, USA), an NVIDIA GeForce RTX 4070 SUPER GPU with 12GB DDR6 VRAM and 7168 CUDA cores (NVIDIA, California, USA) and an NVIDIA GeForce GTX 1650 GPU with 4GB DDR5 VRAM and 896 CUDA cores (NVIDIA, California, USA), running Debian 13. All software ran within a CUDA-enabled Docker container (Merkel 2014) based on Ubuntu 22.04. Software was written in Python 3.11 with Tensorflow 2.19.0 (TensorFlow Developers 2021) and Keras 3.11.3 (Chollet & others 2015) to create, train and evaluate models. All software used in this study is available at https://github.com/tim-murphy/dr_cnn_heatmap_analysis.

Each backbone was paired with a Global Average Pooling and Dense Layer head to provide a binary or multiclass prediction for refer and grade models, respectively. A preprocessing layer was used to resize images to the backbone’s native input shape. All models used the Adam optimiser with Binary (refer) or Sparse Categorical (grade) Cross Entropy loss functions. An overview of the model architectures is shown in Figure 2.

**Figure 2:**
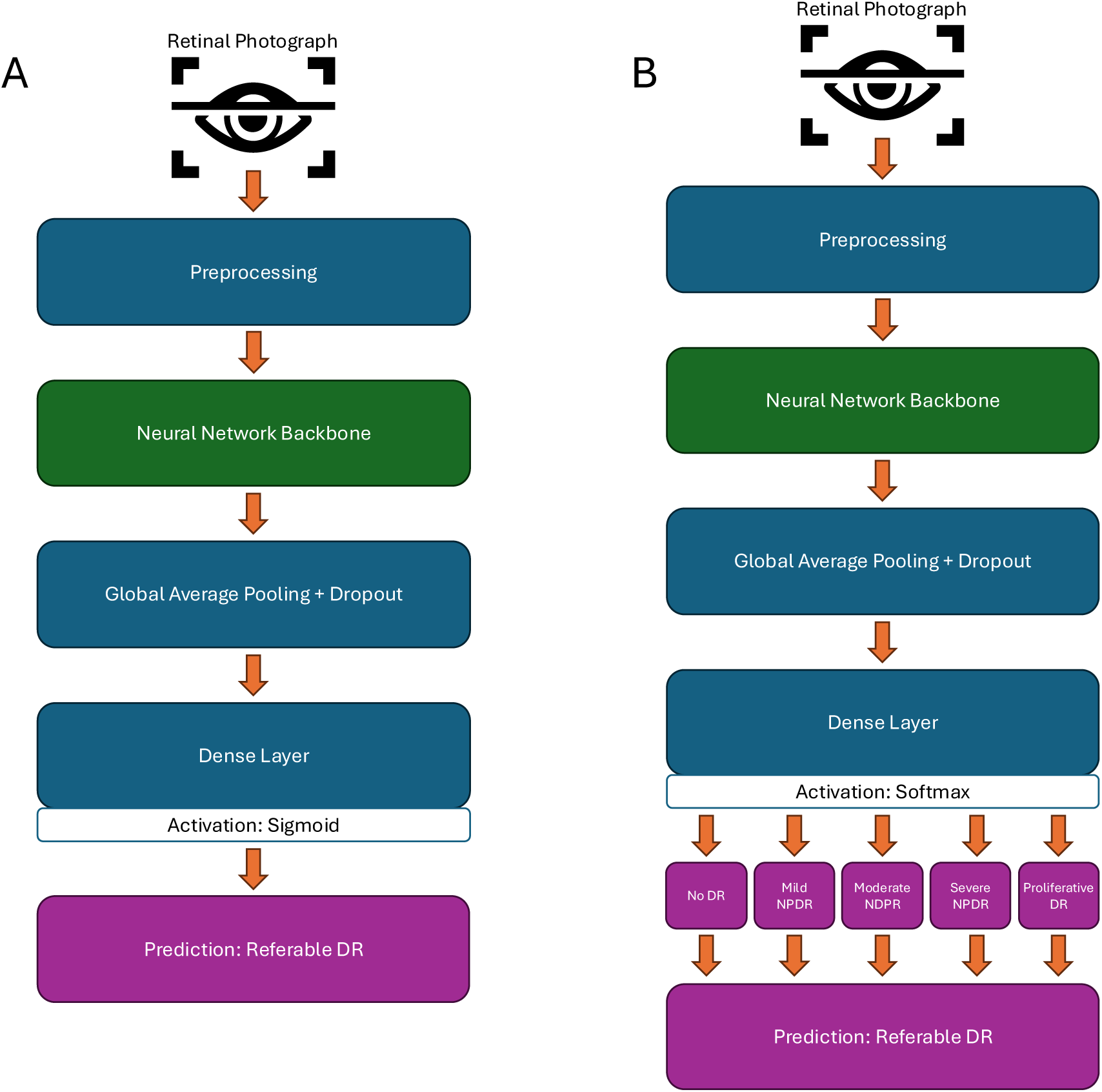
Model architecture for refer (A) and grade (B) models. DR: diabetic retinopathy. NPDR: non-proliferative DR.

Models were pretrained with default ImageNet weights available from Keras. The learning rate was set to 5×10^−5^ with a dropout rate of 0.1. A batch size of 8 was used for all models, except ConvNeXtXLarge which used a batch size of 4 due to VRAM constraints. Models were trained for a maximum of 500 epochs, with early stopping triggered after the validation loss remained above the global minimum for ten consecutive epochs (patience), reverting to the global minimum state after stopping. Additional fine tuning was then performed on the top 10% of layers, with a patience of two epochs, a learning rate of 5×10^−6^, and otherwise the same parameters as above.

### Analysis

Model performance was characterised by sensitivity (true positive rate), specificity (true negative rate), macro F1 score, and Area Under the Receiver Operating Characteristic curve (AUROC). Sensitivity and specificity values provide clinically relevant metrics, whereas F1 score and AUROC provide overall model performance metrics to allow comparison with existing models.

To illustrate the retinal locations contributing the most to a referable prediction, images from the heatmap set were assessed with each model. Grad-CAM (Selvaraju et al. 2017), Integrated Gradients (Sundararajan, Taly & Yan 2017), SHAP (Lundberg & Lee 2017) and LIME (Ribeiro, Singh & Guestrin 2016) heatmaps were generated for each image, with Otsu thresholding applied to each remove noise (Liu, Himel & Wang 2024, Otsu 1979). Heatmaps were then scaled, rotated and translated to align optic discs and foveae, as per Murphy et al. (2024). Heatmaps from the left eye were flipped horizontally to allow alignment and direct comparison with the right eye. For each heatmap type, aggregate heatmaps were generated for each model by summing the normalised heatmaps from each image to highlight retinal locations commonly utilised by that model. Aggregate model heatmaps were then combined to show common hotspot and cold spot regions across all models for that heatmap type.

Model heatmaps were then compared to diabetic retinopathy feature annotations from work by Li et al. (2019) and Murphy et al. (2024). For each feature in each image deemed to contain referable DR, the fraction of DR feature pixel annotations overlapping with non-zero heatmap values was calculated. The median value for each feature annotation was then used to estimate the overall overlap for that feature for each model, with the median of these values used as the overlap across all models.

To evaluate uncertainty of the aggregate heatmaps, bootstrapping was conducted for each model over 10,000 iterations. The mean normalised pixel intensity at each pixel location was extracted from each model and the standard deviation of these values used to determine the variability at each location. Heatmaps were generated from these data to visualise areas of higher variability.

## Results

All models reached the early stopping criteria before the 500 epoch limit. Heatmaps for individual models are provided in the supplementary material.

### Refer models

Performance metrics for the models are shown in Table 3. ConvNeXt, EfficientNetV2 and VGG models produced the highest AUROC, sensitivity and F1 scores, while no backbone family produced consistently higher specificity. All models achieved an AUROC of 0.8 or above, with the exception of MobileNet and MobileNetV2. Specificity was higher than sensitivity for all models.

**Table 3:**
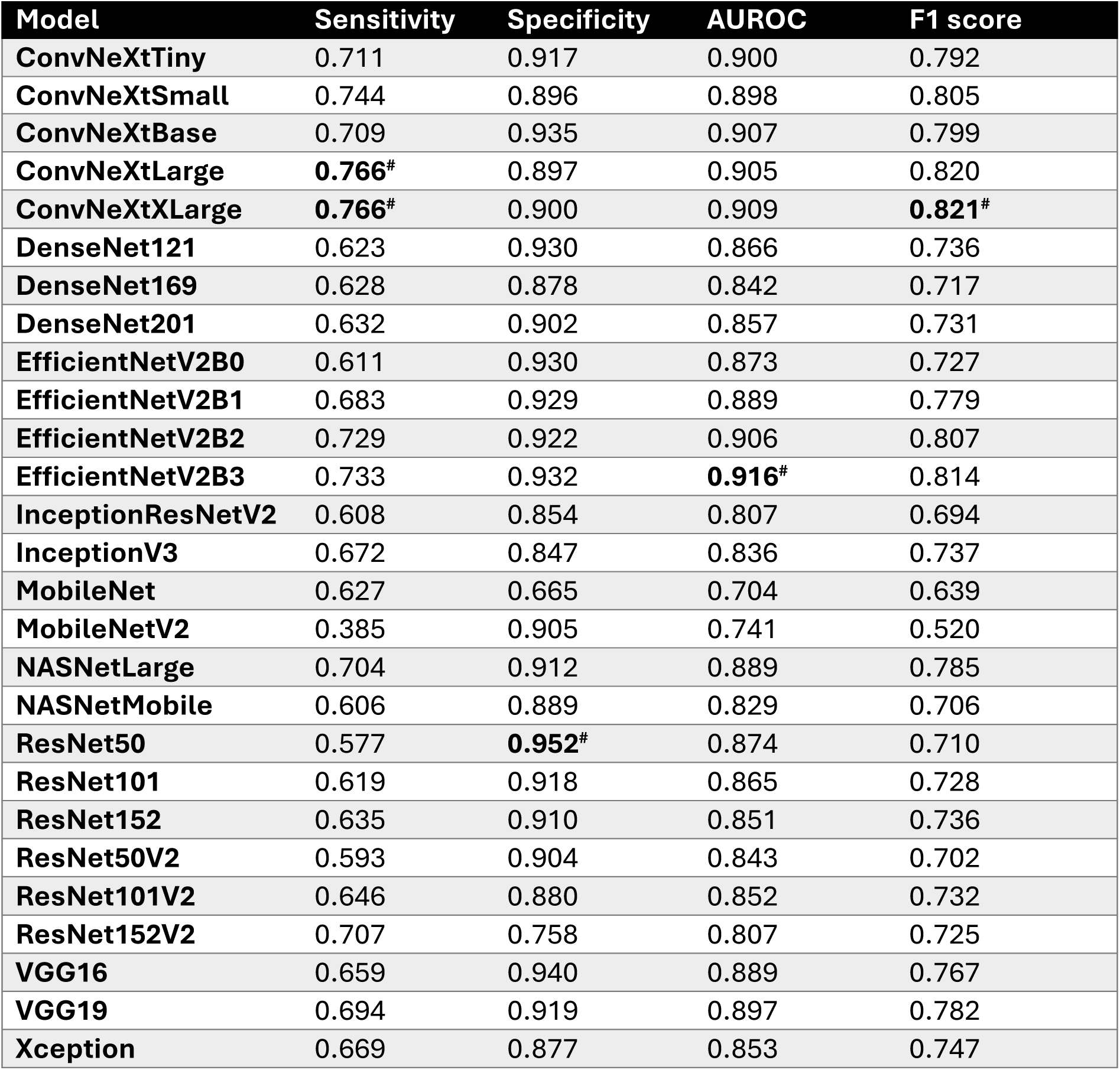
Performance of “refer” models when predicting referable diabetic retinopathy. Hash symbol (#) indicates the best performing model for a given statistic.

| Model | Sensitivity | Specificity | AUROC | F1 score |
| --- | --- | --- | --- | --- |
| ConvNeXtTiny | 0.711 | 0.917 | 0.900 | 0.792 |
| ConvNeXtSmall | 0.744 | 0.896 | 0.898 | 0.805 |
| ConvNeXtBase | 0.709 | 0.935 | 0.907 | 0.799 |
| ConvNeXtLarge | **0.766<sup>#</sup>** | 0.897 | 0.905 | 0.820 |
| ConvNeXtXLarge | **0.766<sup>#</sup>** | 0.900 | 0.909 | **0.821<sup>#</sup>** |
| DenseNet121 | 0.623 | 0.930 | 0.866 | 0.736 |
| DenseNet169 | 0.628 | 0.878 | 0.842 | 0.717 |
| DenseNet201 | 0.632 | 0.902 | 0.857 | 0.731 |
| EfficientNetV2B0 | 0.611 | 0.930 | 0.873 | 0.727 |
| EfficientNetV2B1 | 0.683 | 0.929 | 0.889 | 0.779 |
| EfficientNetV2B2 | 0.729 | 0.922 | 0.906 | 0.807 |
| EfficientNetV2B3 | 0.733 | 0.932 | **0.916<sup>#</sup>** | 0.814 |
| InceptionResNetV2 | 0.608 | 0.854 | 0.807 | 0.694 |
| InceptionV3 | 0.672 | 0.847 | 0.836 | 0.737 |
| MobileNet | 0.627 | 0.665 | 0.704 | 0.639 |
| MobileNetV2 | 0.385 | 0.905 | 0.741 | 0.520 |
| NASNetLarge | 0.704 | 0.912 | 0.889 | 0.785 |
| NASNetMobile | 0.606 | 0.889 | 0.829 | 0.706 |
| ResNet50 | 0.577 | **0.952<sup>#</sup>** | 0.874 | 0.710 |
| ResNet101 | 0.619 | 0.918 | 0.865 | 0.728 |
| ResNet152 | 0.635 | 0.910 | 0.851 | 0.736 |
| ResNet50V2 | 0.593 | 0.904 | 0.843 | 0.702 |
| ResNet101V2 | 0.646 | 0.880 | 0.852 | 0.732 |
| ResNet152V2 | 0.707 | 0.758 | 0.807 | 0.725 |
| VGG16 | 0.659 | 0.940 | 0.889 | 0.767 |
| VGG19 | 0.694 | 0.919 | 0.897 | 0.782 |
| Xception | 0.669 | 0.877 | 0.853 | 0.747 |

Aggregate heatmaps for each heatmap type are shown in Figure 3. All four heatmap types show a hotspot at the fovea, which extends temporally for Grad-CAM, SHAP and LIME. Integrated Gradients show a second hotspot at the optic disc, whereas SHAP and LIME show cold spots in this area. Integrated Gradients and LIME heatmaps are circular which matches the circular border present in retinal photographs (see Figure 1). No clear patterns emerged from images predicted as not containing referable diabetic retinopathy, nor for models with higher performance metrics. These data have been included in the supplementary material for interested readers.

**Figure 3:**
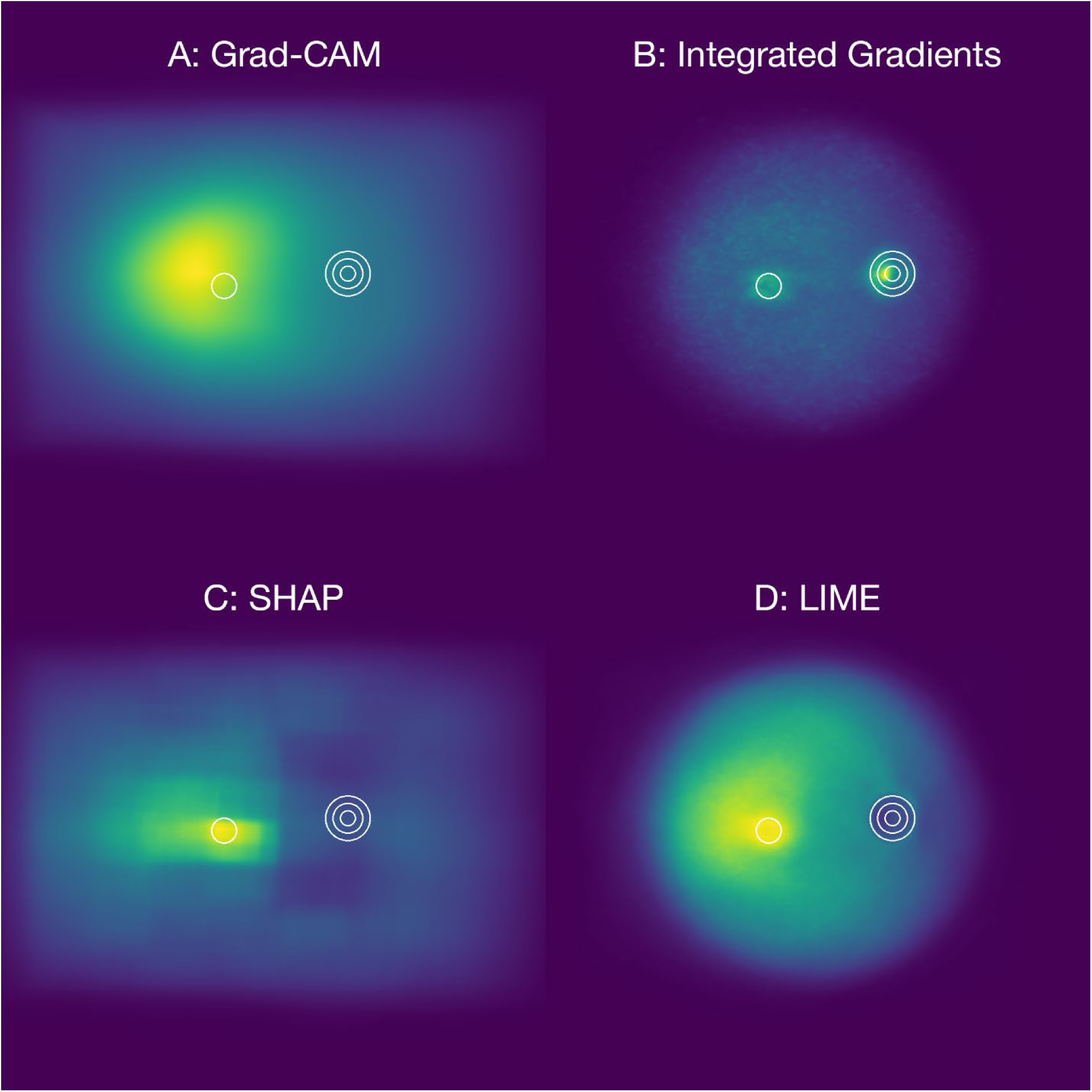
Aggregated “refer” model heatmaps for images predicted as having referable diabetic retinopathy using Grad-CAM (A), Integrated Gradients (B), SHAP (C) and LIME (D). Concentric circles indicate the location of the optic disc, with a single circle indicating the location of the fovea.

The proportion of features covered by heatmaps is shown in Figure 4. All heatmap types show exudates, cotton wool spots, intraretinal haemorrhages and microaneurysms as having higher coverage. Coverage of venous beading and IRMA, which are strong indicators of retinal ischaemia and progression risk, and neovascularisation at the disc and elsewhere, which are diagnostic for proliferative DR, was lower. For all heatmap types except Grad-CAM, coverage of venous beading and neovascularisation at the disc was close to zero.

**Figure 4:**
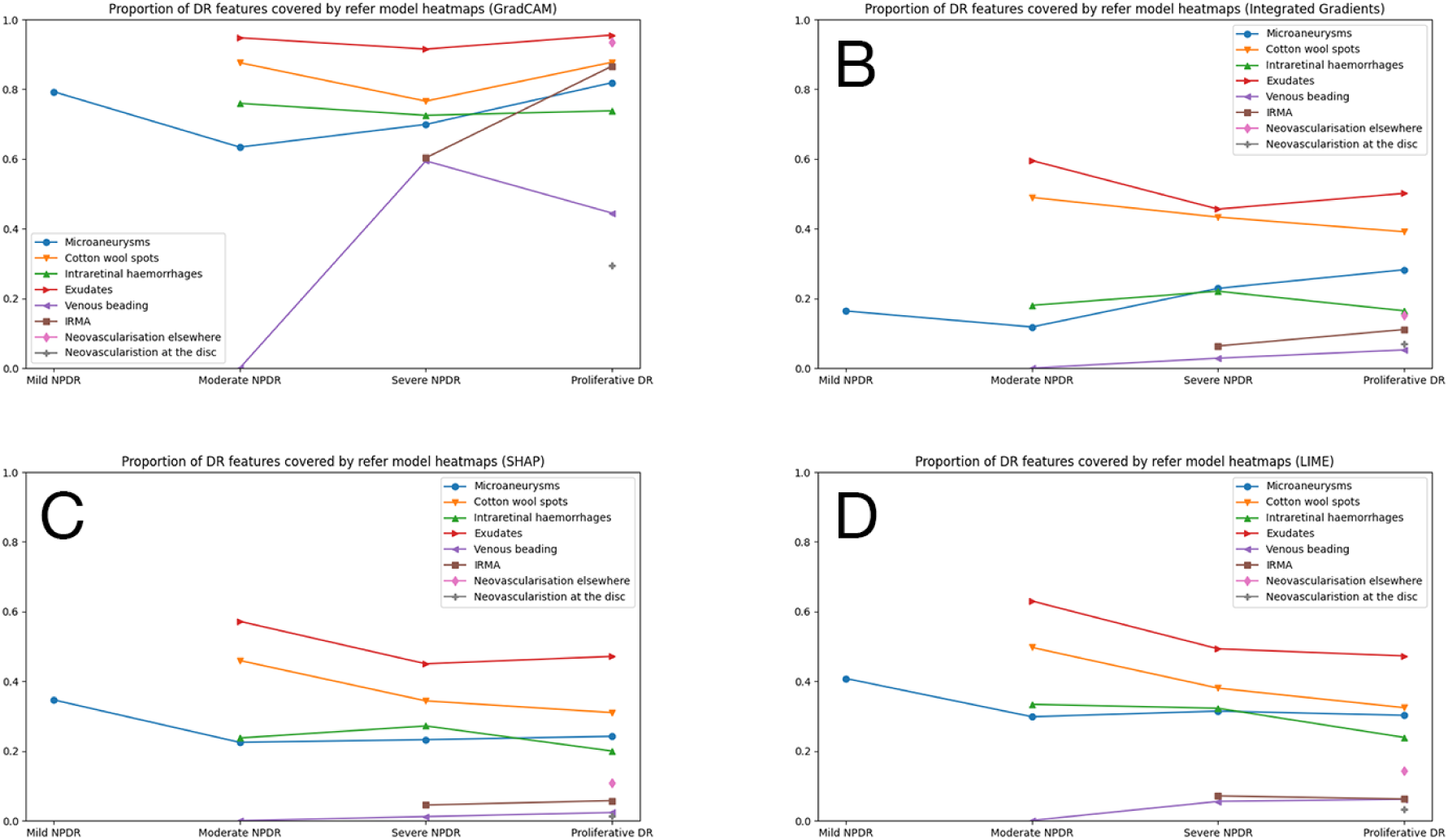
Median proportion of diabetic retinopathy features covered by Grad-CAM (A), Integrated Gradients (B), SHAP (C) and LIME (D) heatmaps, aggregated across all “refer” models.

Looking deeper into the poor coverage of neovascularisation at the disc, using Grad-CAM as an example, all models had a range of 0.00 to 1.00 except NASNetLarge which had a range of 0.45 to 1.00 (Figure 5). While the majority of models (67%) had a median coverage of less than 0.5, five of the 27 models (19%) had a median coverage of 1.00 including all MobileNet and NASNet models. This large range of coverage was reflected in most DR features across disease severities. MobileNet and NASNet backbones had very small interquartile ranges clustered around 1.0, while VGG backbones had interquartile ranges clustered around zero.

**Figure 5:**
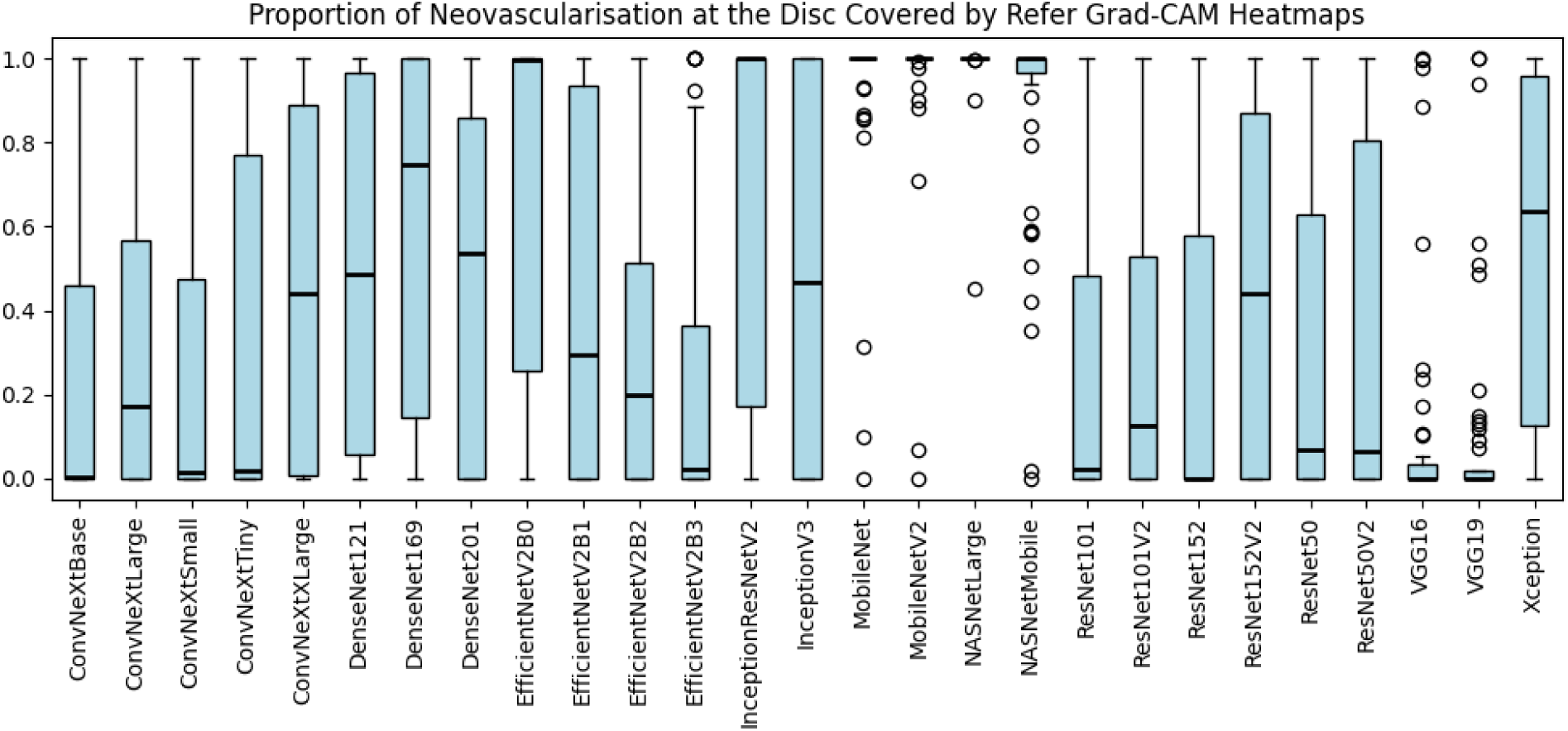
Median proportion of neovascularisation at the disc covered by “refer” model Grad-CAM heatmaps.

### Grade models

Performance metrics are shown in Table 4. ConvNeXt, EfficientNetV2 and VGG models produced the highest AUROC, sensitivity and F1 scores, while EfficientNetV2 models outperformed others for specificity. All models except MobileNetV2 achieved AUROC values above 0.8.

**Table 4:** Performance of “grade” models when predicting referable diabetic retinopathy. Hash symbol (#) indicates the best performing model for a given statistic.

| Model | Sensitivity | Specificity | AUROC | F1 score |
| --- | --- | --- | --- | --- |
| ConvNeXtTiny | 0.851 | 0.861 | 0.926 | 0.855 |
| ConvNeXtSmall | **0.884<sup>#</sup>** | 0.813 | 0.931 | 0.854 |
| ConvNeXtBase | 0.836 | 0.877 | 0.932 | 0.853 |
| ConvNeXtLarge | 0.880 | 0.786 | 0.934 | 0.841 |
| ConvNeXtXLarge | 0.824 | 0.897 | **0.938<sup>#</sup>** | 0.855 |
| DenseNet121 | 0.690 | **0.925<sup>#</sup>** | 0.889 | 0.782 |
| DenseNet169 | 0.769 | 0.883 | 0.906 | 0.815 |
| DenseNet201 | 0.778 | 0.857 | 0.894 | 0.810 |
| EfficientNetV2B0 | 0.766 | 0.899 | 0.903 | 0.820 |
| EfficientNetV2B1 | 0.771 | 0.903 | 0.912 | 0.825 |
| EfficientNetV2B2 | 0.811 | 0.891 | 0.918 | 0.854 |
| EfficientNetV2B3 | 0.819 | 0.913 | 0.932 | **0.859<sup>#</sup>** |
| InceptionResNetV2 | 0.804 | 0.800 | 0.888 | 0.802 |
| InceptionV3 | 0.803 | 0.786 | 0.876 | 0.796 |
| MobileNet | 0.534 | 0.883 | 0.811 | 0.647 |
| MobileNetV2 | 0.452 | 0.849 | 0.727 | 0.564 |
| NASNetLarge | 0.815 | 0.825 | 0.900 | 0.819 |
| NASNetMobile | 0.717 | 0.883 | 0.880 | 0.782 |
| ResNet50 | 0.731 | 0.907 | 0.900 | 0.802 |
| ResNet101 | 0.743 | 0.913 | 0.904 | 0.812 |
| ResNet152 | 0.804 | 0.829 | 0.901 | 0.814 |
| ResNet50V2 | 0.757 | 0.861 | 0.883 | 0.798 |
| ResNet101V2 | 0.799 | 0.857 | 0.896 | 0.823 |
| ResNet152V2 | 0.679 | 0.857 | 0.851 | 0.745 |
| VGG16 | 0.854 | 0.823 | 0.923 | 0.841 |
| VGG19 | 0.817 | 0.873 | 0.921 | 0.841 |
| Xception | 0.761 | 0.871 | 0.891 | 0.805 |

Aggregate heatmaps for each heatmap type are shown in Figure 6. Grad-CAM, SHAP and LIME show a hotspot around the fovea, which is larger than those seen in the “grade” heatmaps (Figure 3). Integrated Gradients and SHAP show a hotspot at the optic disc, whereas LIME shows a cold spot in this area. As with refer models, no clear patterns emerged from images predicted as not containing referable diabetic retinopathy, nor for models with higher performance metrics.

**Figure 6:**
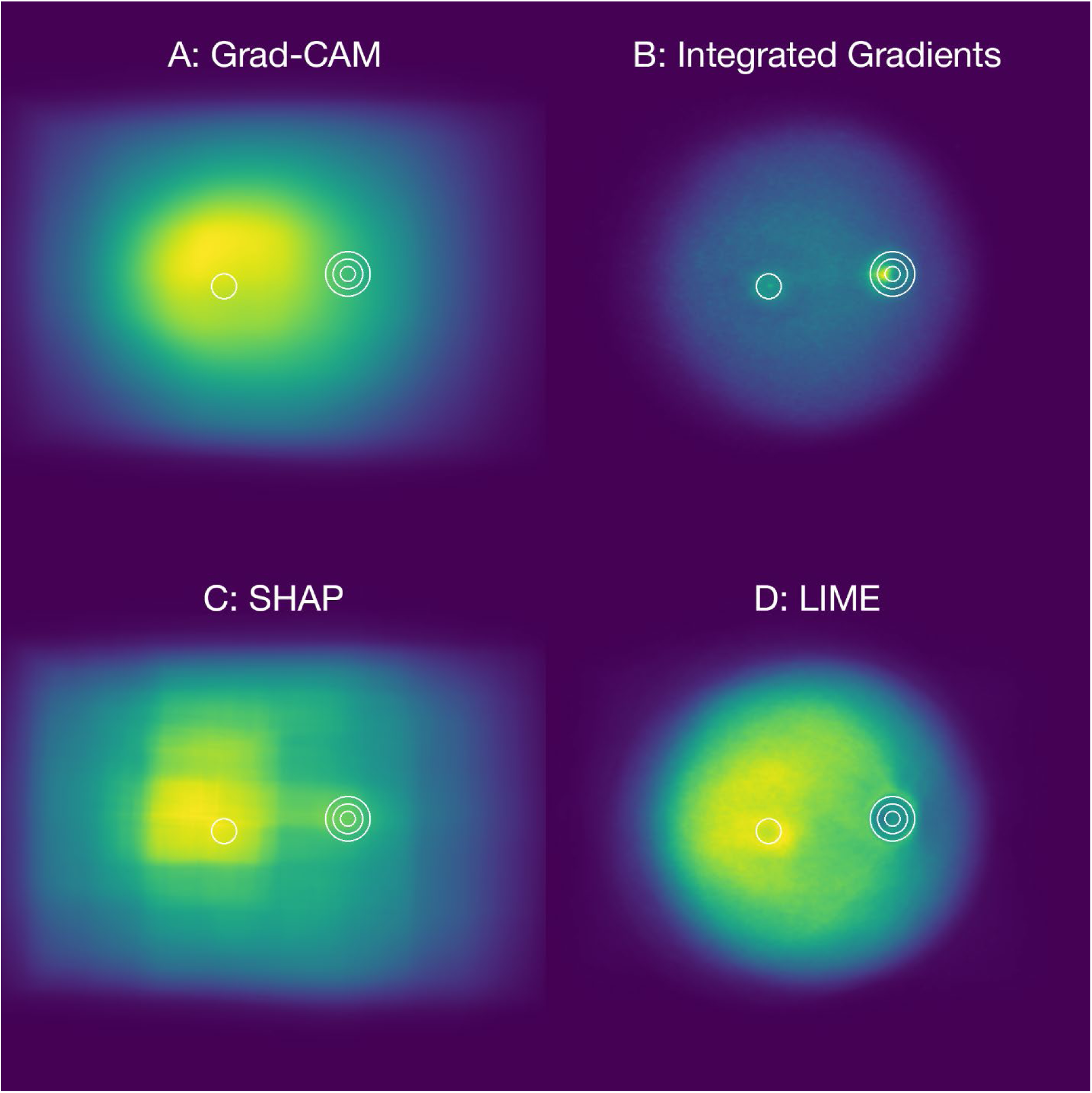
Aggregated “grade” model heatmaps for images predicted as having referable diabetic retinopathy using Grad-CAM (A), Integrated Gradients (B), SHAP (C) and LIME (D). Concentric circles indicate the location of the optic disc, with a single circle indicating the location of the fovea.

The proportion of features covered by heatmaps is shown in Figure 7. All heatmap types show exudates, cotton wool spots, intraretinal haemorrhages and microaneurysms as having higher coverage. Coverage of venous beading, IRMA and neovascularisation was higher than refer models but still lower than other features.

**Figure 7:**
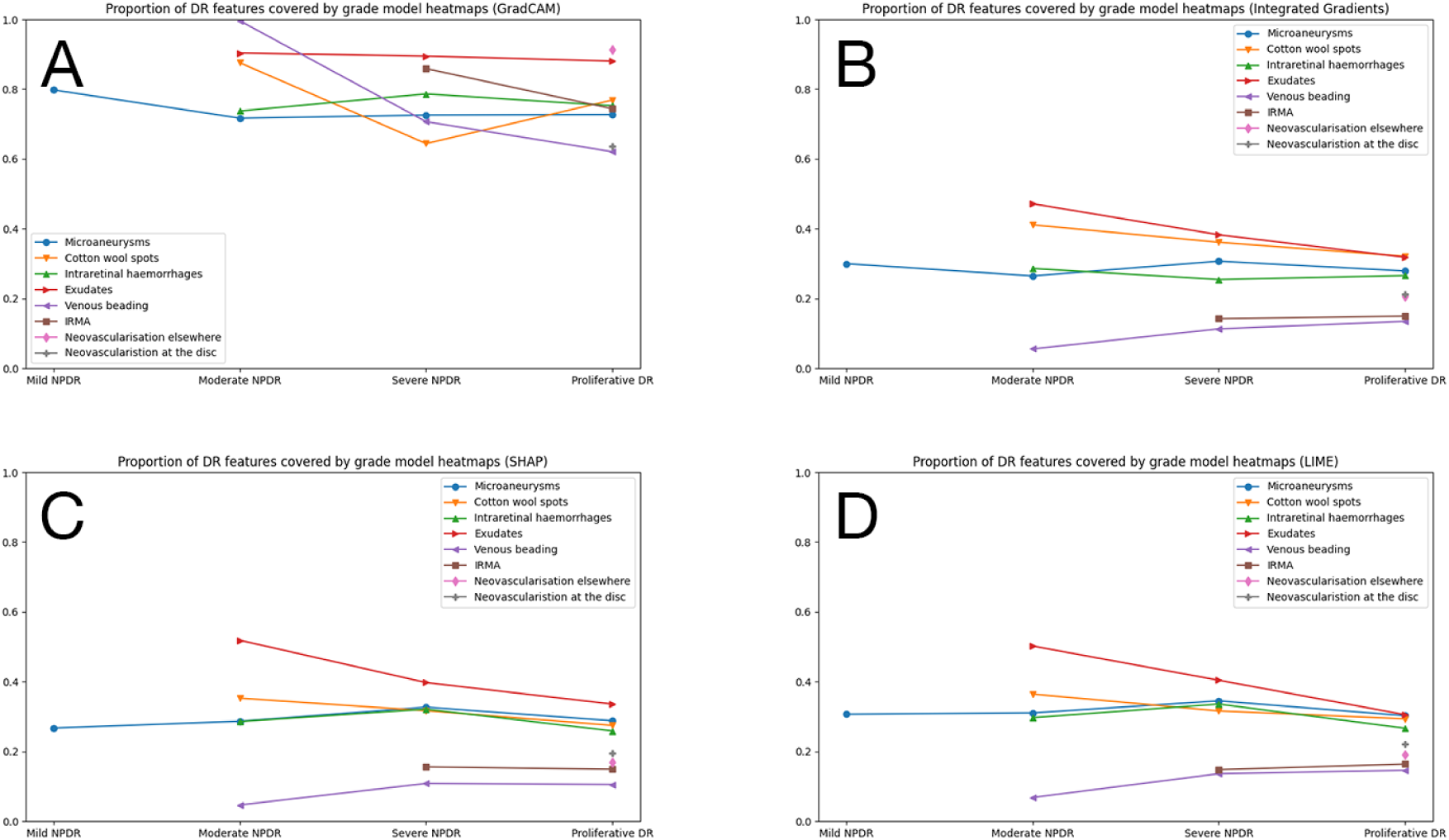
Median proportion of diabetic retinopathy features covered by Grad-CAM (A), Integrated Gradients (B), SHAP (C) and LIME (D) heatmaps, aggregated across all “grade” models.

As with refer models, neovascularisation at the disc had the lowest heatmap coverage. Using Grad-CAM again as an example, both MobileNet backbones performed well with interquartile ranges clustering around 1.0 (Figure 8). VGG backbones performed poorly with median values around 0.05, however the interquartile ranges were larger than their refer-model counterparts.

**Figure 8:**
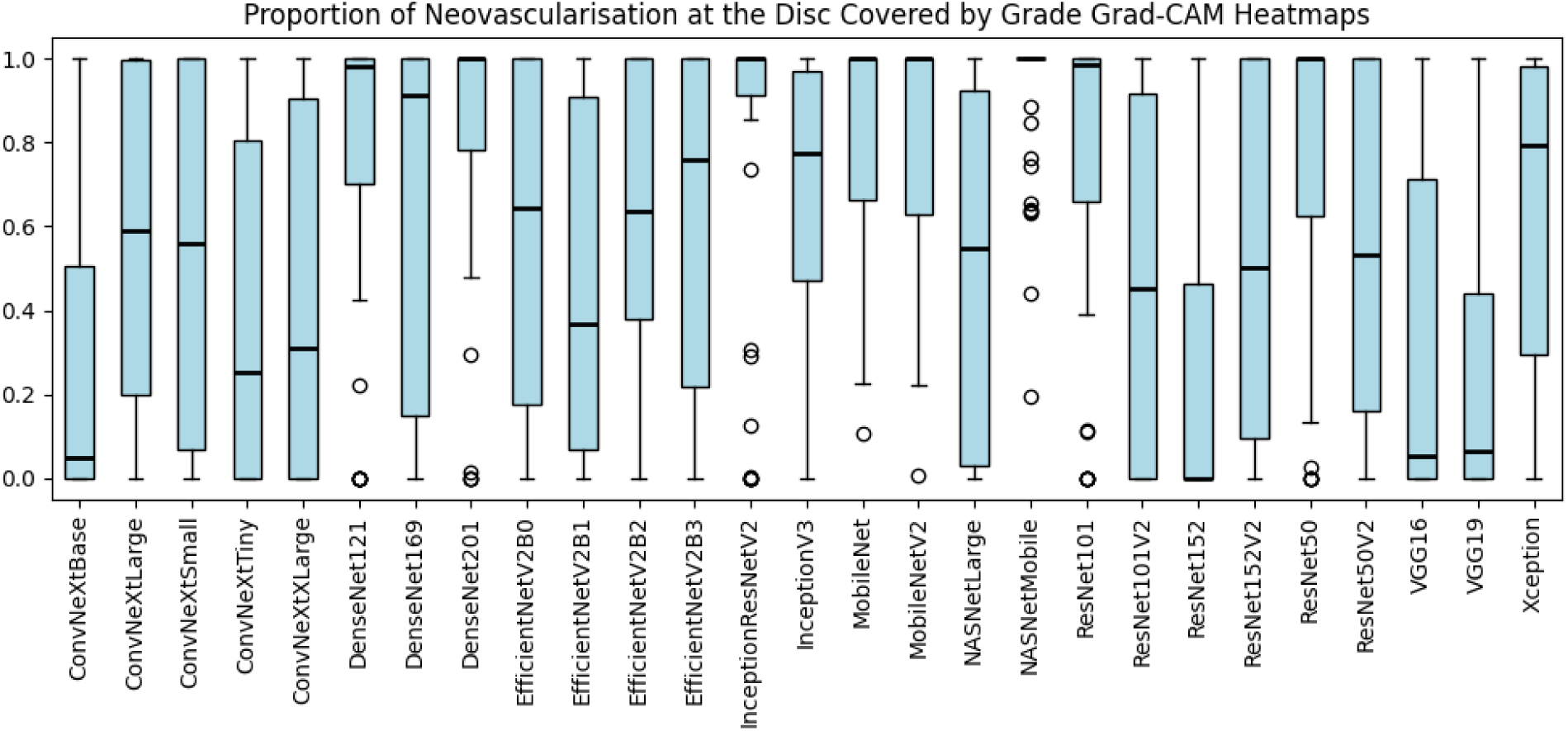
Median proportion of neovascularisation at the disc covered by “grade” model Grad-CAM heatmaps.

### Uncertainty

Bootstrapping was undertaken for all models to find the standard deviations of heatmap intensities at each pixel location, which were then visualised as heatmaps. The largest standard deviations over the image canvas are shown in Table 5. These data are represented as raw standard deviations and relative standard deviations as a percentage of the maximum heatmap value range [0, 255]. Grad-CAM heatmaps had the lowest variability, with the highest relative standard deviations being around 1%. Integrated Gradients exhibited more variability, with relative standard deviations of 7.57% for refer models and 10.32% for grade models. This overall low variability suggests the aggregated heatmaps were highly stable across resamples. When visualised as SD heatmaps these hotspots are barely perceptible, but have been included in the supplementary material for transparency.

**Table 5:** Absolute and relative maximum standard deviation for each heatmap and model type. Relative SD is the maximum standard deviation as a percentage of the full dynamic range [0, 255].

| Model type | Heatmap type | Maximum SD | Relative SD |
| --- | --- | --- | --- |
| Refer | Grad-CAM | 2.61 | 1.03% |
|  | Integrated Gradients | 19.30 | 7.57% |
|  | SHAP | 11.95 | 4.69% |
|  | LIME | 13.24 | 5.19% |
| Grade | Grad-CAM | 2.87 | 1.13% |
|  | Integrated Gradients | 26.31 | 10.32% |
|  | SHAP | 12.30 | 4.82% |
|  | LIME | 14.36 | 5.63% |

## Discussion

Understanding how a diagnostic test works is crucial to using it effectively in clinical practice (De Moraes et al. 2008). Modern AI systems predict the presence of pathology, sometimes with a heatmap to “explain” the prediction, without revealing the underlying logic behind it. This opaque processing leads to a trust deficit for clinicians, limiting uptake and widespread use (Asan, Bayrak & Choudhury 2020).

The results of this study suggest these concerns are well founded. Many AI systems used to detect referable diabetic retinopathy consider each case as a binary problem: the image contains referable disease or it does not. This study suggests AI designed and trained on this simplified grading system may rely on disease features such as exudates, which carry a high risk of vision loss only when they occur within 500µm of the fovea and are indicative of macular oedema, while neglecting features such as neovascularisation or venous beading which carry a significantly higher risk of vision loss. Many presentations of proliferative (and sight threatening) diabetic retinopathy actually present with very few microaneurysms, intraretinal haemorrhages and exudate because capillary beds have become blocked and have regressed. In cases where only neovascularisation or venous beading are visible in the image there is a risk of misclassification and vision loss. An example of such a case is shown in Figure 9, with neovascularisation clearly visible at the disc with little other retinopathy apparent. When using refer models, this image is graded incorrectly as not referable by VGG16 (confidence 73%) and VGG19 (55%), but correctly as referable by MobileNet (66%) and MobileNetV2 (77%) despite VGG backbones being superior to MobileNet in all performance metrics (Table 3). These results are predictable from the heatmap analyses of neovascularisation at the disc (NVD), with MobileNet showing higher heatmap coverage of NVD for all heatmap types compared to VGG (Figure 5). If such data were available, it may be possible for clinicians to use AI more effectively by counteracting known weakness with additional testing, potentially using a second AI system with corresponding strengths. Further research into this possibility is warranted.

**Figure 9:**
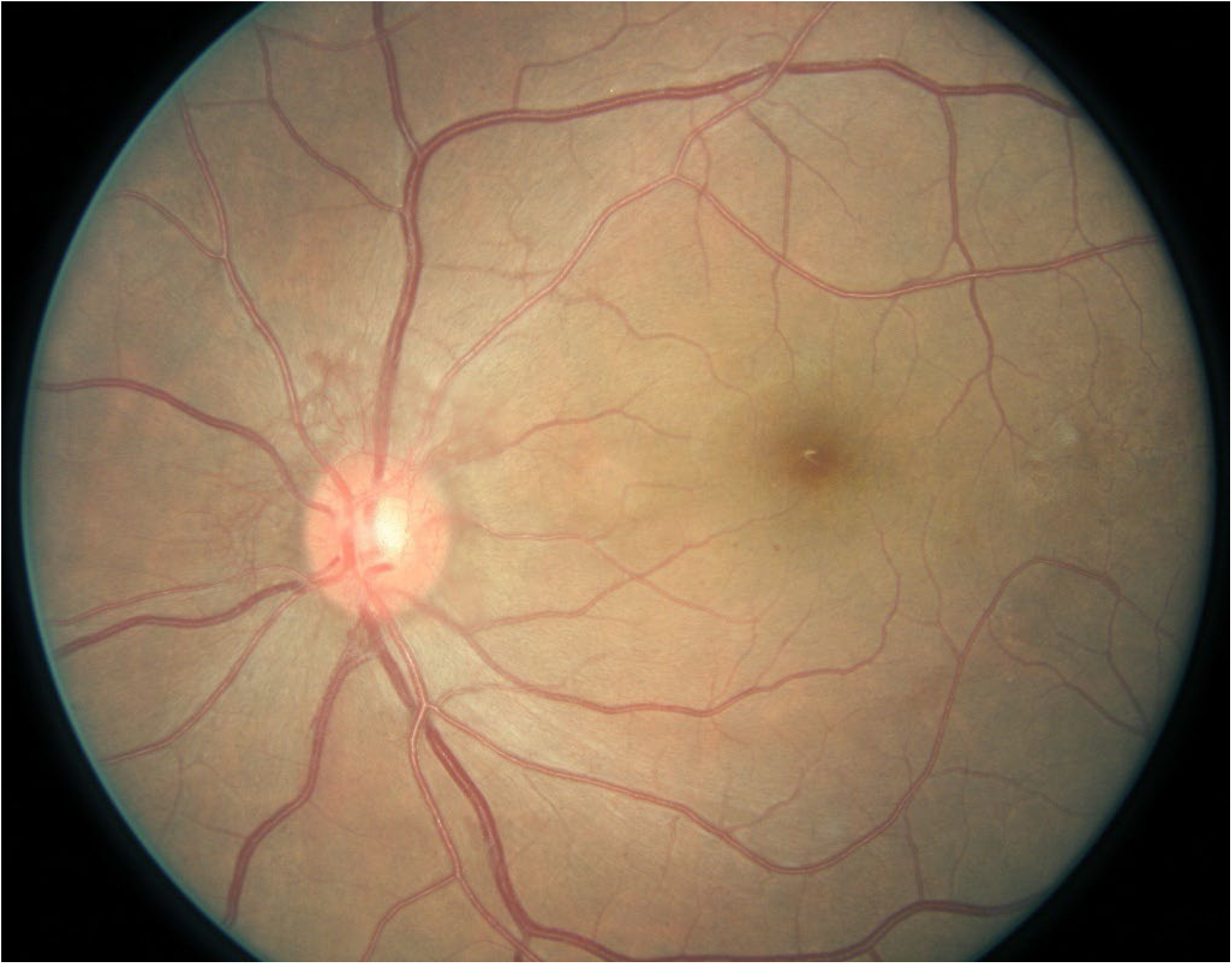
Image 10193_right.jpg from the Kaggle dataset (Warner 2019) showing proliferative diabetic retinopathy due to neovascularisation at the disc. Because capillary beds have become blocked and have regresses, there are very few microaneurysms and intraretinal haemorrhages.

Ideally, training data should be expanded to include additional examples of unusual presentations to prevent less common features from being neglected. However, such data are rare and the number of unusual presentations too high for this to be feasible. Developing models that classify disease with more granularity, and simplifying the classification from these results, may alleviate these inherent weaknesses. Results from the present study, where models first grade images against the ICDR scale and then convert to referable/non-referable, suggest this to be the case, and these models performed better overall with less training data. Indeed, for the case above (Figure 9), grade models using VGG16 and VGG19 backbones correctly flagged this image as referable (66% and 98% confidence, respectively). Different heatmap patterns were also observed for all heatmap types when the models were developed in this manner, suggesting that these varying strategies are reliant on different image features.

High correlations were observed between heatmaps and exudate and cotton wool spot locations for both training modalities. This is a surprise finding given the limited role these features play in determining disease severity according to the ICDR severity scale: the presence of either feature is diagnostic of Moderate NPDR (referable), although the number, size or morphology of these features do not otherwise influence severity. Earlier work by Klein et al. (1984) regarded these features as having a higher risk of progression to proliferative disease, however ETDRS studies (Early Treatment Diabetic Retinopathy Study Research Group 1991), which the ICDR scale is based on, downgraded this risk profile based on their larger study. It is worth noting that few of these participants showed only exudates or cotton wool spots. This raises the possibility that Severe NPDR and Proliferative DR are associated with specific exudate and/or cotton wool spot presentations, which warrants further investigation.

As the AI systems appear to be associated with disease features which are not part of the ICDR scale, risk profiles and management decisions suggested by the scale may not be appropriate when diagnosed by AI. The grading scale is used to both track disease severity and estimate the risk of disease progression and is based on the presence and number of disease features seen in the retina (Early Treatment Diabetic Retinopathy Study Research Group 1991, Wilkinson et al. 2003). While the AI may correlate an image with a severity grade, it is not clear whether additional information is being used to make this classification and, therefore, whether the risks of progression are the same. Indeed, non-vascular and functional changes have been shown to coincide or precede vascular changes in diabetic retinopathy (Rai, Maddess & Nolan 2025) which may be detectable with AI. Additional work investigating patient outcomes after AI-driven diagnoses should be undertaken to better understand this potential misalignment.

Techniques employed in this study may be used to create more efficient ensemble models. As relative strengths and weaknesses may be predictable, it may be possible to use multiple models together to maximise predictive performance and reduce uncertainty. This work could involve AI models being selectively run based on an initial screening assessment, or with an ensemble method such as linear combinations (Hashem 1997).

This study is not without limitations. A total of 54 neural networks were created and trained using the same technique, which is unlikely to be optimal. While using a specialised training regimen for each backbone may have produced higher-performing models, overall model performance was sufficient for the purposes of the study, and the consistent training technique avoided perceived bias from training one model better or worse than others. It is possible, however, that heatmaps would differ with optimal training so the results may not be widely generalisable.

While training data contained images from the USA, India, China and Brazil, the data used for diabetic retinopathy feature locations were sourced only from the DDR dataset. This dataset is sourced from a Chinese population and contains an uneven distribution of disease severity which may have skewed the results. Furthermore, this study only investigated associations between heatmap and feature locations. It is possible for a heatmap to coincide with a disease feature without that feature carrying high weighting in the prediction algorithm. There are also notable limitations with heatmap-based explanation techniques, such as features not being highlighted when multiple features influence a prediction (Chattopadhay et al. 2018, Salih et al. 2025). While 849 images were used to generate these heatmaps, it is not certain whether this was enough to account for these limitations. At the time of writing, only the DDR dataset contained necessary annotations which limited the number of images which could be used for this task. Future work using larger and more diverse datasets is recommended to confirm these findings.

Finally, all backbones used in this study were convolutional neural networks (CNNs). While vision transformers and foundation models are now commonly used in research, we decided to restrict this study to CNNs as this technology is used in most commercial systems (Wang et al. 2025). As newer technologies become more widespread for regulator-approved medical AI systems, additional research using these technologies is warranted.

Ultimately, the regulators, legal courts and consumers/patients have an expectation that the expert clinician takes responsibility for treatment and monitoring decisions. As a result, these professionals need to be satisfied that they can trust the decision support tools they use, and such trust is increased when they understand the assumptions that a tool may be relying upon when making decisions. Development of AI-based decision support tools is likely to accelerate in the foreseeable future, therefore, system architects should look to incorporate visualisation and verification features that empower the clinician to critically review model outputs. When clinicians can interrogate the logic and the image features that a model apportions high importance, they are more likely to trust output and fully utilise the decision-support system.

## Data Availability

Raw data used in this study is available from
- DDR dataset https://github.com/nkicsl/DDR-dataset
- BRSET dataset https://github.com/luisnakayama/BRSET
- Kaggle 2015 and 2019 blindness detection dataset https://www.kaggle.com/datasets/benjaminwarner/resized-2015-2019-blindness-detection-images
All software used in this study is available at https://github.com/tim-murphy/dr_cnn_heatmap_analysis.

