## Supplementary Materials for "Diabetic retinopathy feature utilisation in 54 convolutional neural networks: a heatmap analysis"

### Supplementary material

For all heatmap images, concentric circles indicate the location of the optic disc, and a single circle indicating the location of the fovea.

### 1 Refer models

#### 1.1 No referable diabetic retinopathy predicted

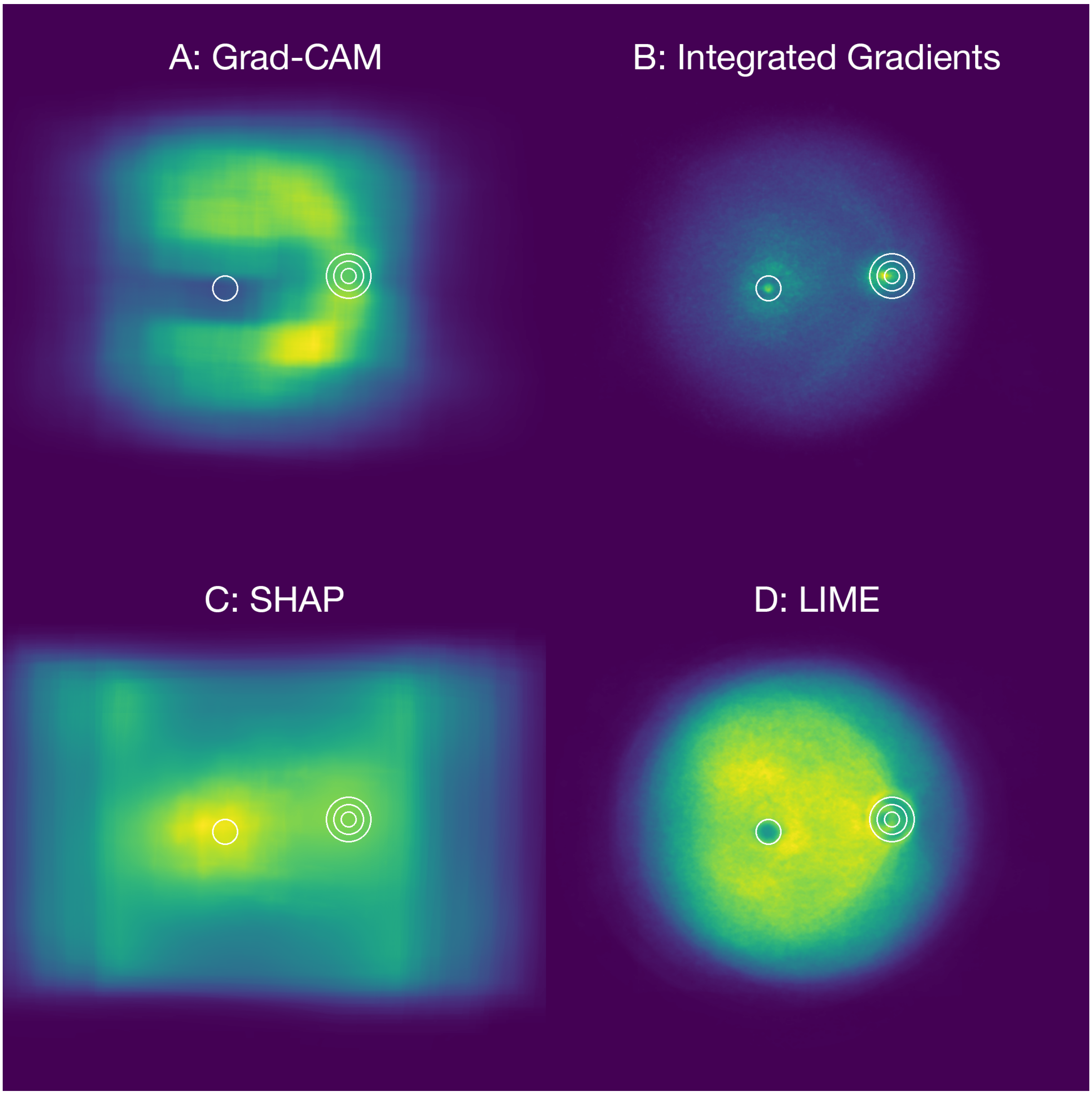

Figure 1: Aggregated “refer” model heatmaps for images predicted as not having referable diabetic retinopathy using Grad-CAM (A), Integrated Gradients (B), SHAP (C) and LIME (D).

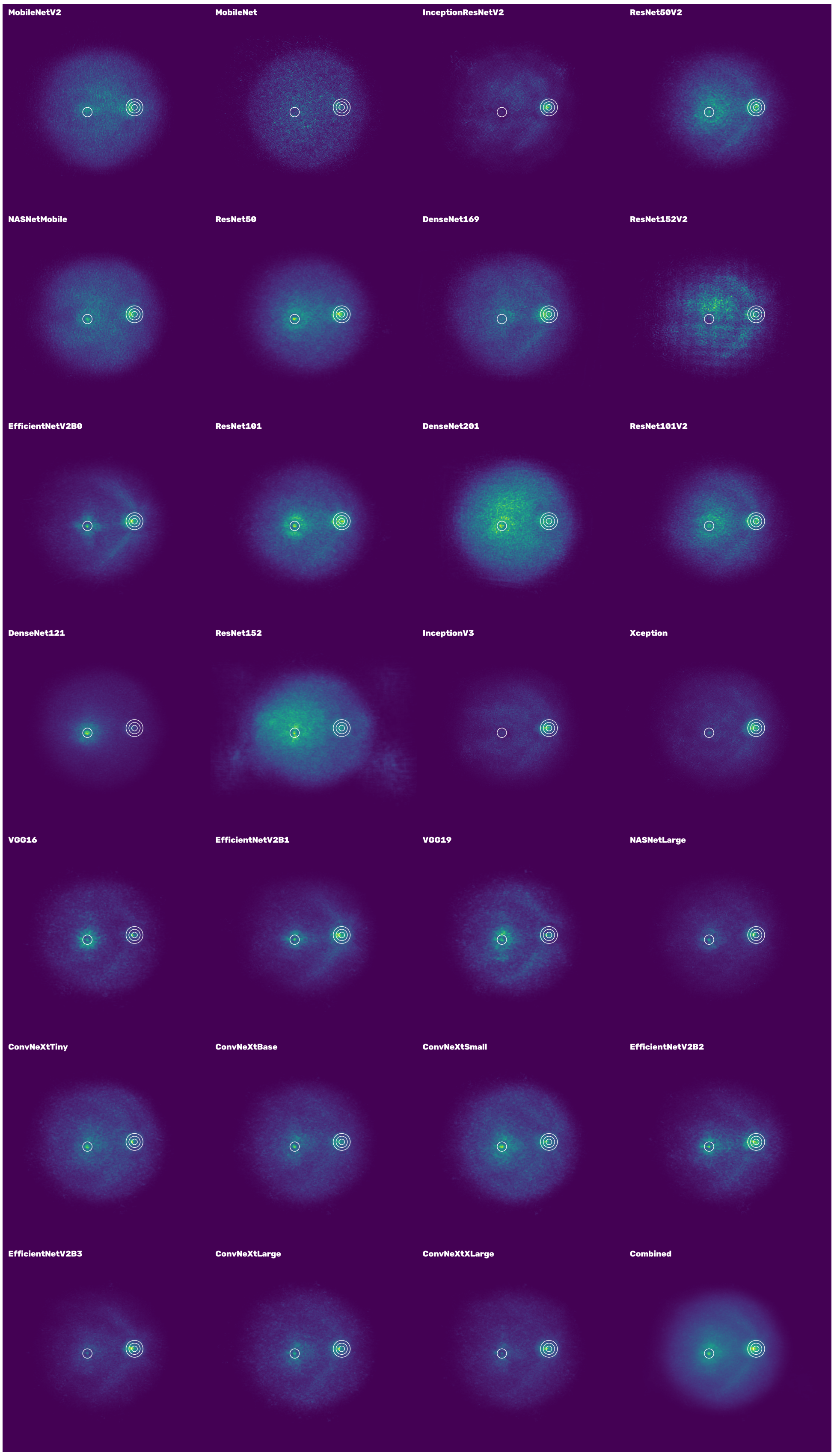

Figure 2: Collated Integrated Gradient heatmaps for "refer" models for images predicted as not having referable diabetic retinopathy. Heatmaps are ordered by Macro-F1 score in ascending order.

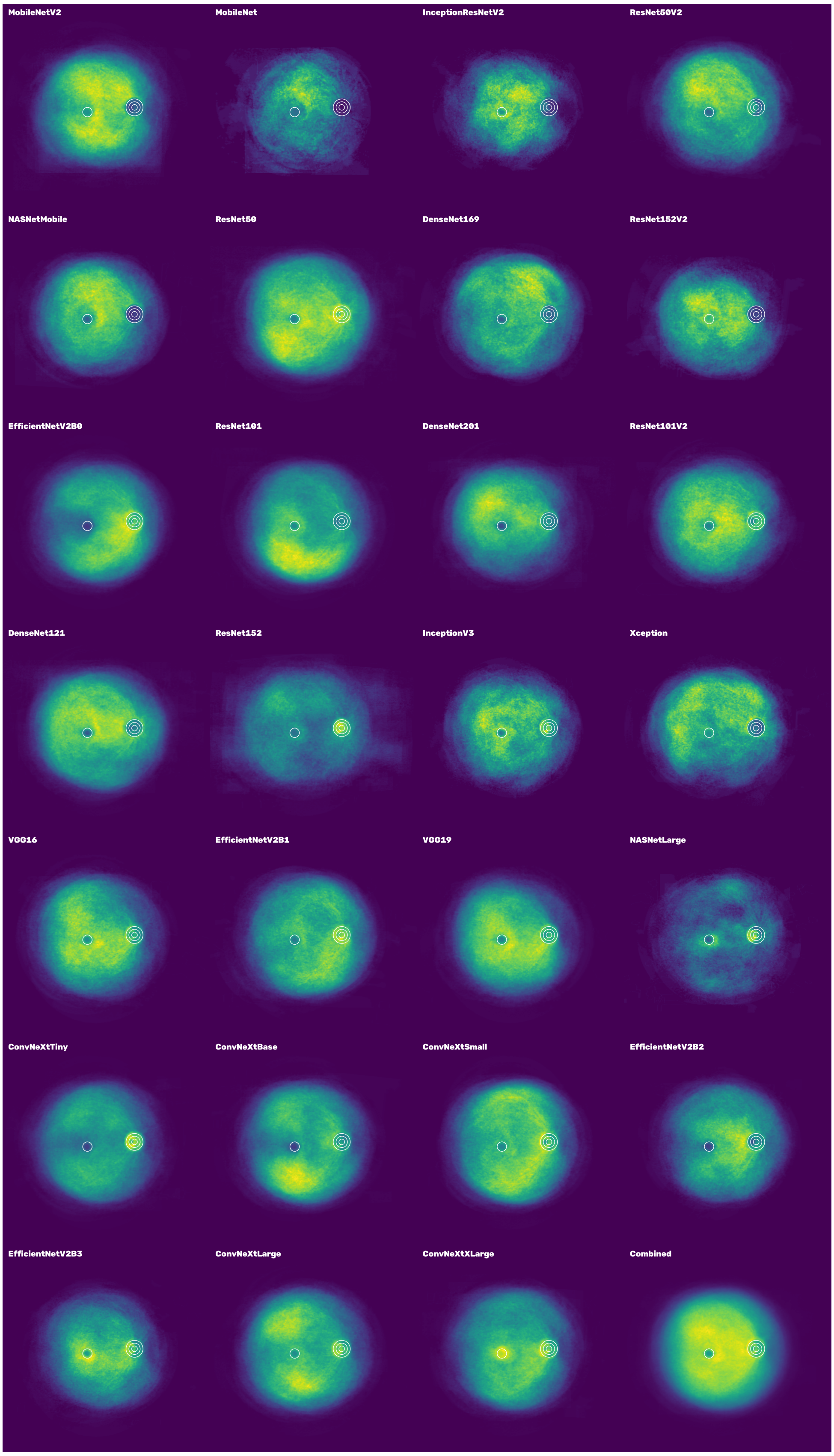

Figure 3: Collated LIME heatmaps for ”refer” models for images predicted as not having referable diabetic retinopathy. Heatmaps are ordered by Macro-F1 score in ascending order.

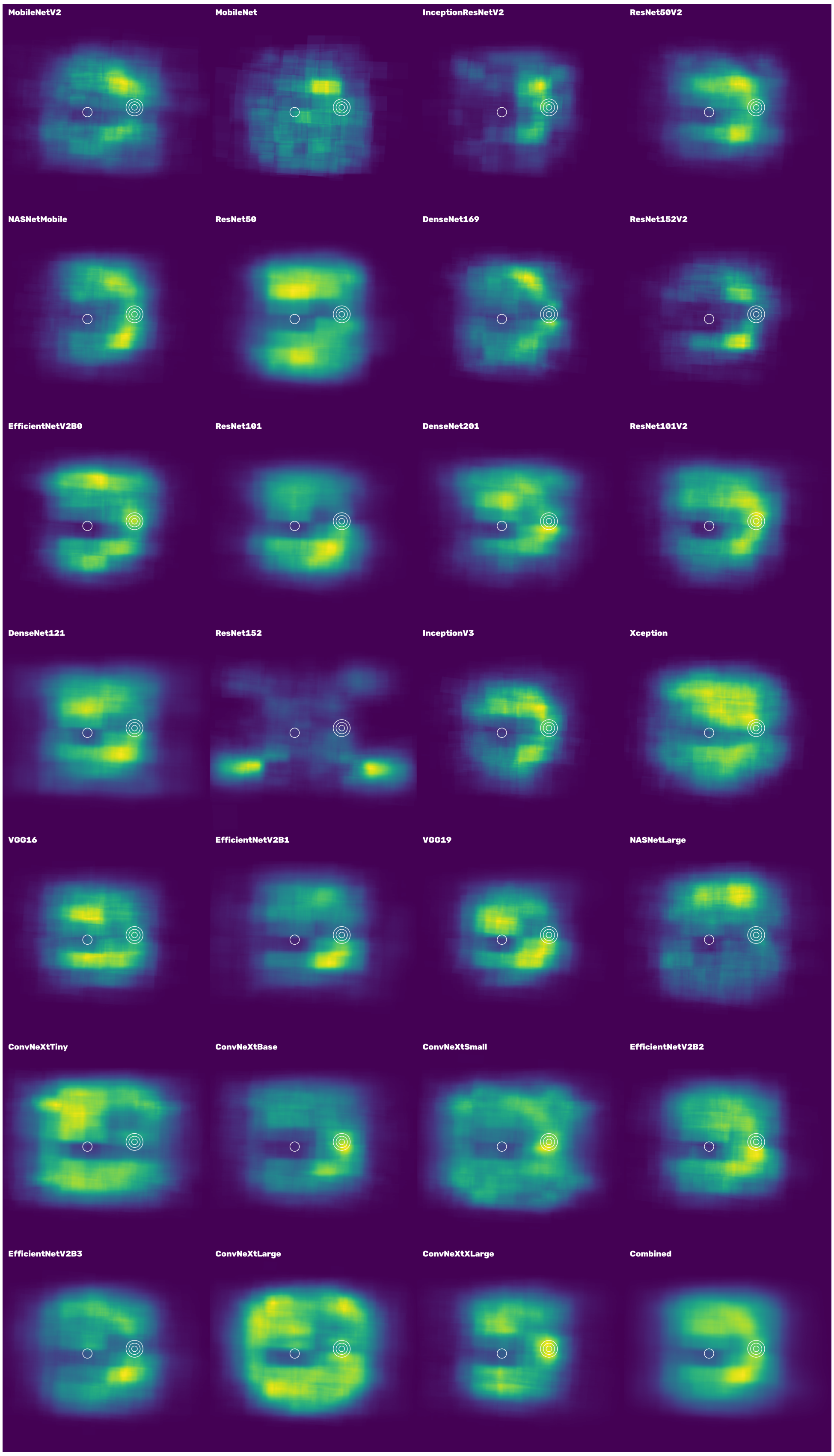

Figure 4: Collated SHAP heatmaps for "refer" models for images predicted as not having referable diabetic retinopathy. Heatmaps are ordered by Macro-F1 score in ascending order.

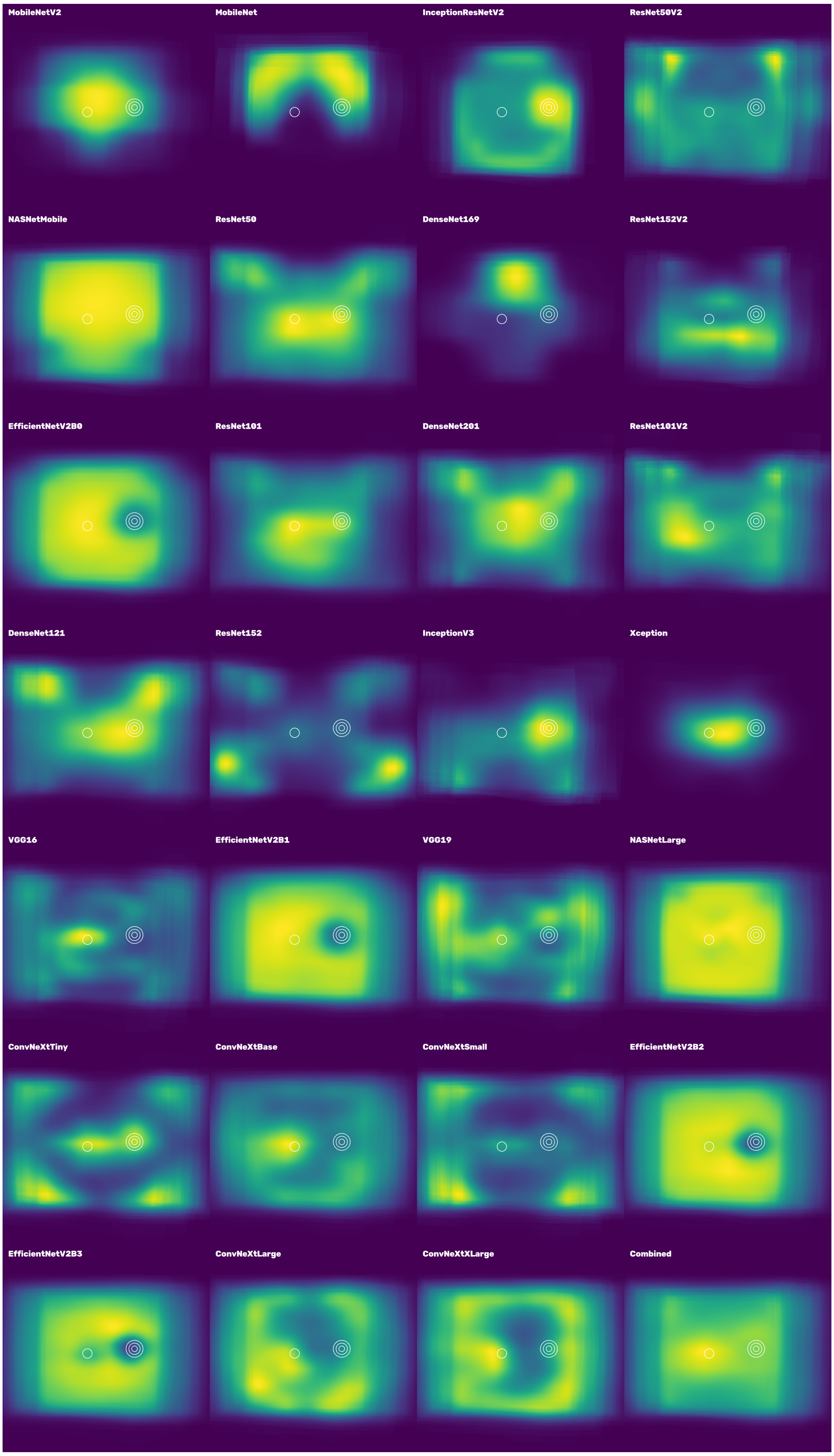

Figure 5: Collated Grad-CAM heatmaps for "refer" models for images predicted as not having referable diabetic retinopathy. Heatmaps are ordered by Macro-F1 score in ascending order.

#### 1.2 Referable diabetic retinopathy predicted

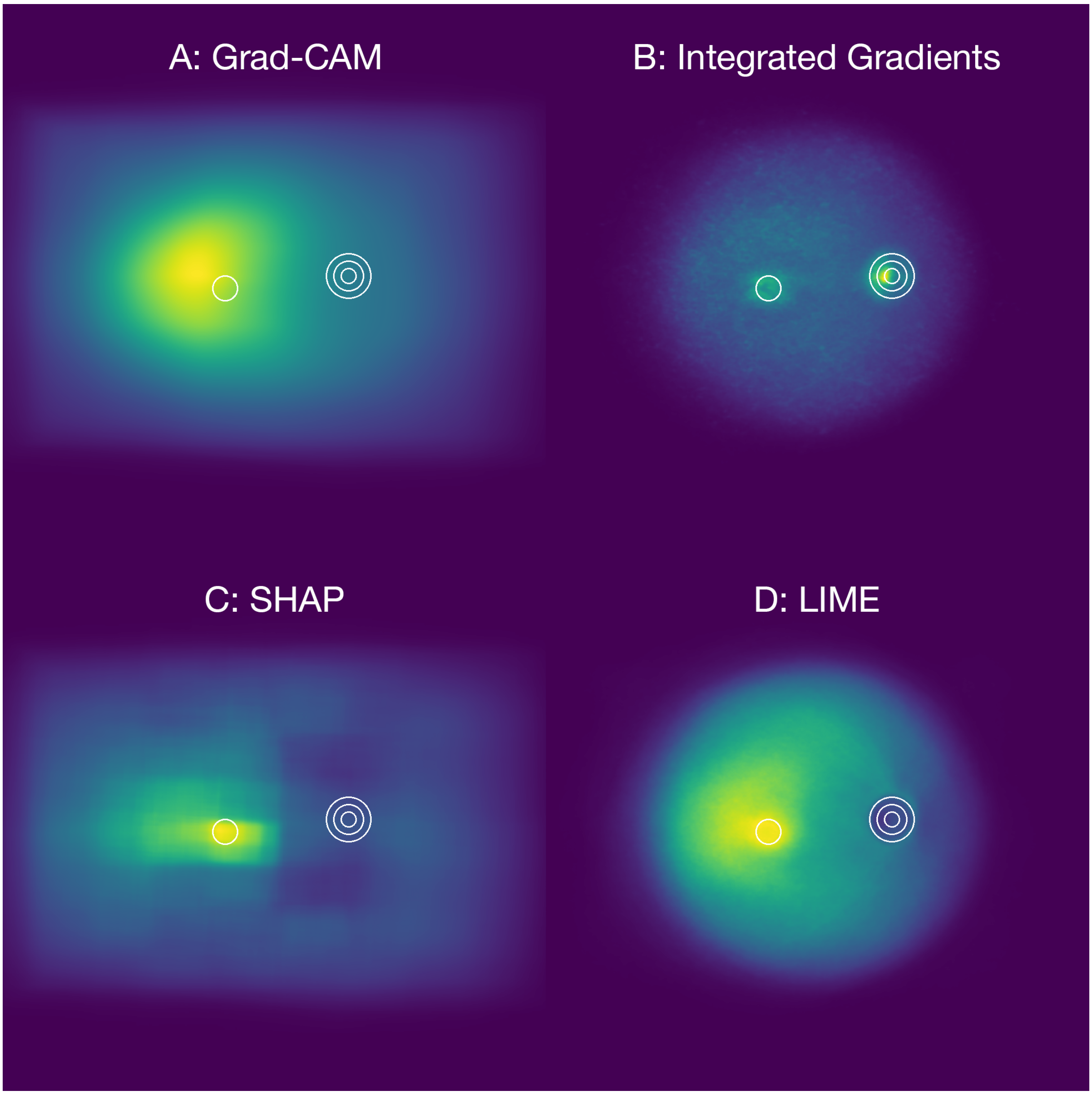

Figure 6: Aggregated “refer” model heatmaps for images predicted as having referable diabetic retinopathy using Grad-CAM (A), Integrated Gradients (B), SHAP (C) and LIME (D).

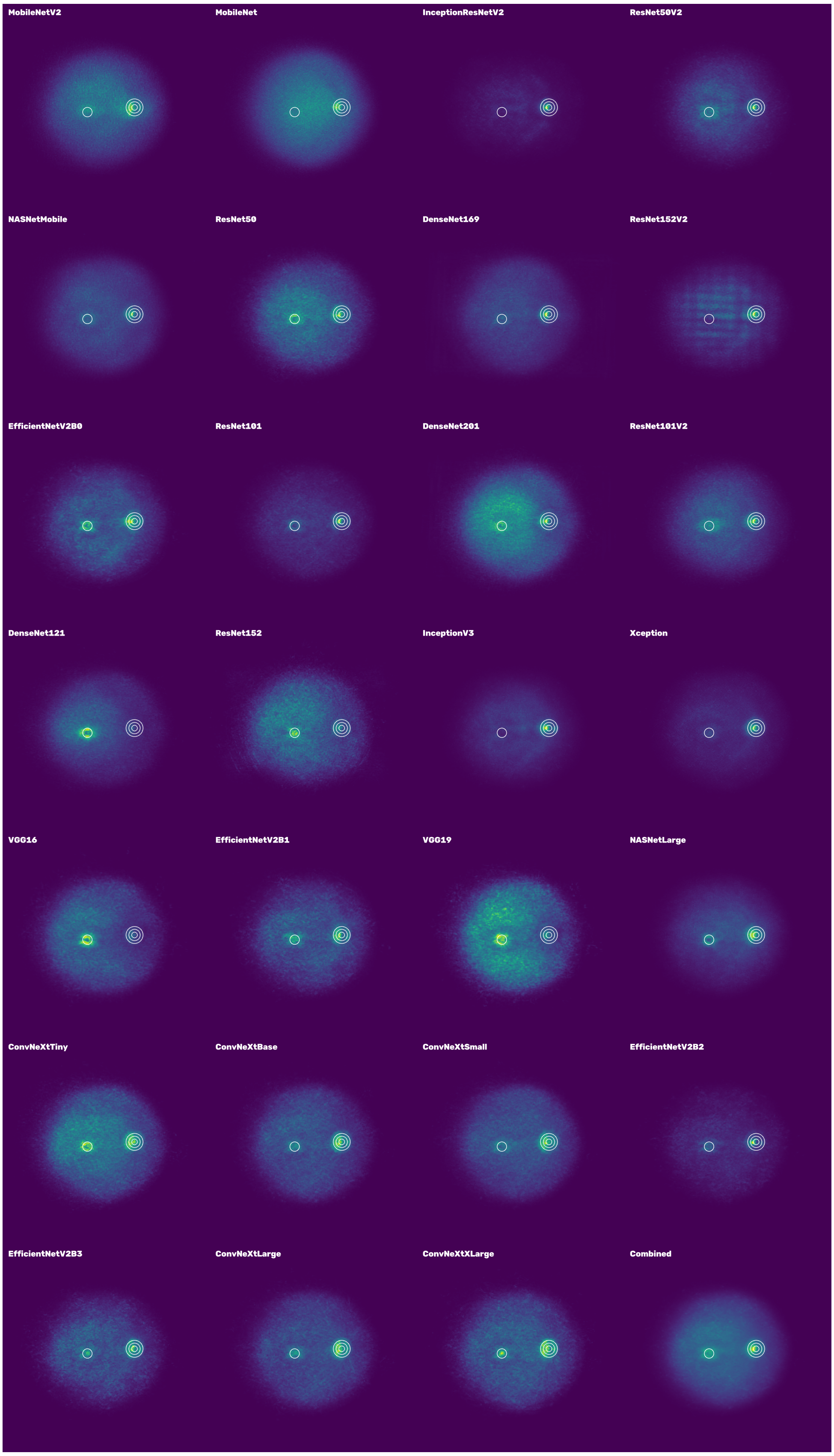

Figure 7: Collated Integrated Gradient heatmaps for "refer" models for images predicted as having referable diabetic retinopathy. Heatmaps are ordered by Macro-F1 score in ascending order.

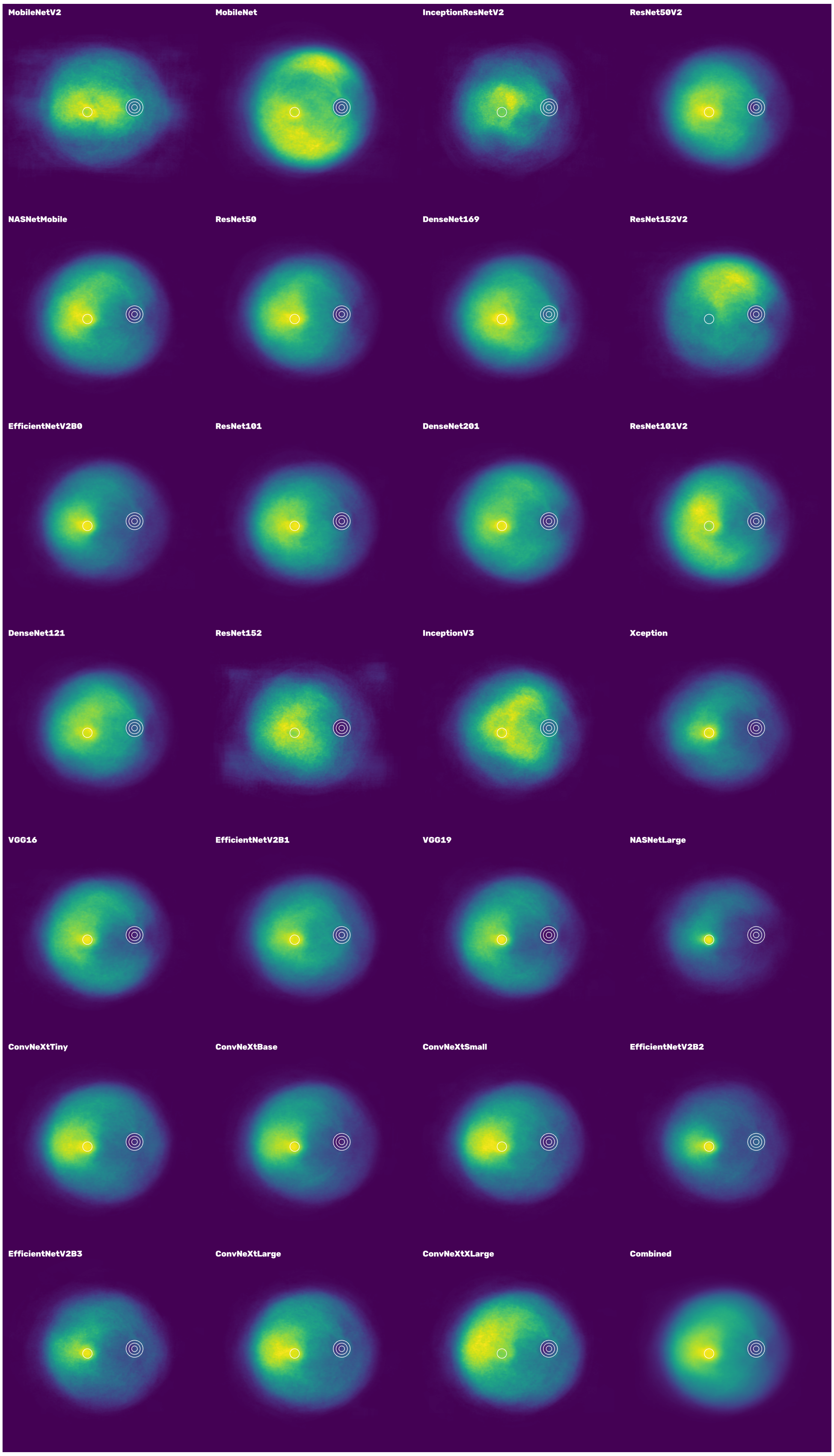

Figure 8: Collated LIME heatmaps for "refer" models for images predicted as having referable diabetic retinopathy. Heatmaps are ordered by Macro-F1 score in ascending order.

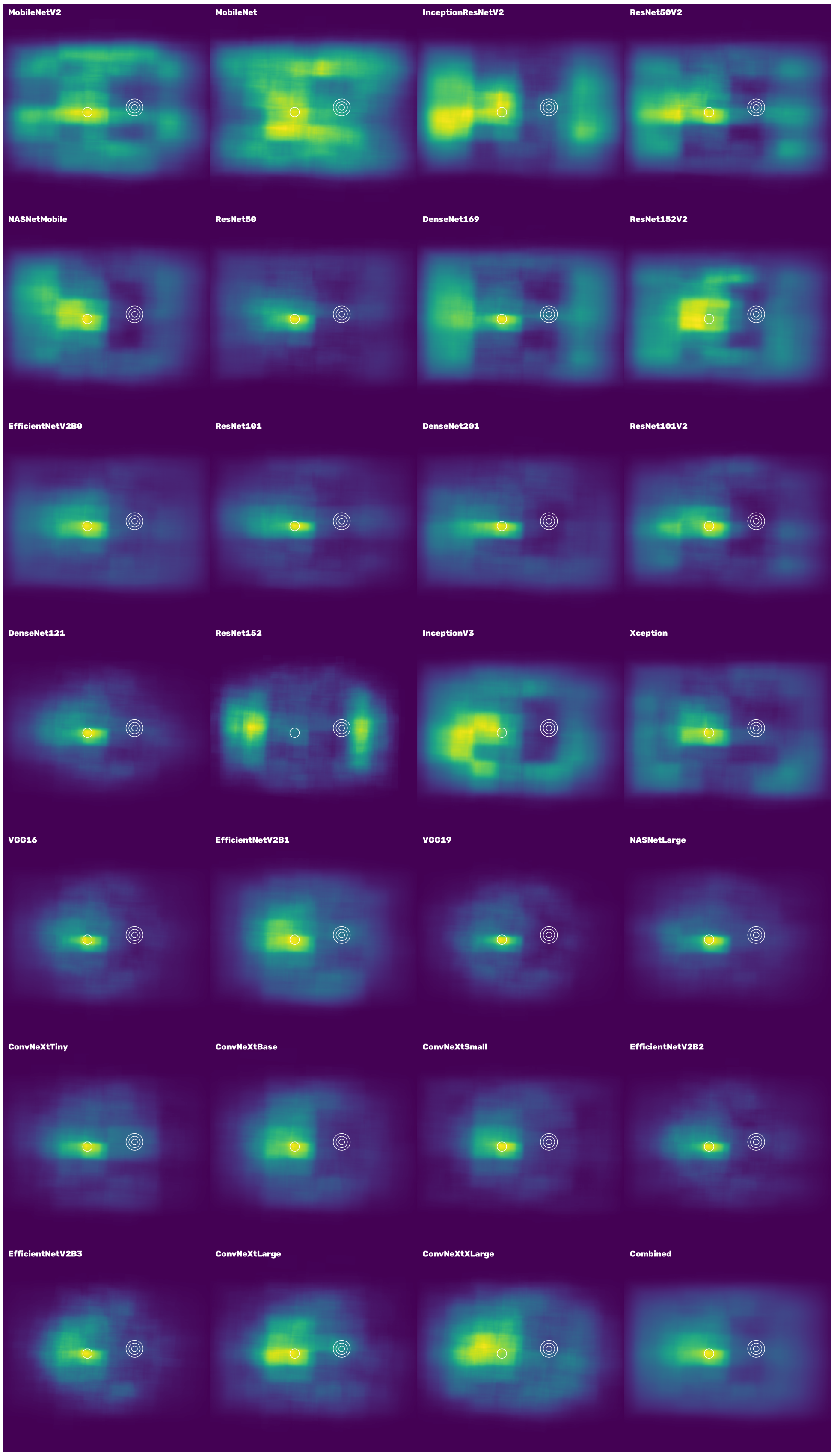

Figure 9: Collated SHAP heatmaps for "refer" models for images predicted as having referable diabetic retinopathy. Heatmaps are ordered by Macro-F1 score in ascending order.

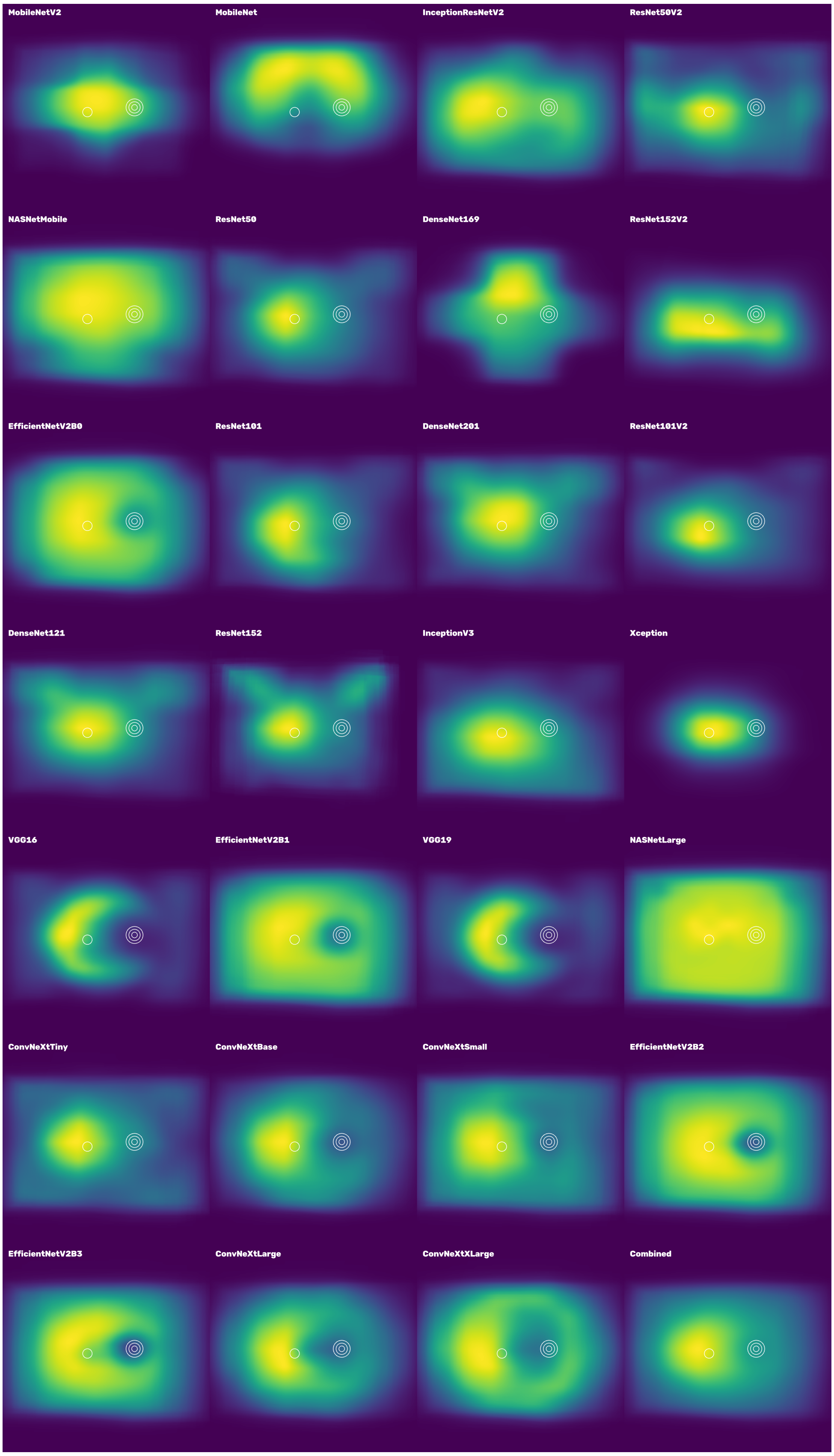

Figure 10: Collated Grad-CAM heatmaps for ”refer” models for images predicted as having referable diabetic retinopathy. Heatmaps are ordered by Macro-F1 score in ascending order.

#### 1.3 Diabetic retinopathy feature coverage

##### 1.3.1 Grad-CAM

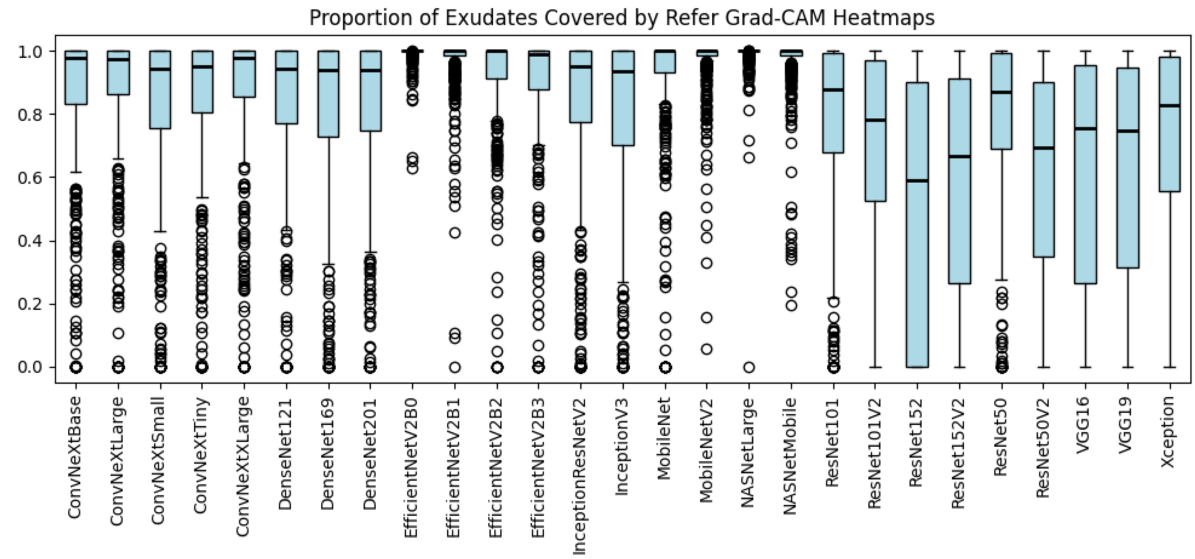

Figure 11: Proportion of exudates covered by "refer" Grad-CAM heatmaps.

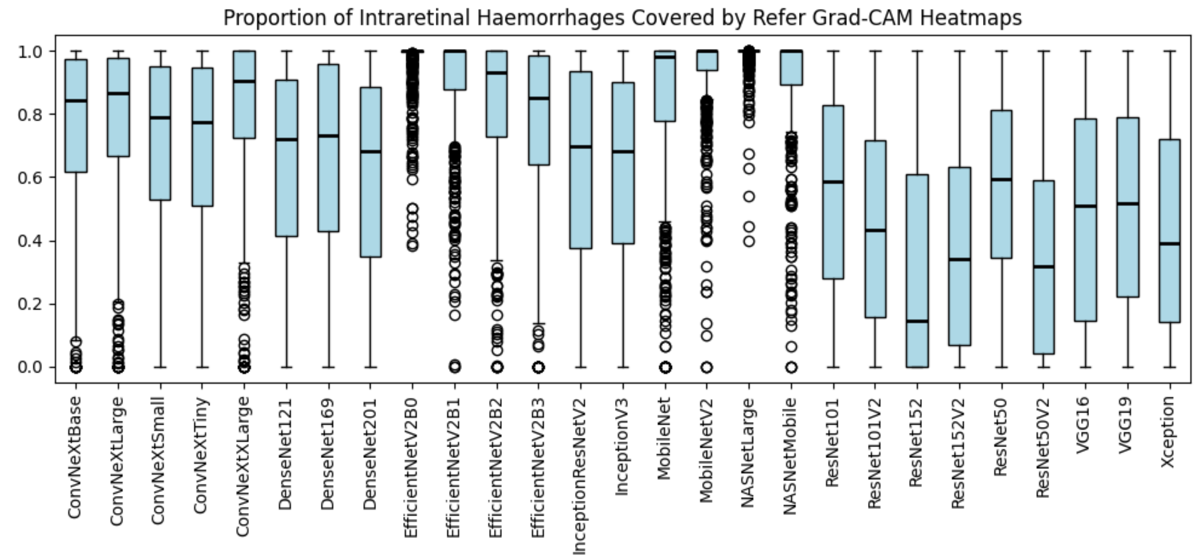

Figure 12: Proportion of intraretinal haemorrhages covered by "refer" Grad-CAM heatmaps.

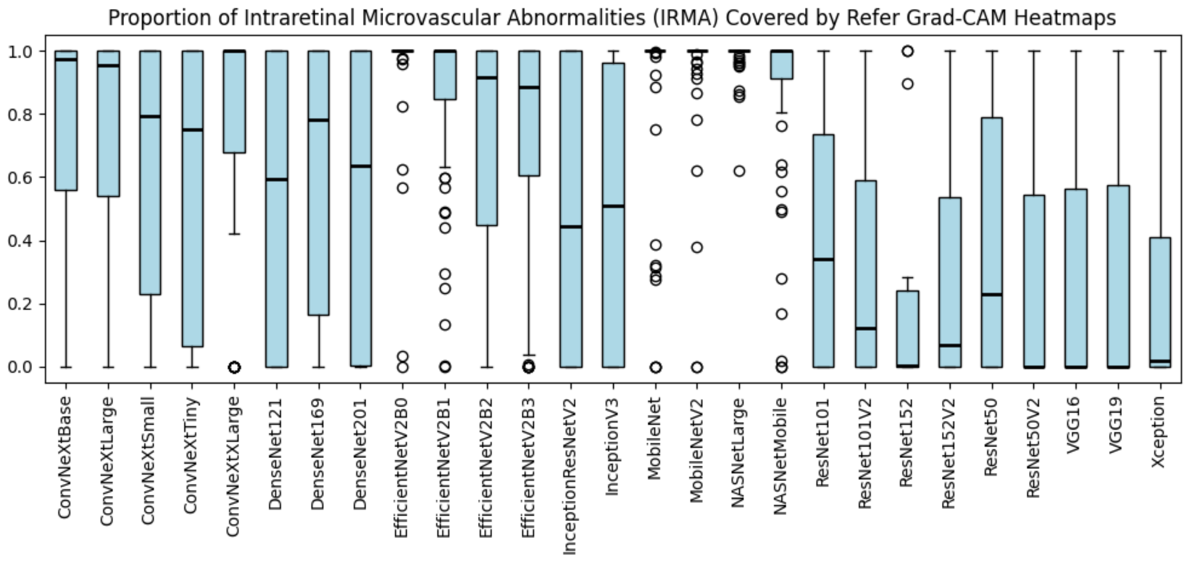

Figure 13: Proportion of intraretinal microvascular abnormalities (IRMA) covered by "refer" Grad-CAM heatmaps.

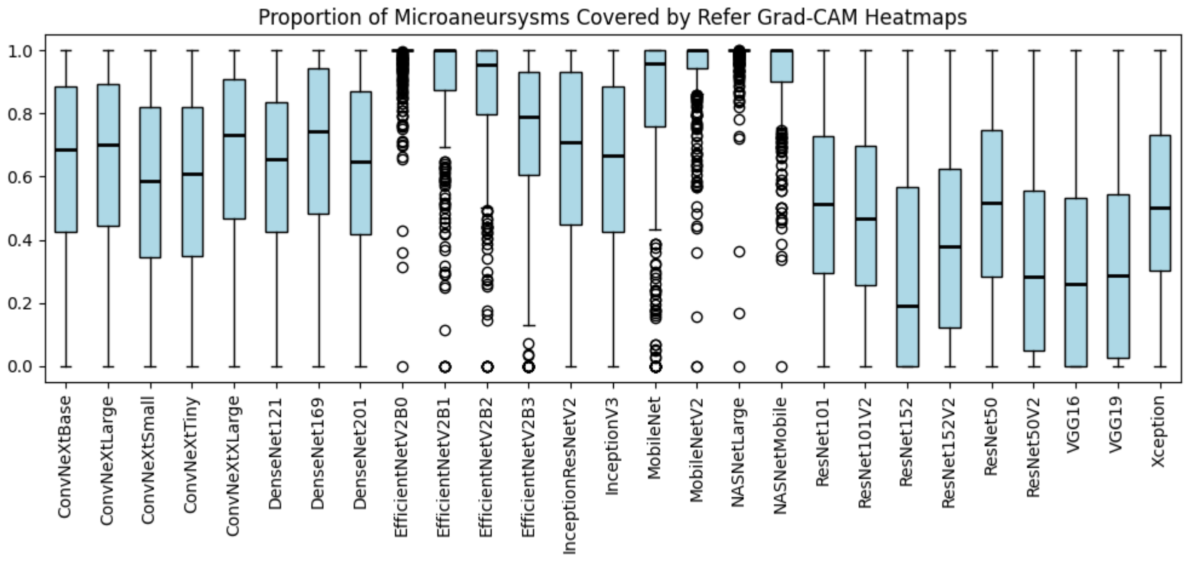

Figure 14: Proportion of microaneurysms covered by "refer" Grad-CAM heatmaps.

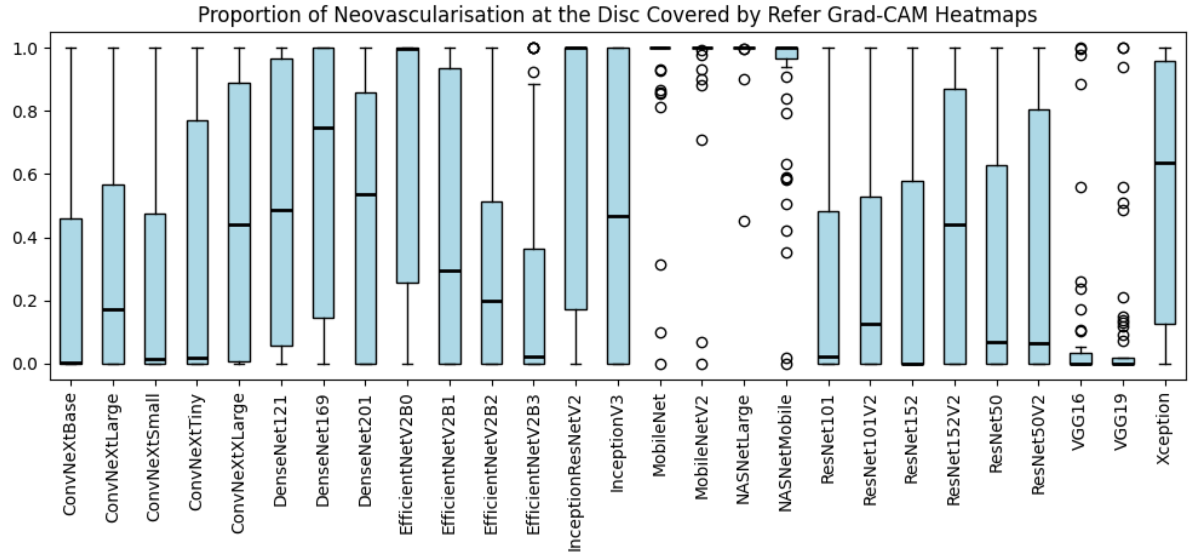

Figure 15: Proportion of neovascularisation at the disc covered by “refer” Grad-CAM heatmaps.

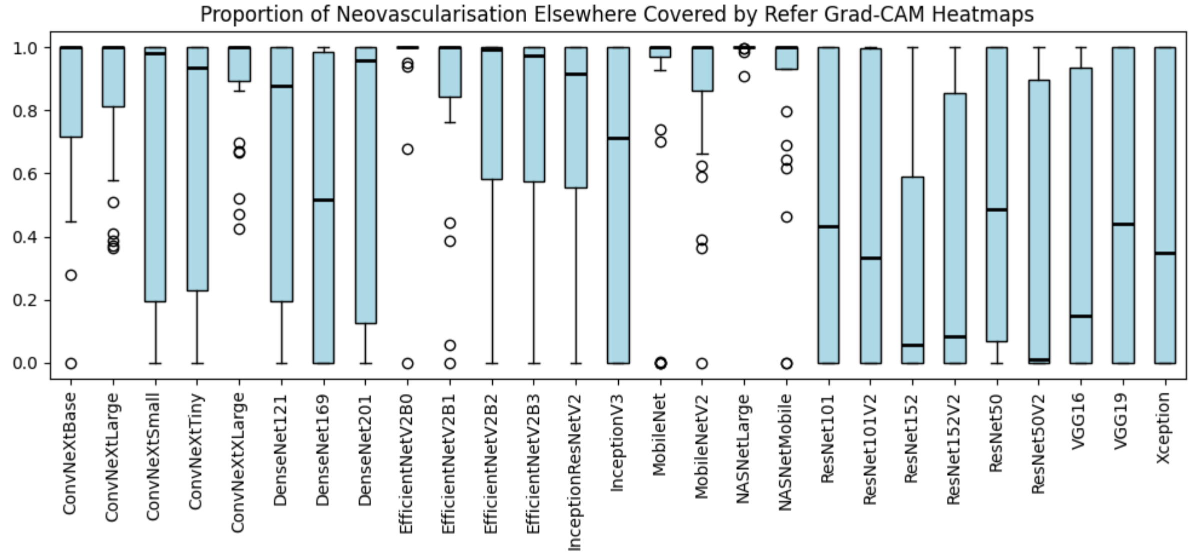

Figure 16: Proportion of neovascularisation elsewhere covered by “refer” Grad-CAM heatmaps.

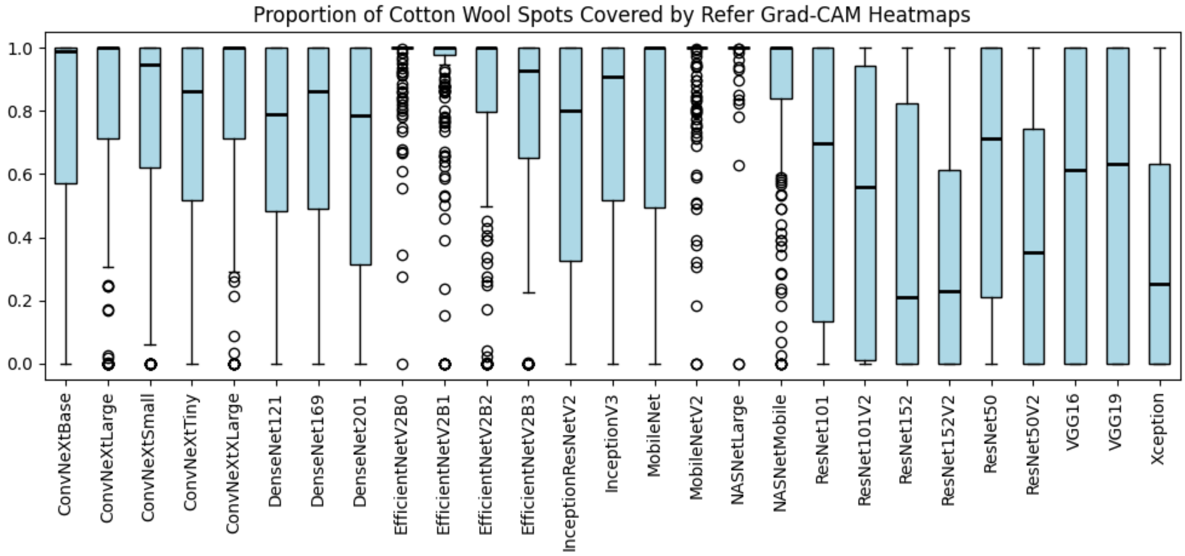

Figure 17: Proportion of cotton wool spots covered by "refer" Grad-CAM heatmaps.

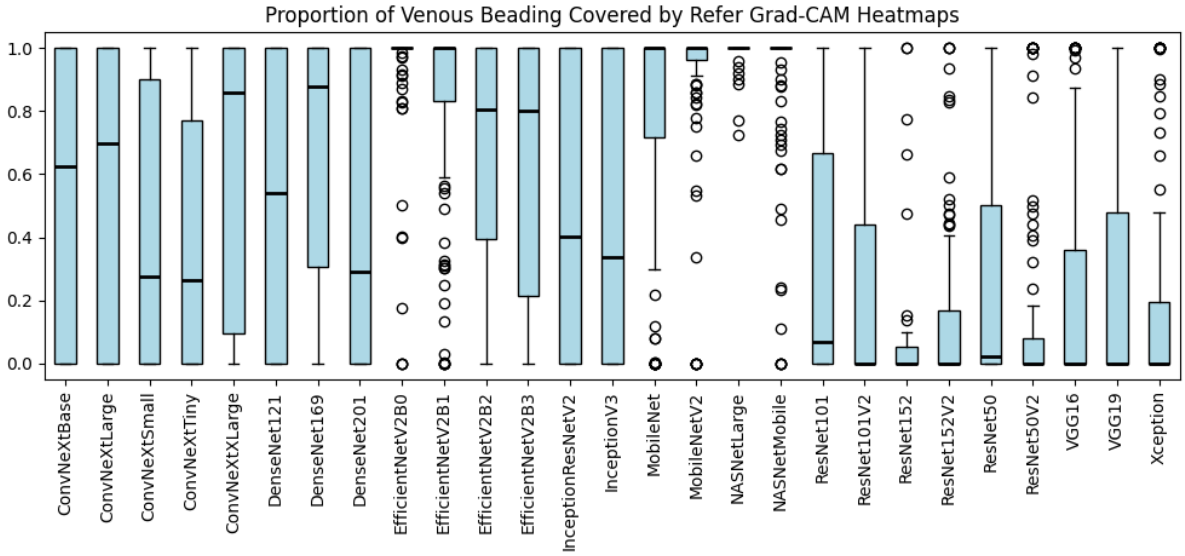

Figure 18: Proportion of venous beading covered by "refer" Grad-CAM heatmaps.

##### 1.3.2 Integrated Gradients

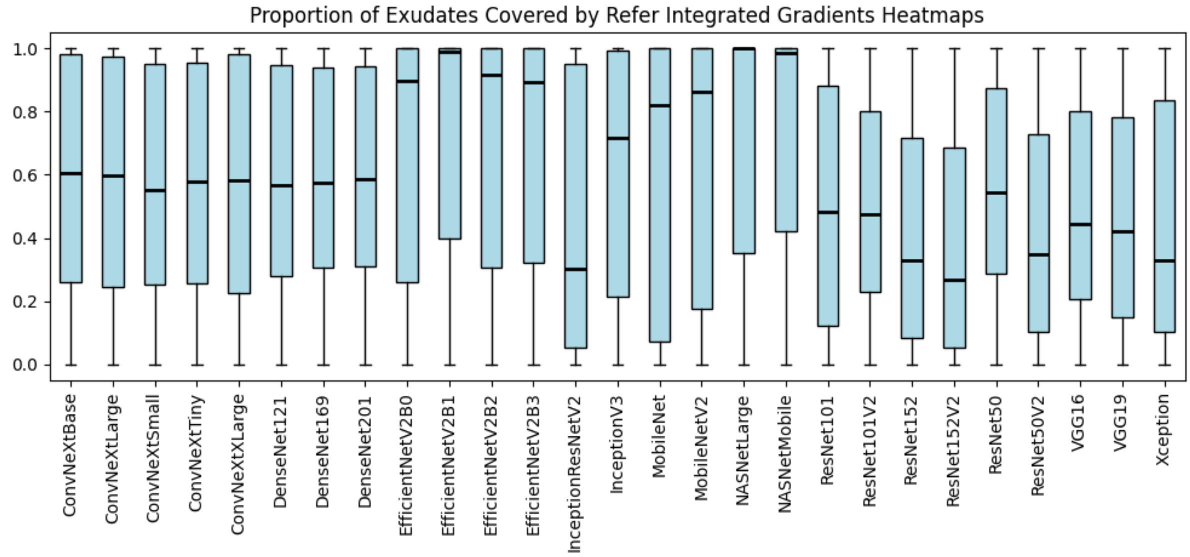

Figure 19: Proportion of exudates covered by "refer" Integrated Gradient heatmaps.

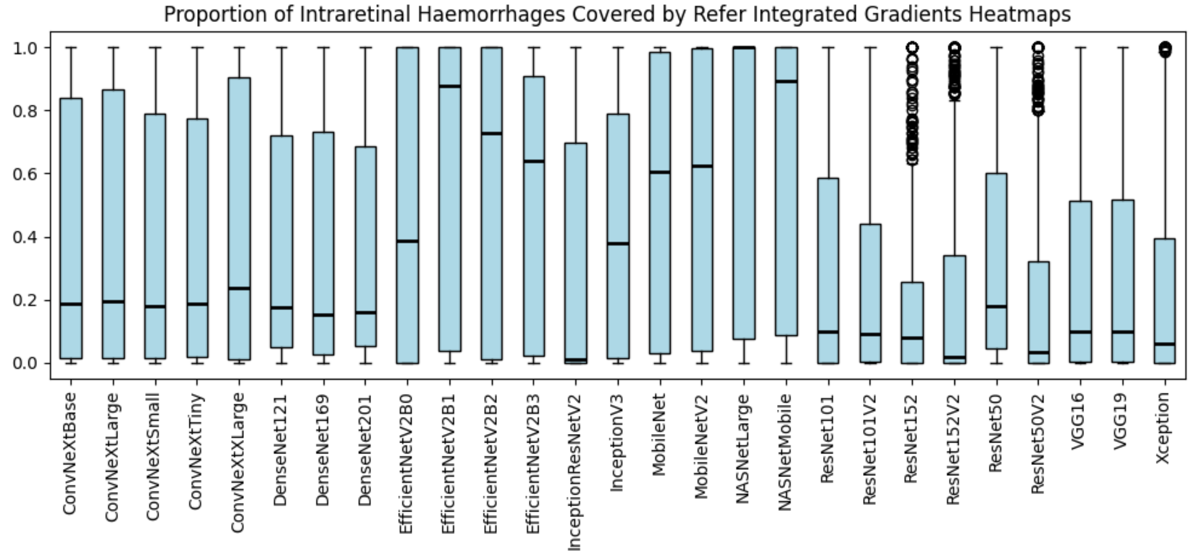

Figure 20: Proportion of intraretinal haemorrhages covered by "refer" Integrated Gradient heatmaps.

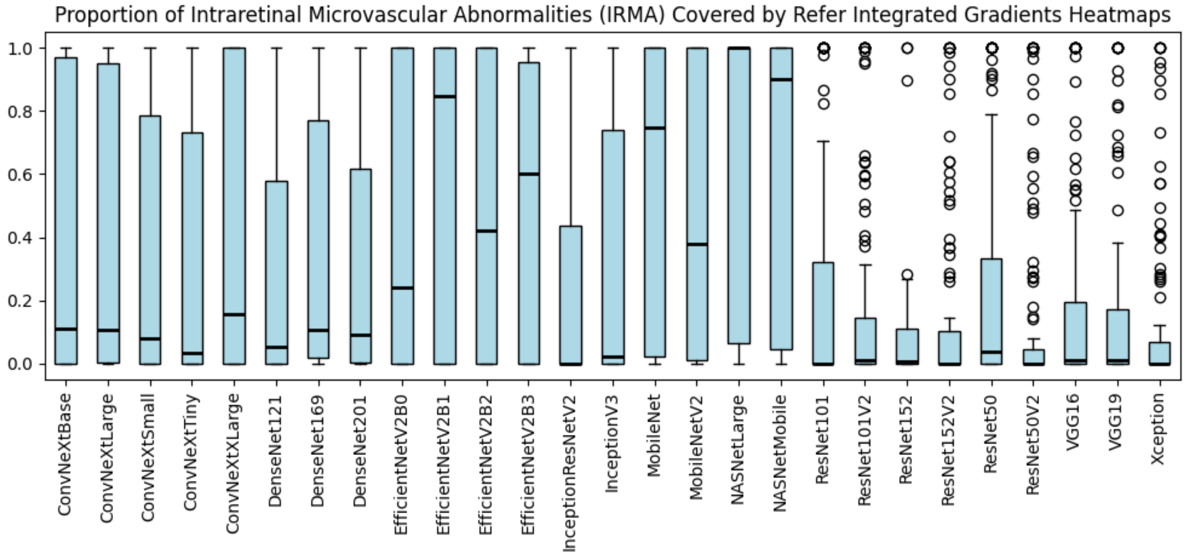

Figure 21: Proportion of intraretinal microvascular abnormalities (IRMA) covered by "refer" Integrated Gradient heatmaps.

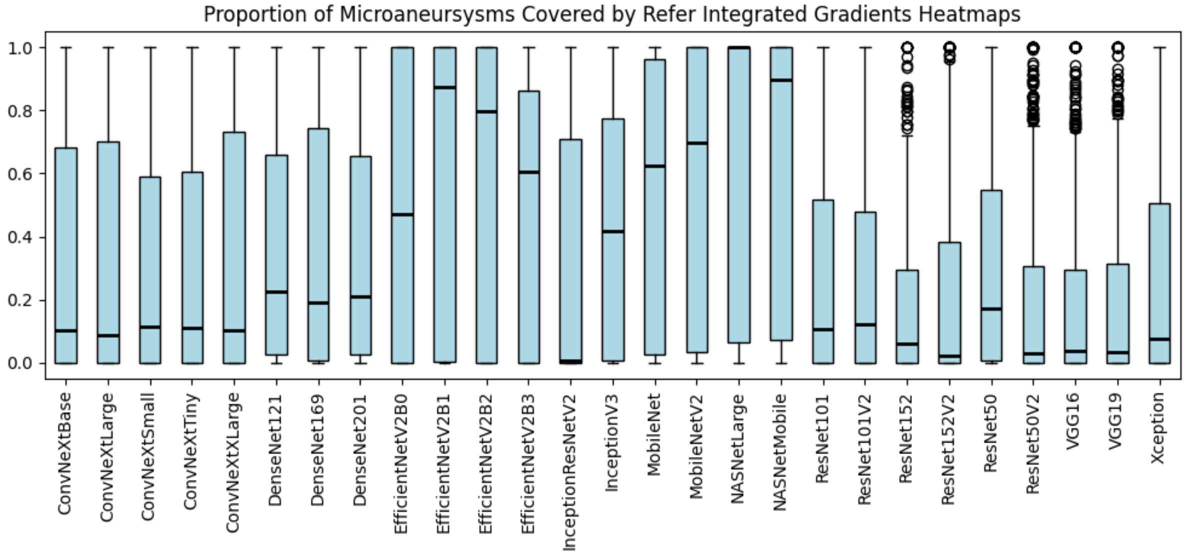

Figure 22: Proportion of microaneurysms covered by "refer" Integrated Gradient heatmaps.

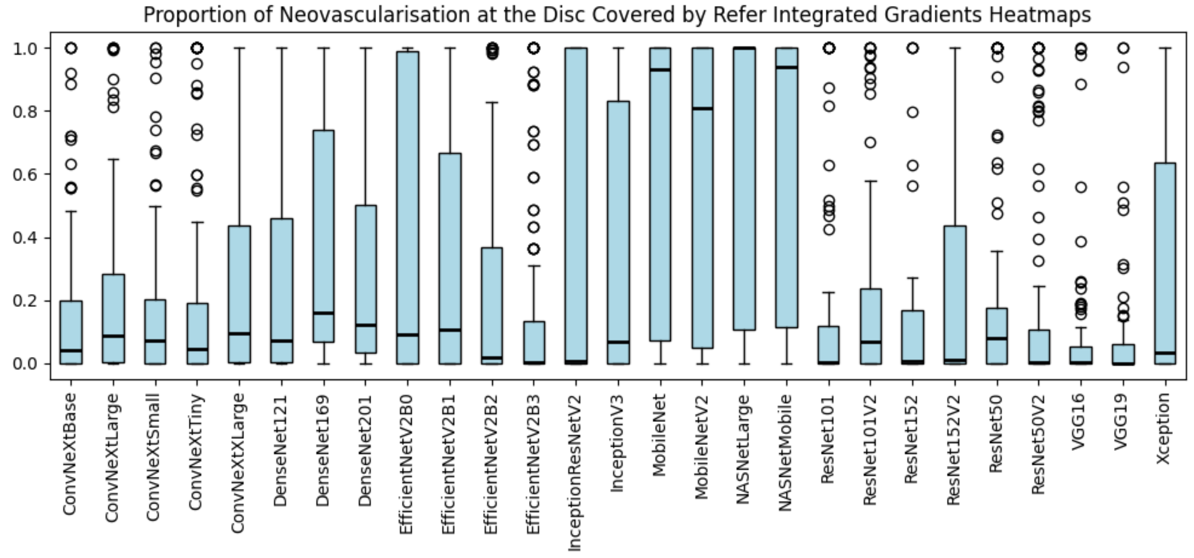

Figure 23: Proportion of neovascularisation at the disc covered by “refer” Integrated Gradient heatmaps.

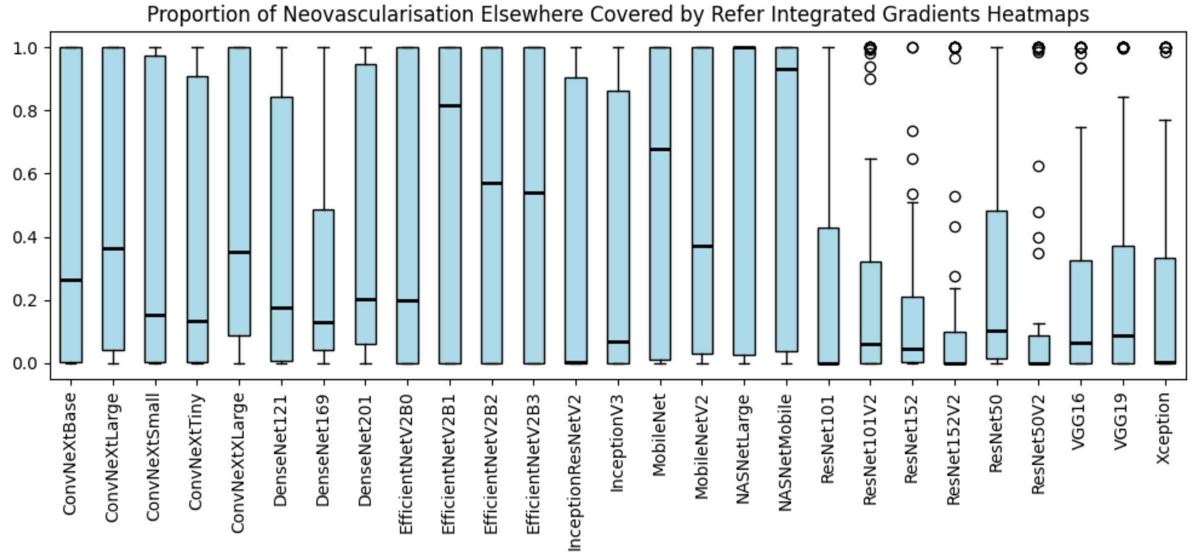

Figure 24: Proportion of neovascularisation elsewhere covered by “refer” Integrated Gradient heatmaps.

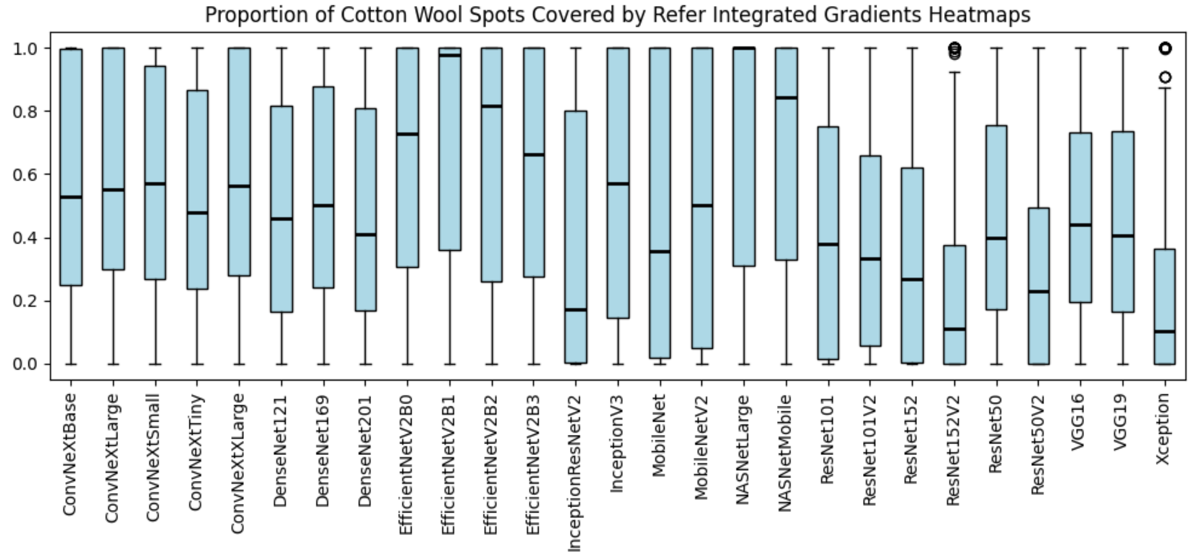

Figure 25: Proportion of cotton wool spots covered by “refer” Integrated Gradient heatmaps.

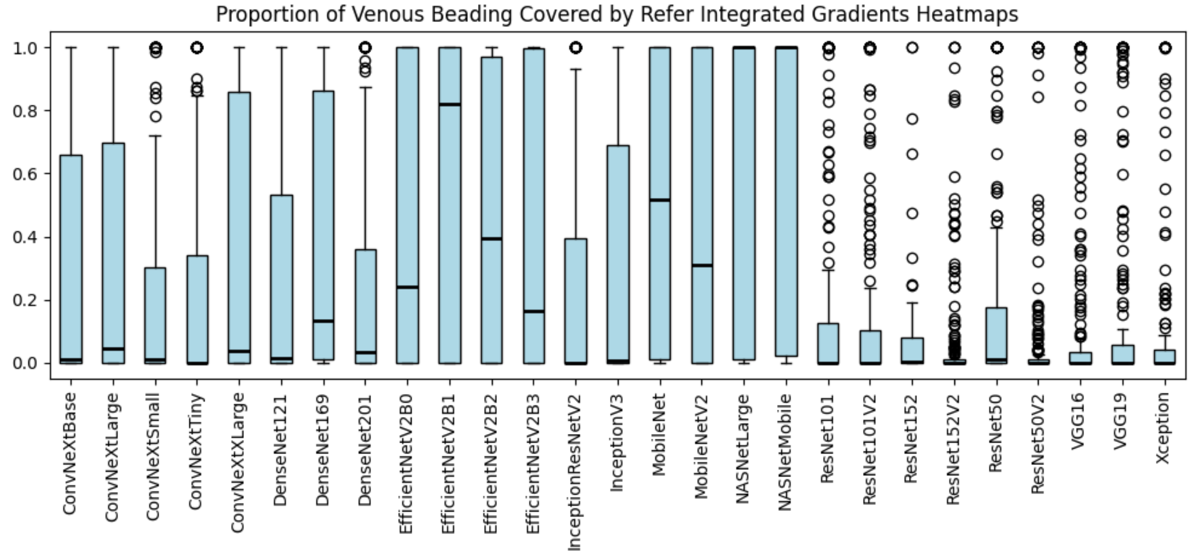

Figure 26: Proportion of venous beading covered by “refer” Integrated Gradient heatmaps.

##### 1.3.3 LIME

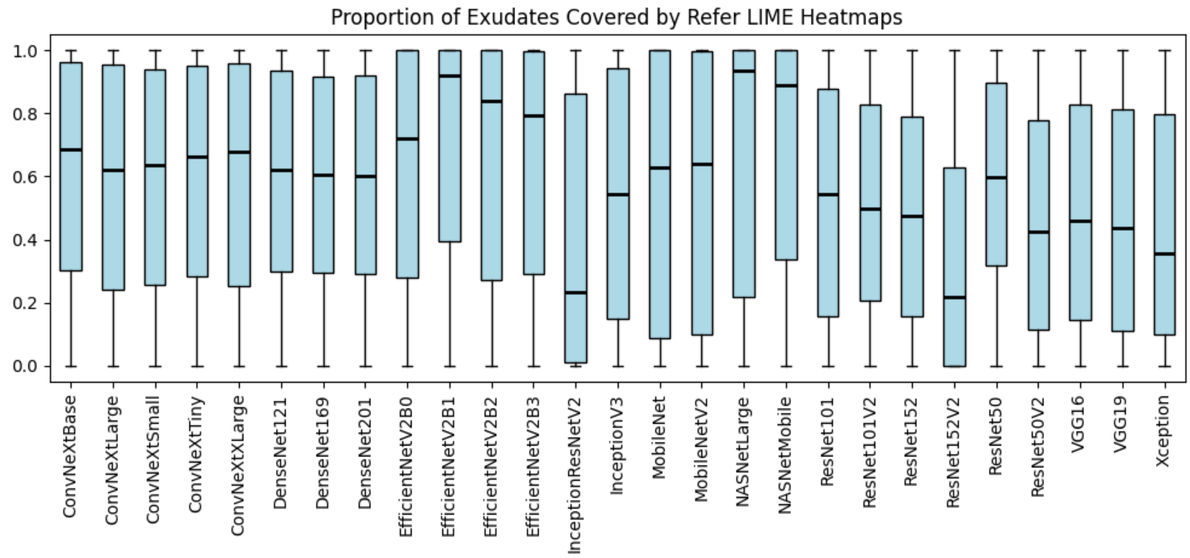

Figure 27: Proportion of exudates covered by “refer” LIME heatmaps.

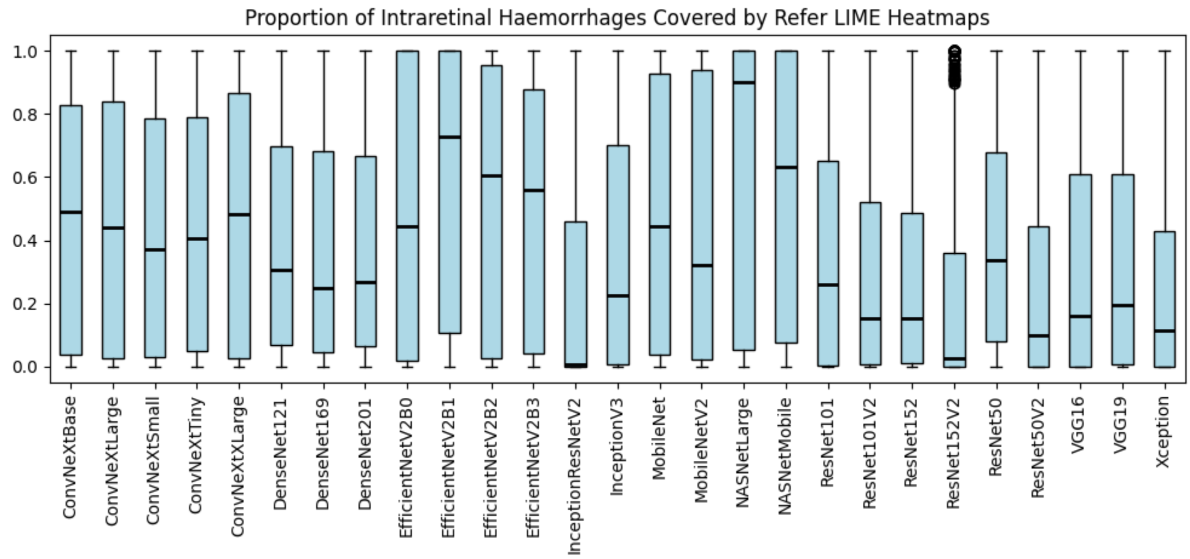

Figure 28: Proportion of intraretinal haemorrhages covered by “refer” LIME heatmaps.

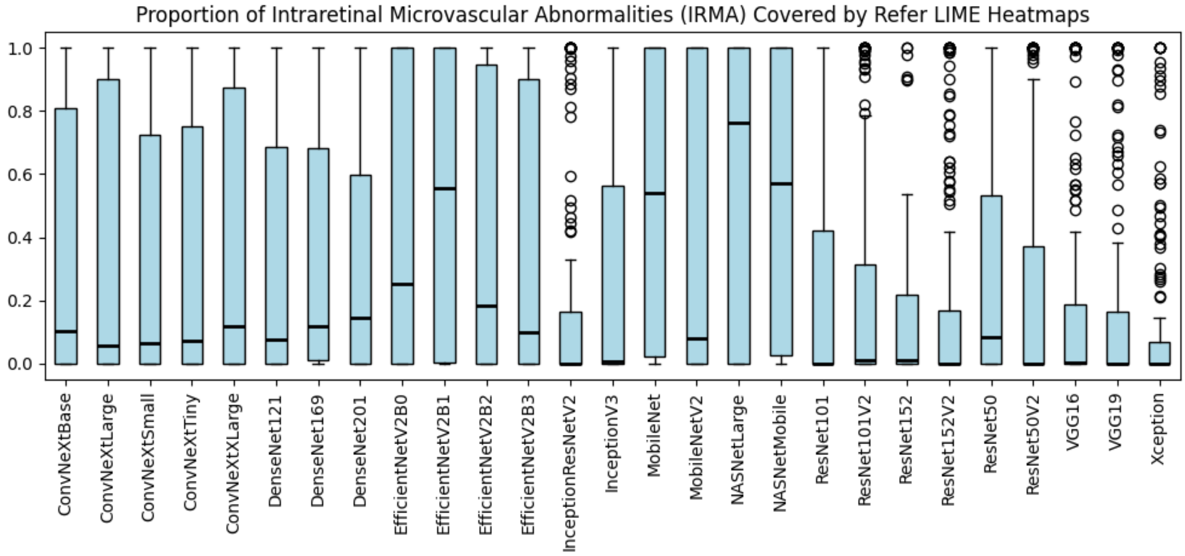

Figure 29: Proportion of intraretinal microvascular abnormalities (IRMA) covered by “refer” LIME heatmaps.

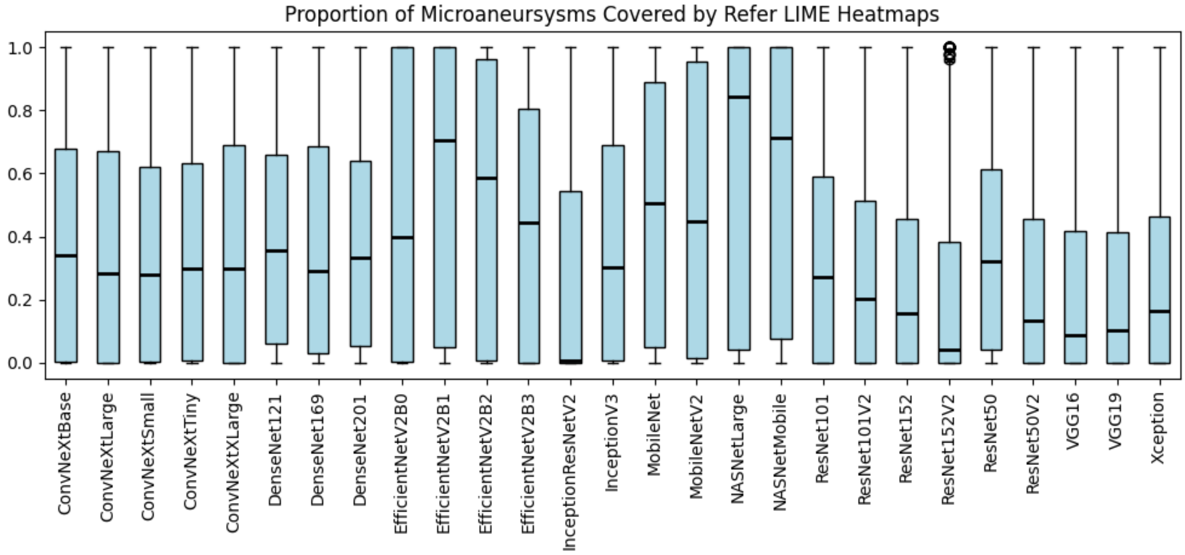

Figure 30: Proportion of microaneurysms covered by “refer” LIME heatmaps.

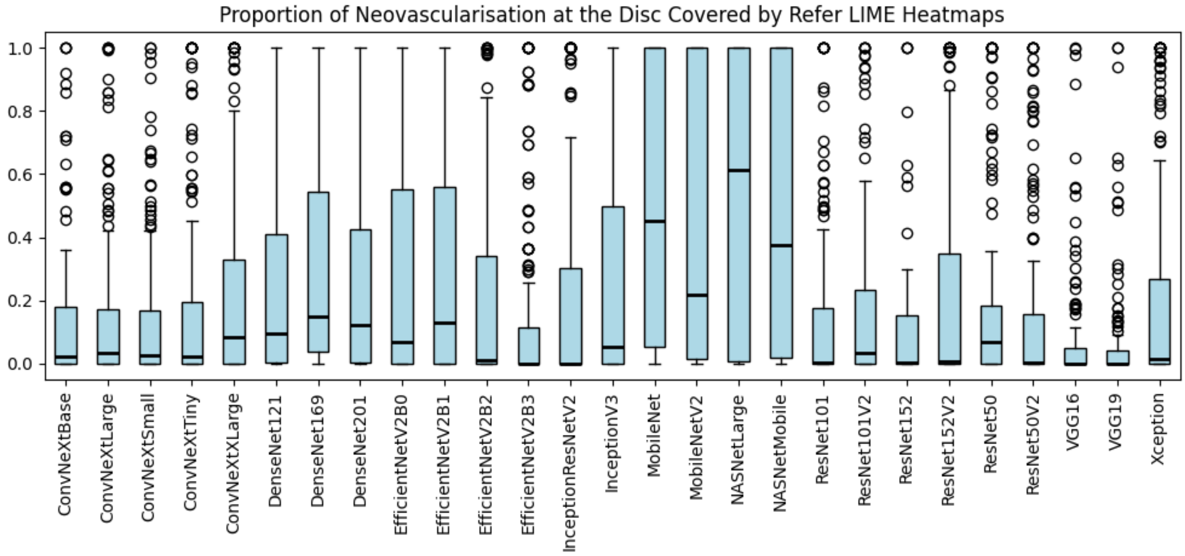

Figure 31: Proportion of neovascularisation at the disc covered by “refer” LIME heatmaps.

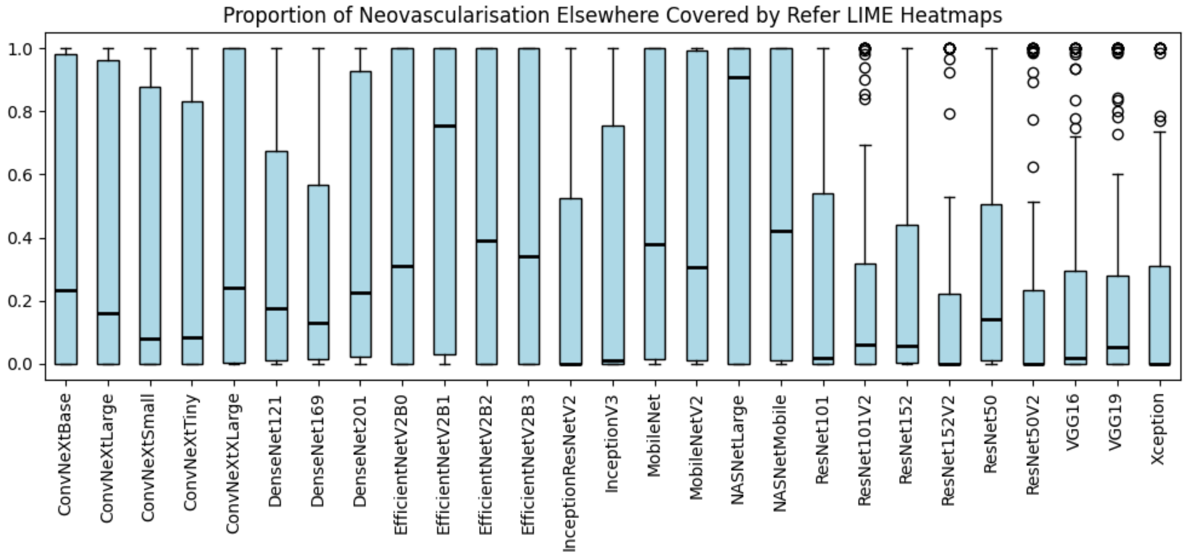

Figure 32: Proportion of neovascularisation elsewhere covered by “refer” LIME heatmaps.

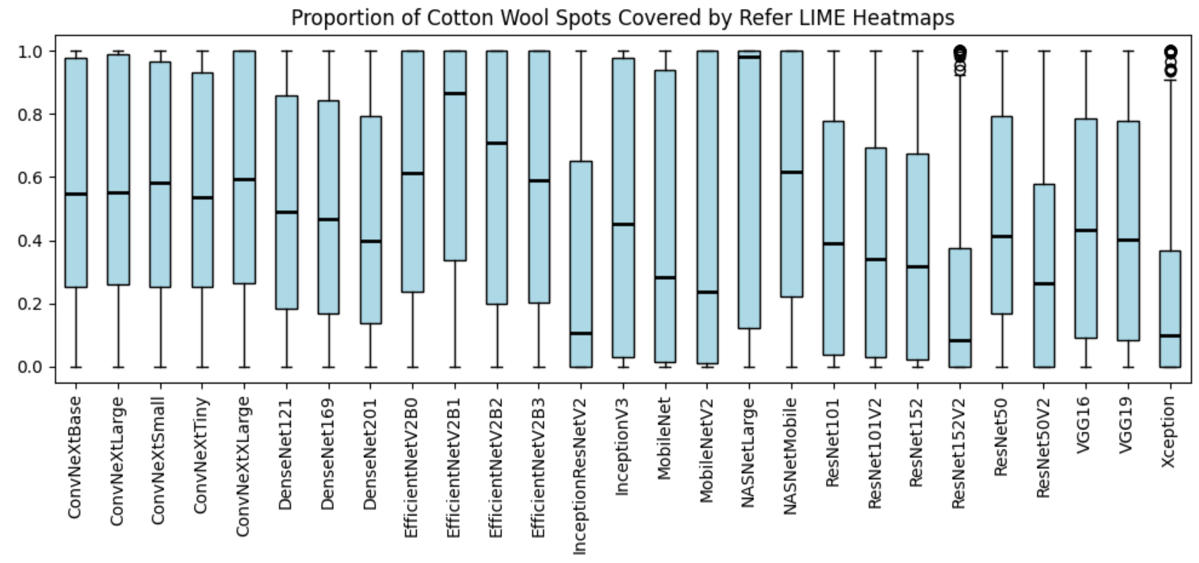

Figure 33: Proportion of cotton wool spots covered by “refer” LIME heatmaps.

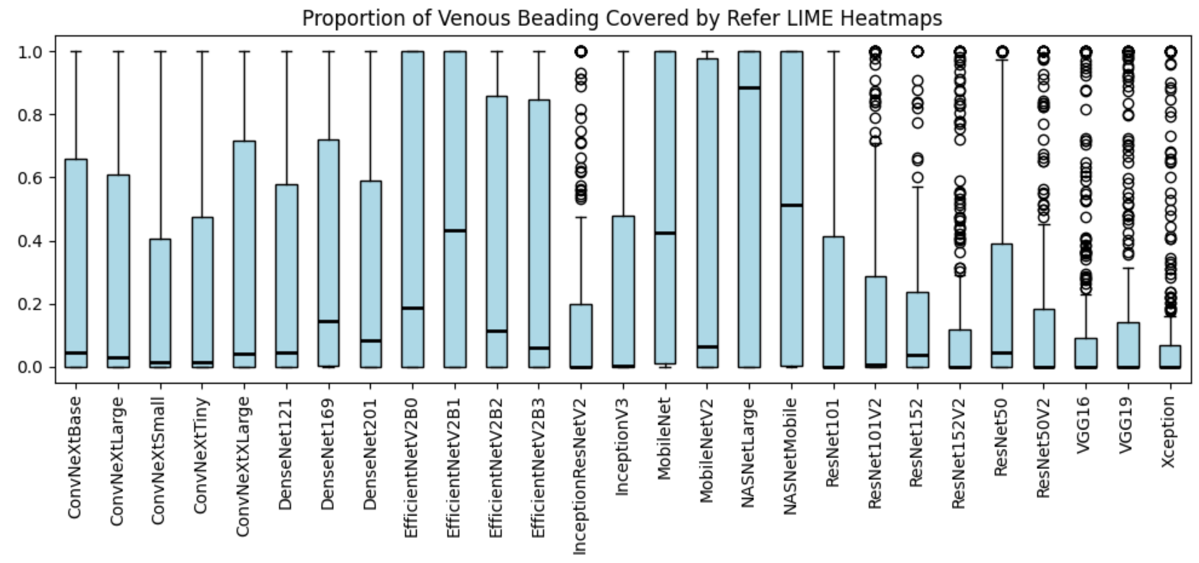

Figure 34: Proportion of venous beading covered by “refer” LIME heatmaps.

##### 1.3.4 SHAP

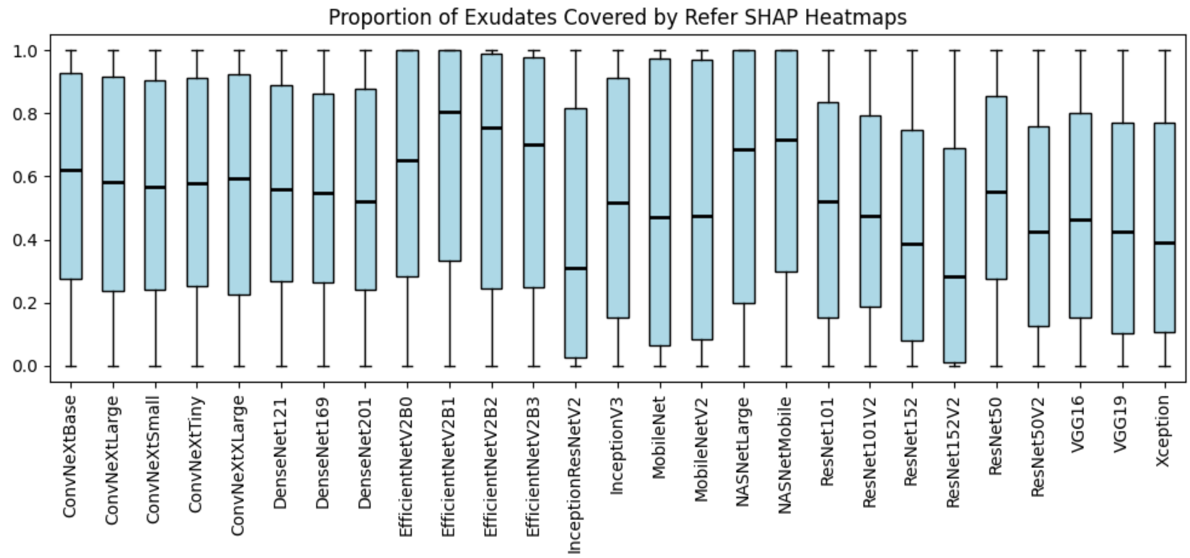

Figure 35: Proportion of exudates covered by “refer” SHAP heatmaps.

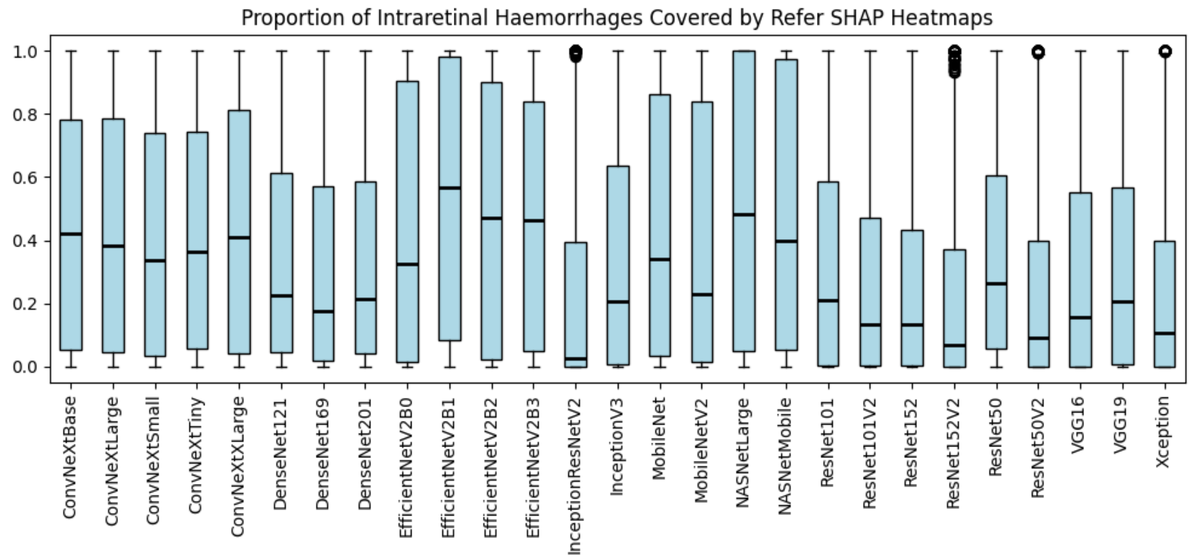

Figure 36: Proportion of intraretinal haemorrhages covered by “refer” SHAP heatmaps.

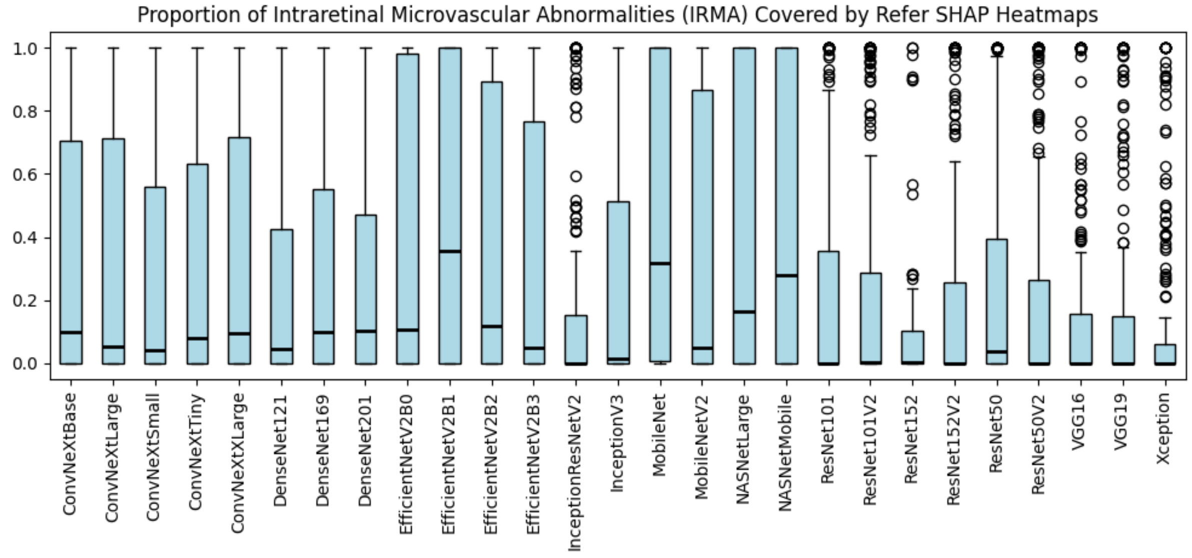

Figure 37: Proportion of intraretinal microvascular abnormalities (IRMA) covered by “refer” SHAP heatmaps.

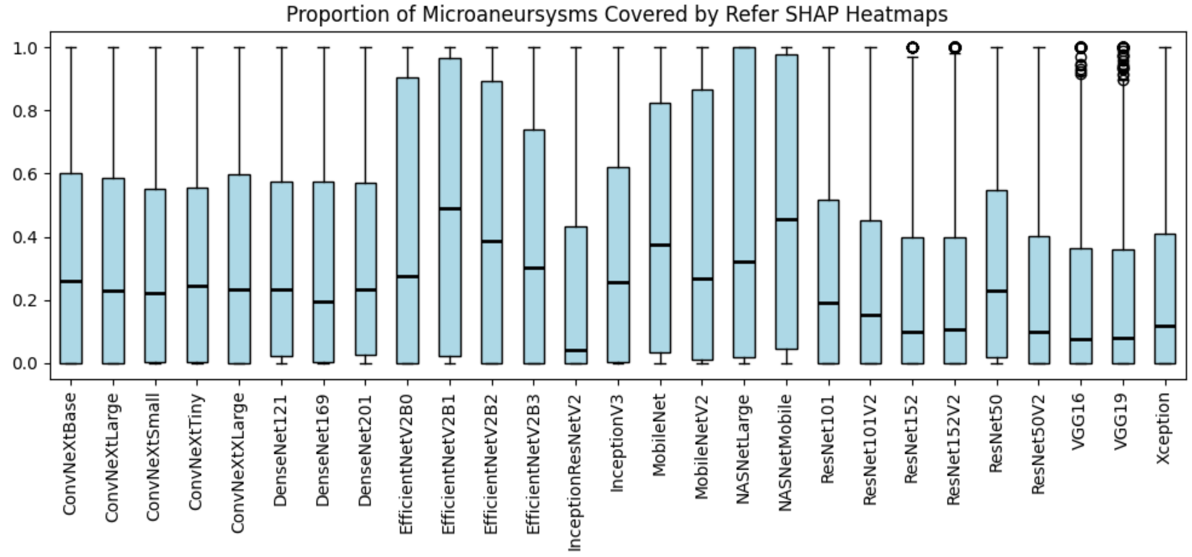

Figure 38: Proportion of microaneurysms covered by “refer” SHAP heatmaps.

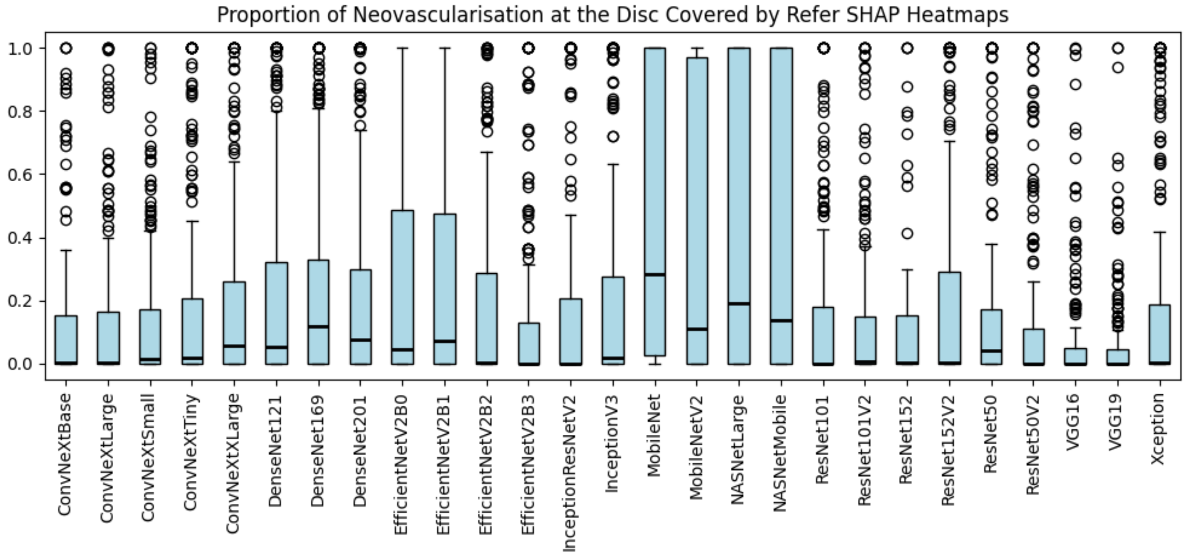

Figure 39: Proportion of neovascularisation at the disc covered by “refer” SHAP heatmaps.

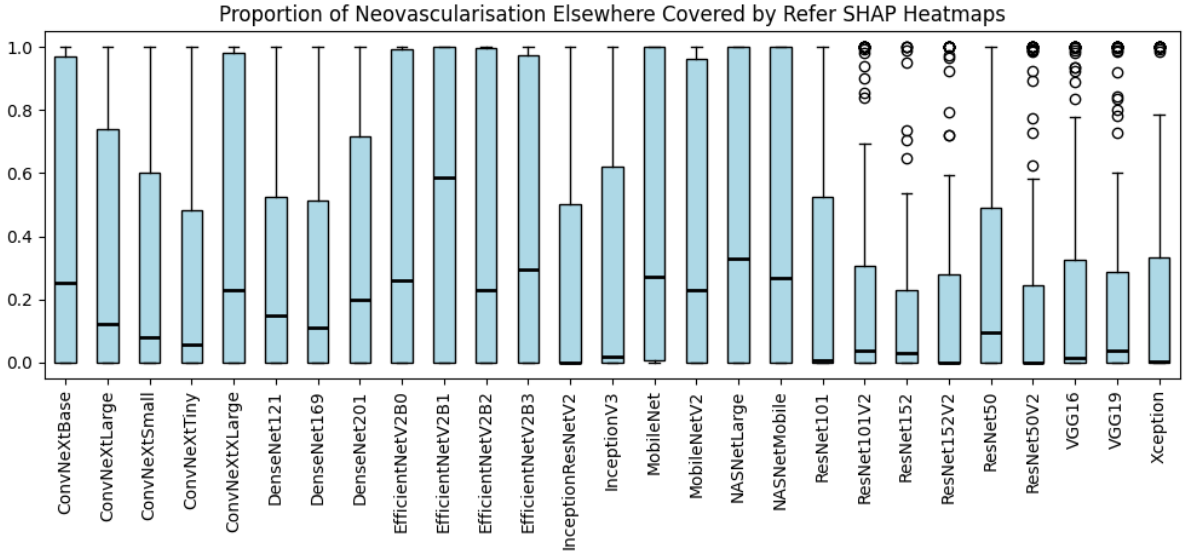

Figure 40: Proportion of neovascularisation elsewhere covered by “refer” SHAP heatmaps.

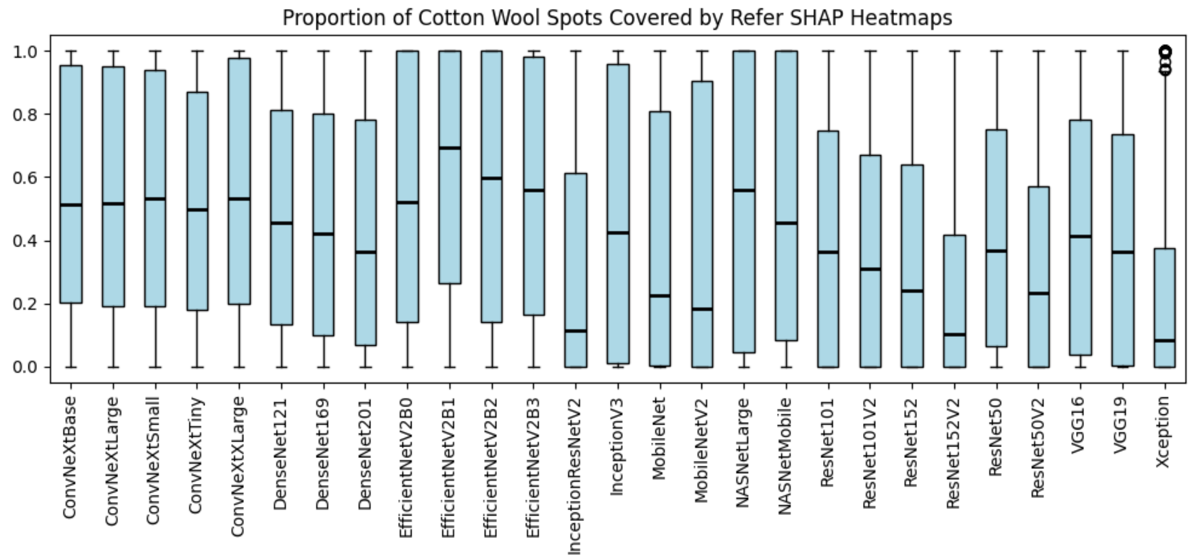

Figure 41: Proportion of cotton wool spots covered by "refer" SHAP heatmaps.

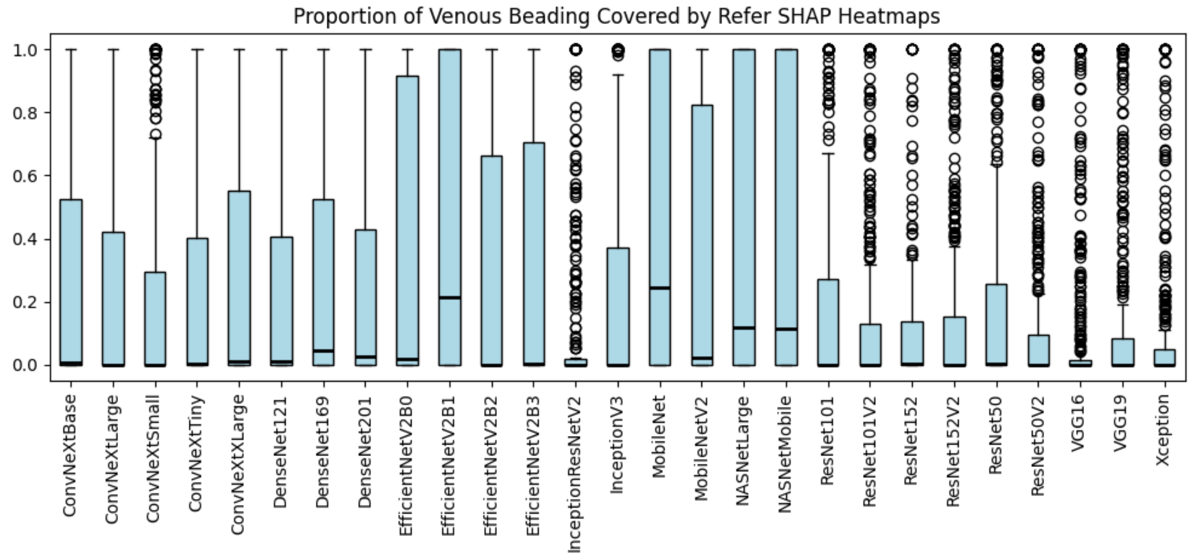

Figure 42: Proportion of venous beading covered by "refer" SHAP heatmaps.

#### 1.4 Heatmap variability

The following images represent the standard deviation of pixel values after bootstrapping with 10,000 iterations. Two heatmaps are presented: the standard deviation relative to the full dynamic range (e.g. SD / 255), and a normalised version to make the patterns easier to see.

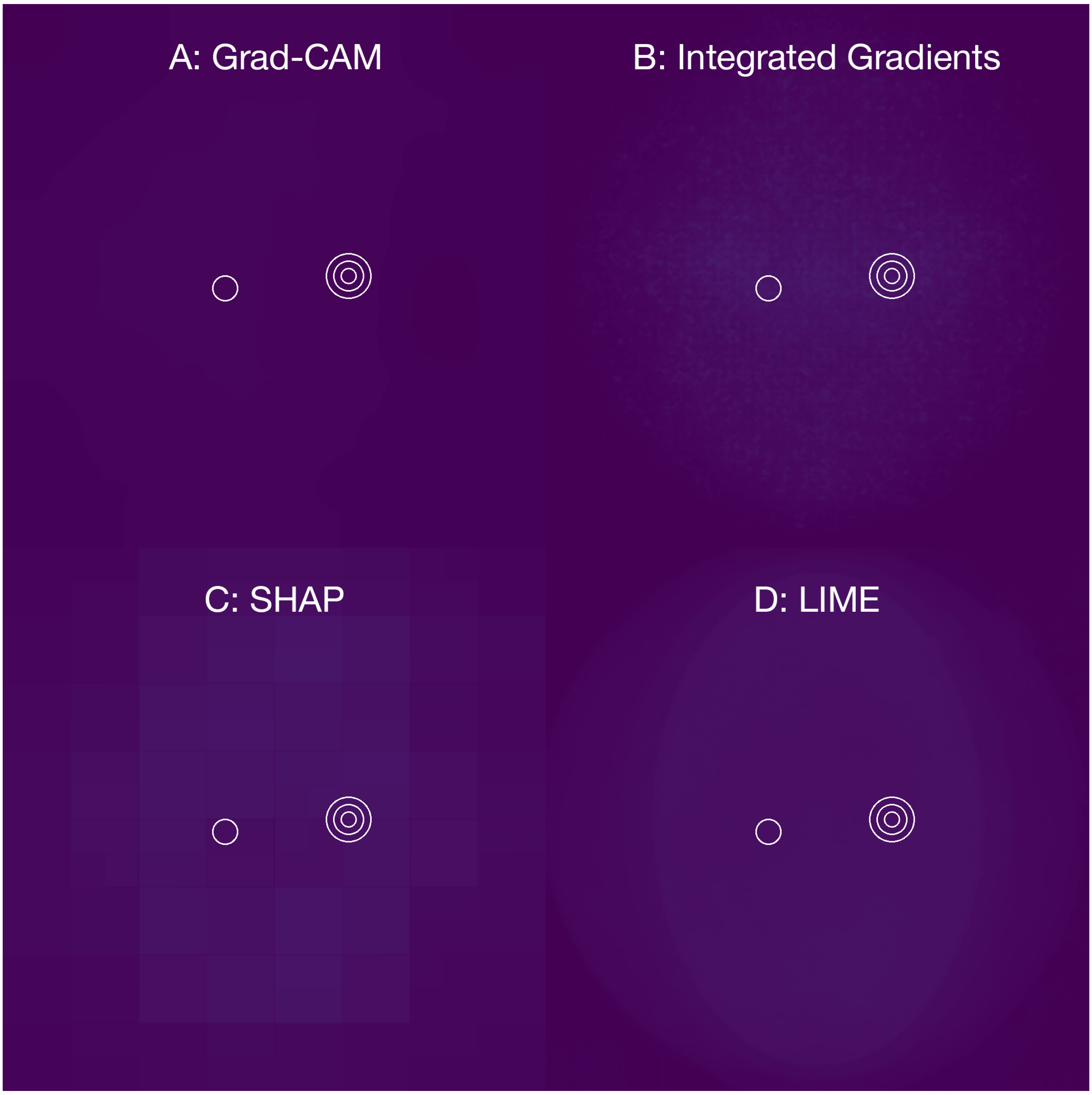

Figure 43: Standard deviation visualisation of “refer” model heatmaps after bootstrapping with 10,000 iterations for Grad-CAM (A), Integrated Gradients (B), SHAP (C) and LIME (D). Values are displayed relative to the maximum pixel intensity.

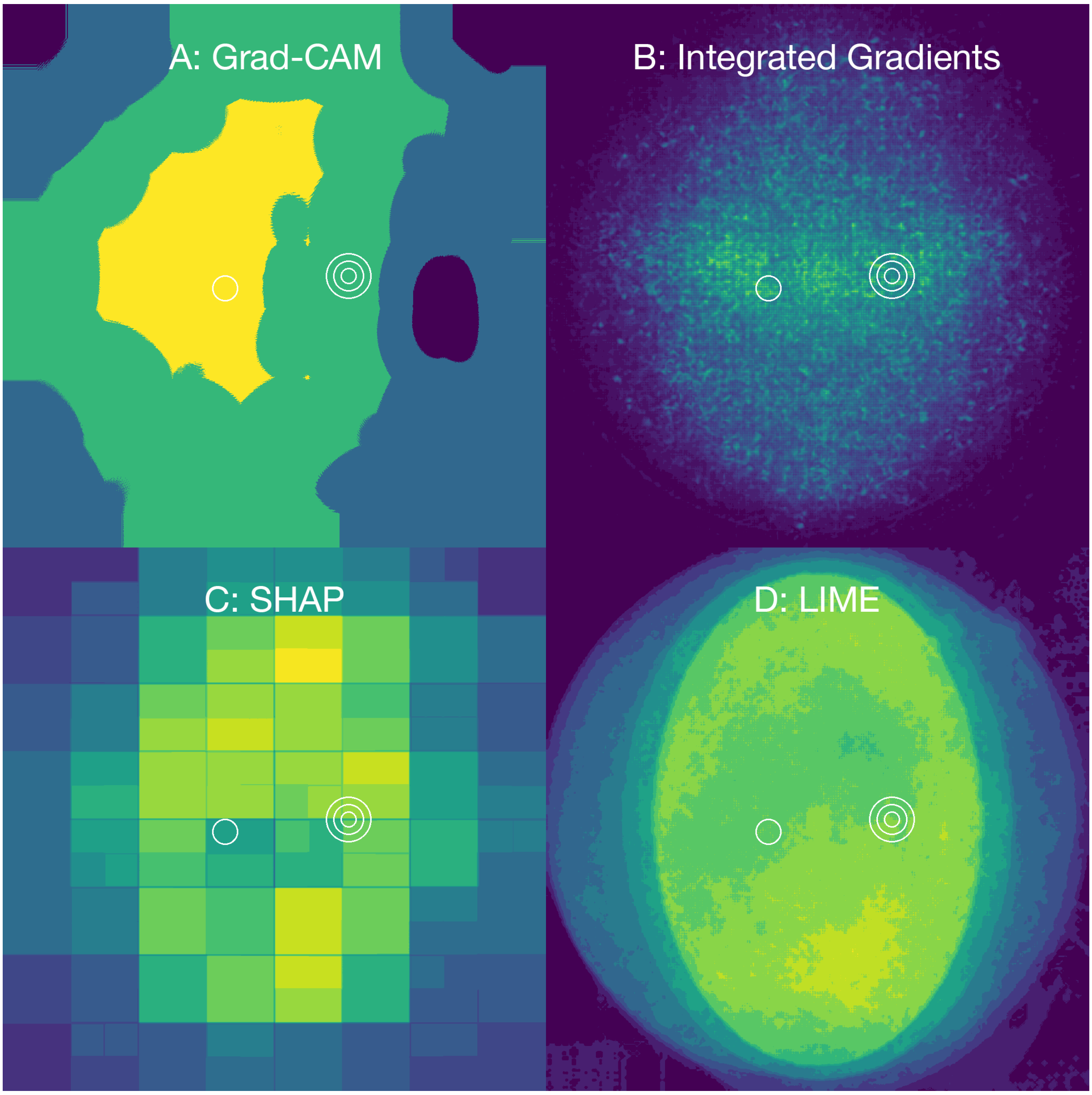

Figure 44: Standard deviation visualisation of "refer" model heatmaps after bootstrapping with 10,000 iterations for Grad-CAM (A), Integrated Gradients (B), SHAP (C) and LIME (D). Values have been normalised for clarity.

### 2 Grade models

#### 2.1 No gradeable diabetic retinopathy predicted

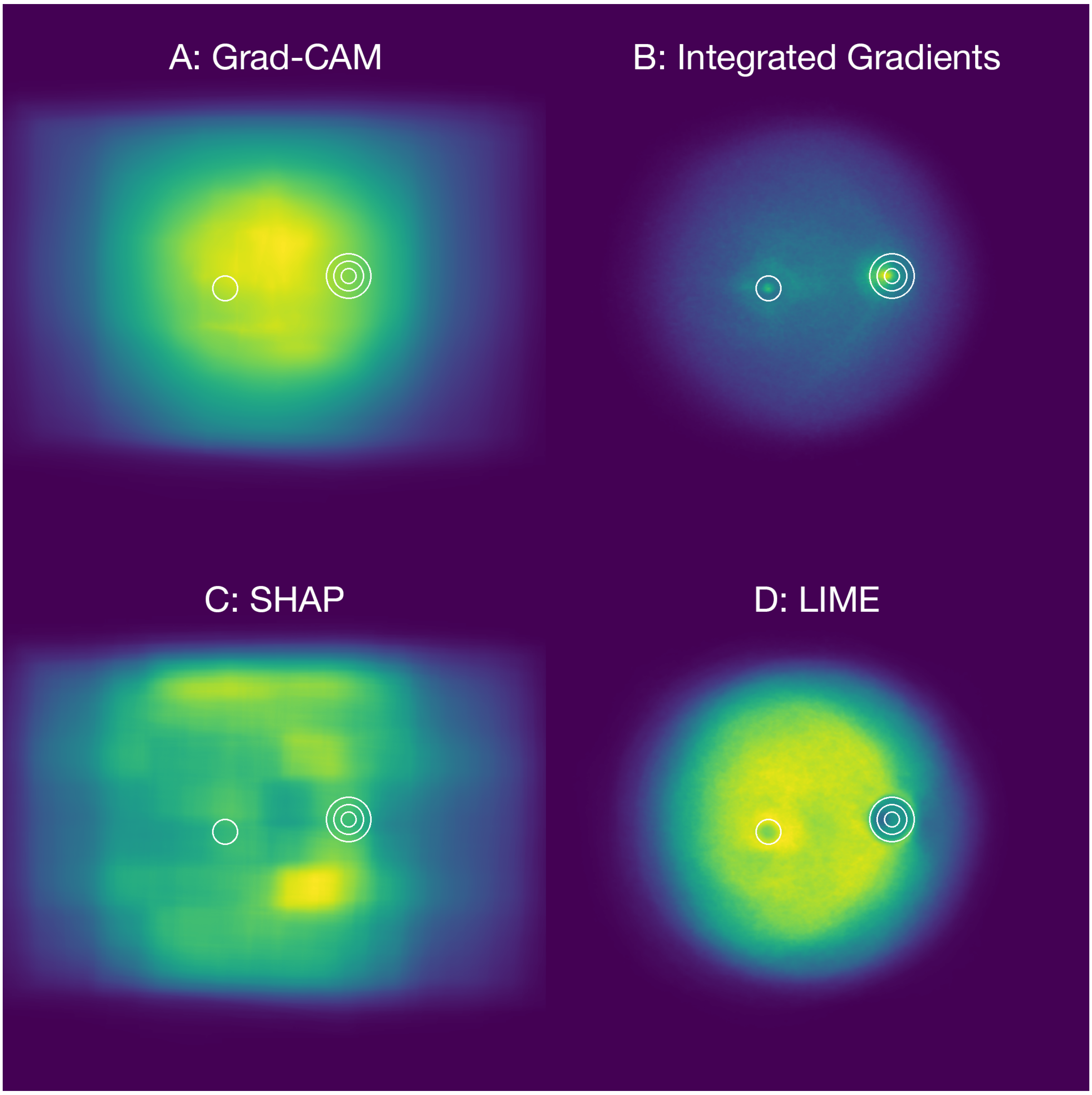

Figure 45: Aggregated "grade" model heatmaps for images predicted as not having gradeable diabetic retinopathy using Grad-CAM (A), Integrated Gradients (B), SHAP (C) and LIME (D).

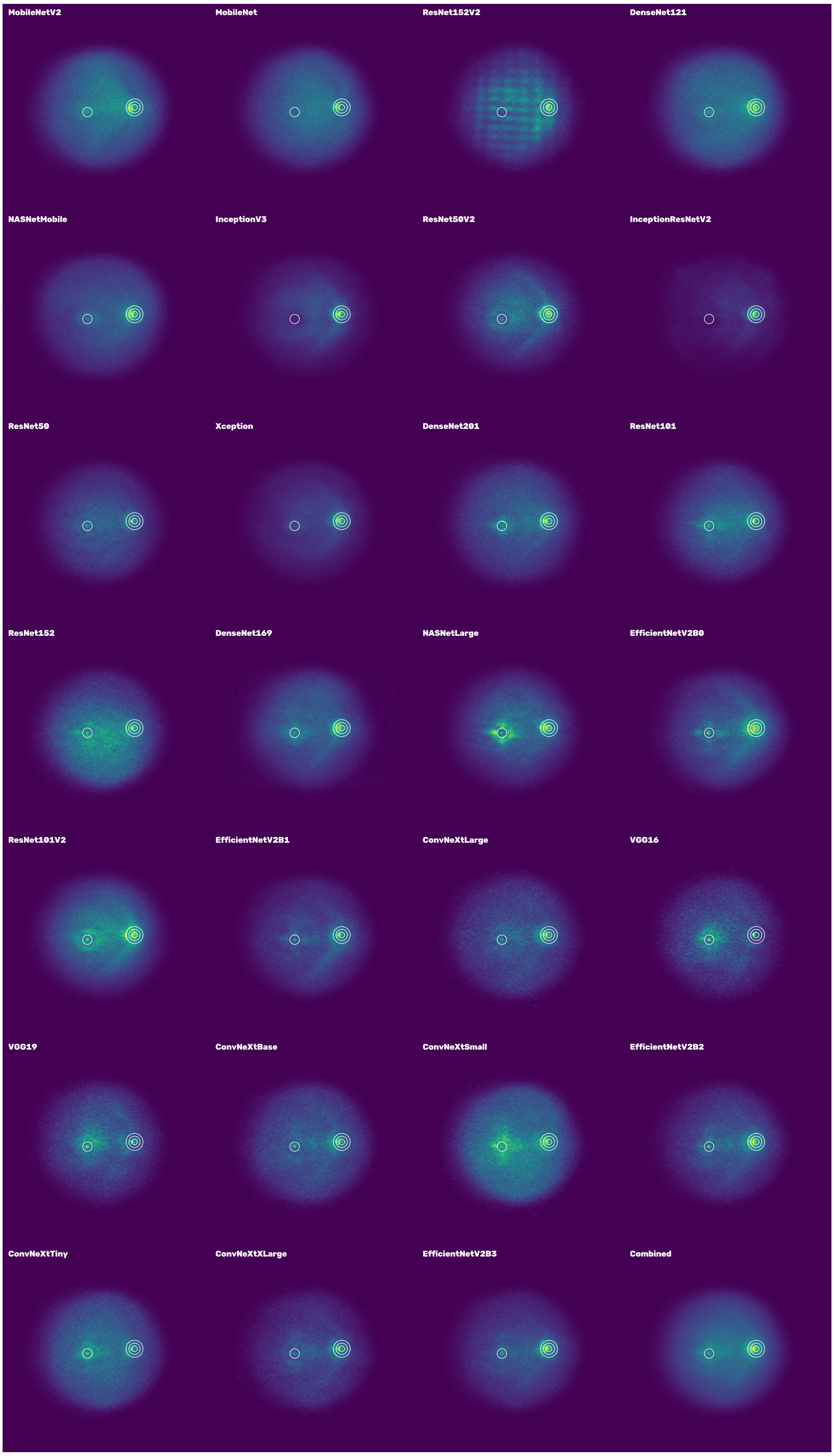

Figure 46: Collated Integrated Gradient heatmaps for "grade" models for images predicted as not having gradeable diabetic retinopathy. Heatmaps are ordered by Macro-F1 score in ascending order.

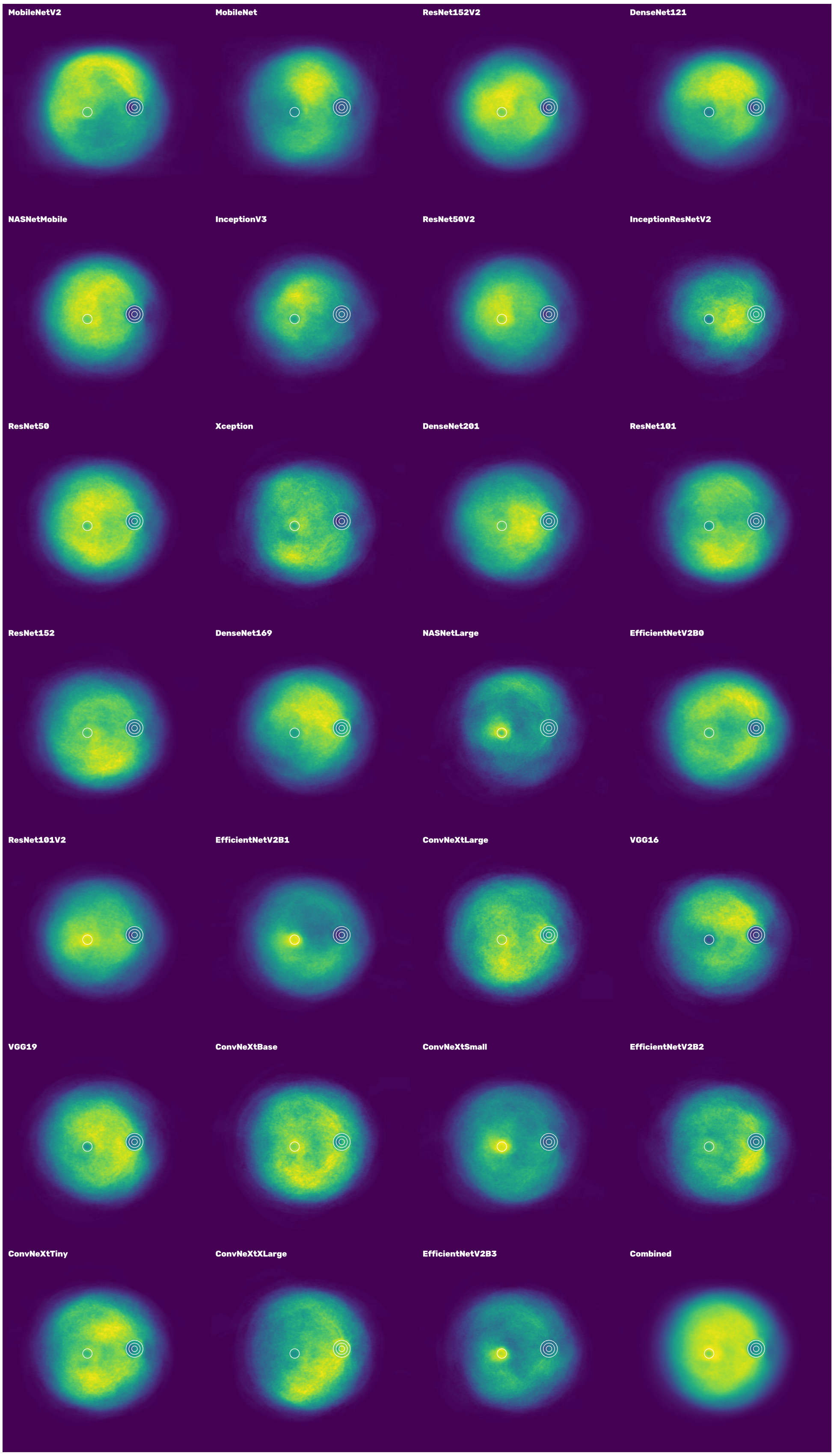

Figure 47: Collated LIME heatmaps for "grade" models for images predicted as not having gradeable diabetic retinopathy. Heatmaps are ordered by Macro-F1 score in ascending order.

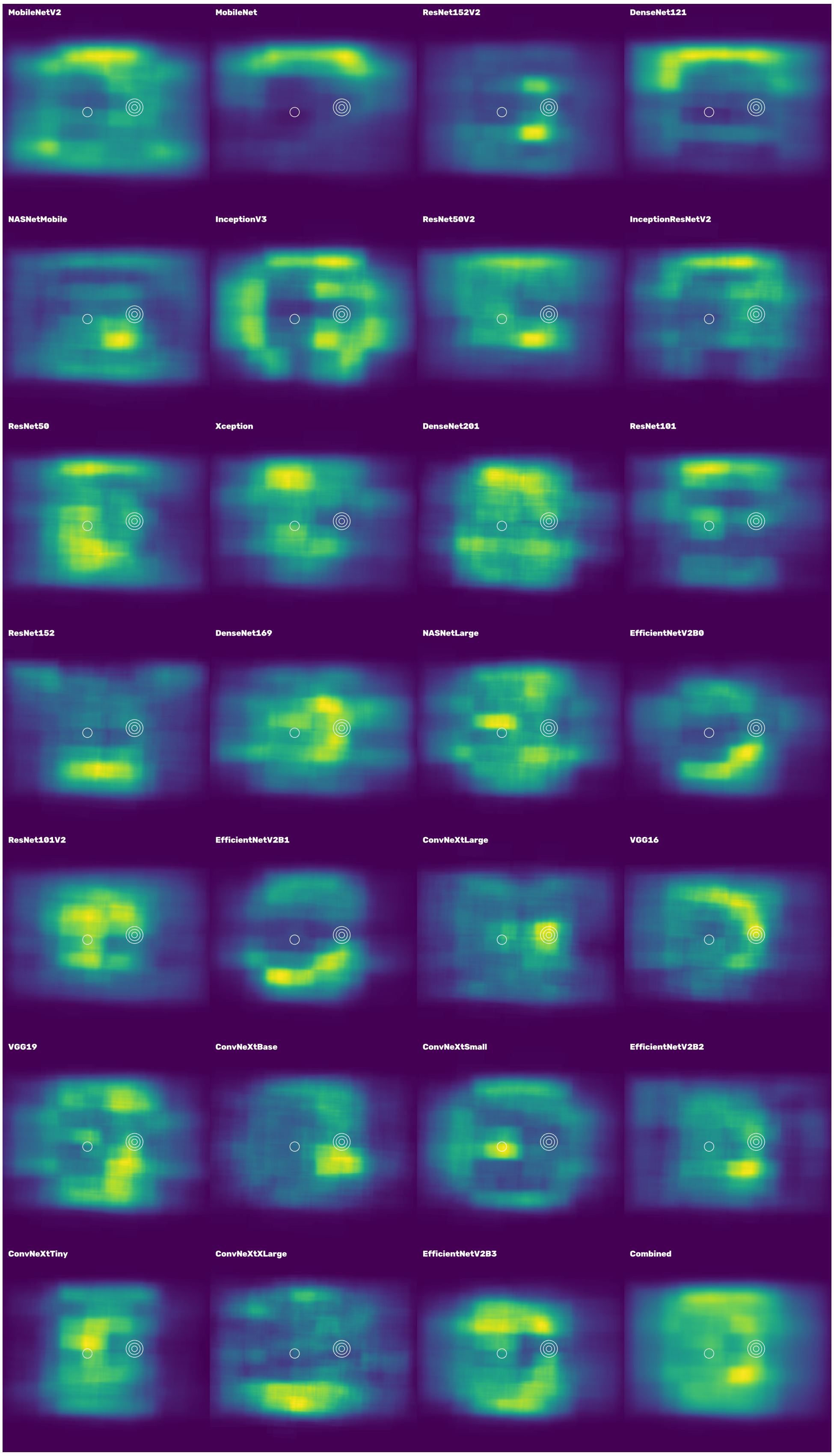

Figure 48: Collated SHAP heatmaps for "grade" models for images predicted as not having gradeable diabetic retinopathy. Heatmaps are ordered by Macro-F1 score in ascending order.

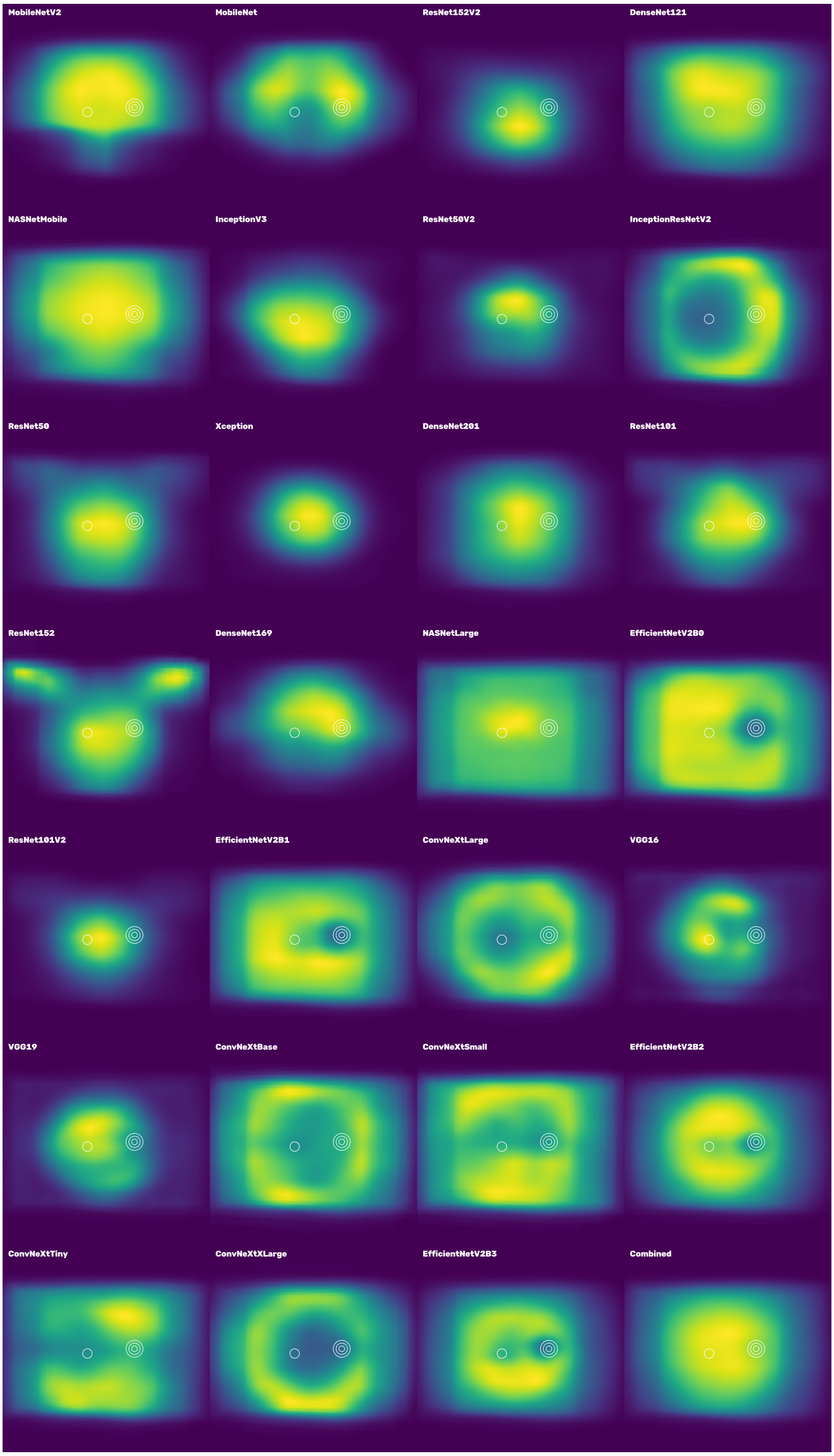

Figure 49: Collated Grad-CAM heatmaps for "grade" models for images predicted as not having gradeable diabetic retinopathy. Heatmaps are ordered by Macro-F1 score in ascending order.

#### 2.2 Referable diabetic retinopathy predicted

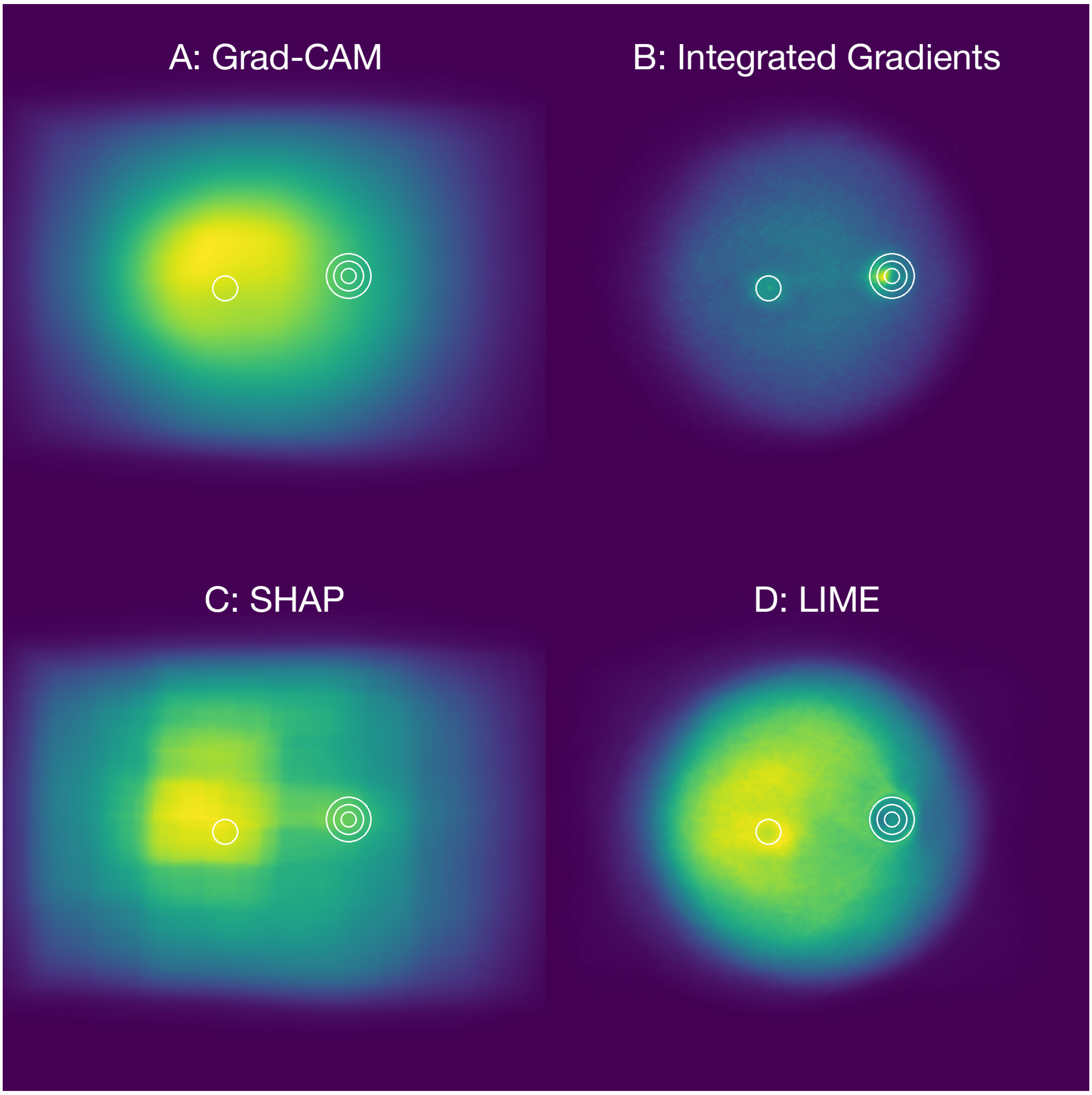

Figure 50: Aggregated "grade" model heatmaps for images predicted as having gradeable diabetic retinopathy using Grad-CAM (A), Integrated Gradients (B), SHAP (C) and LIME (D).

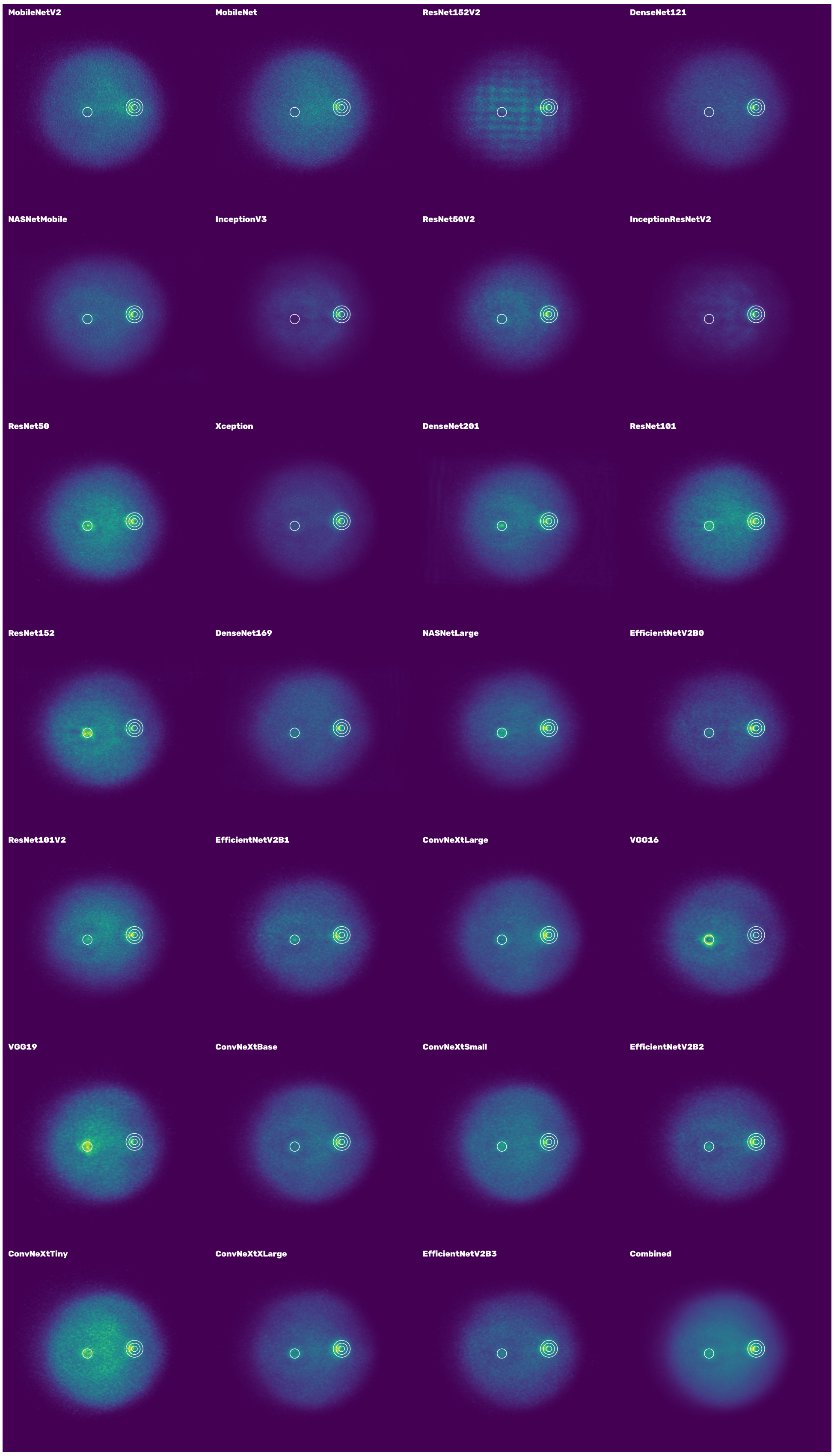

Figure 51: Collated Integrated Gradient heatmaps for "grade" models for images predicted as having gradeable diabetic retinopathy. Heatmaps are ordered by Macro-F1 score in ascending order.

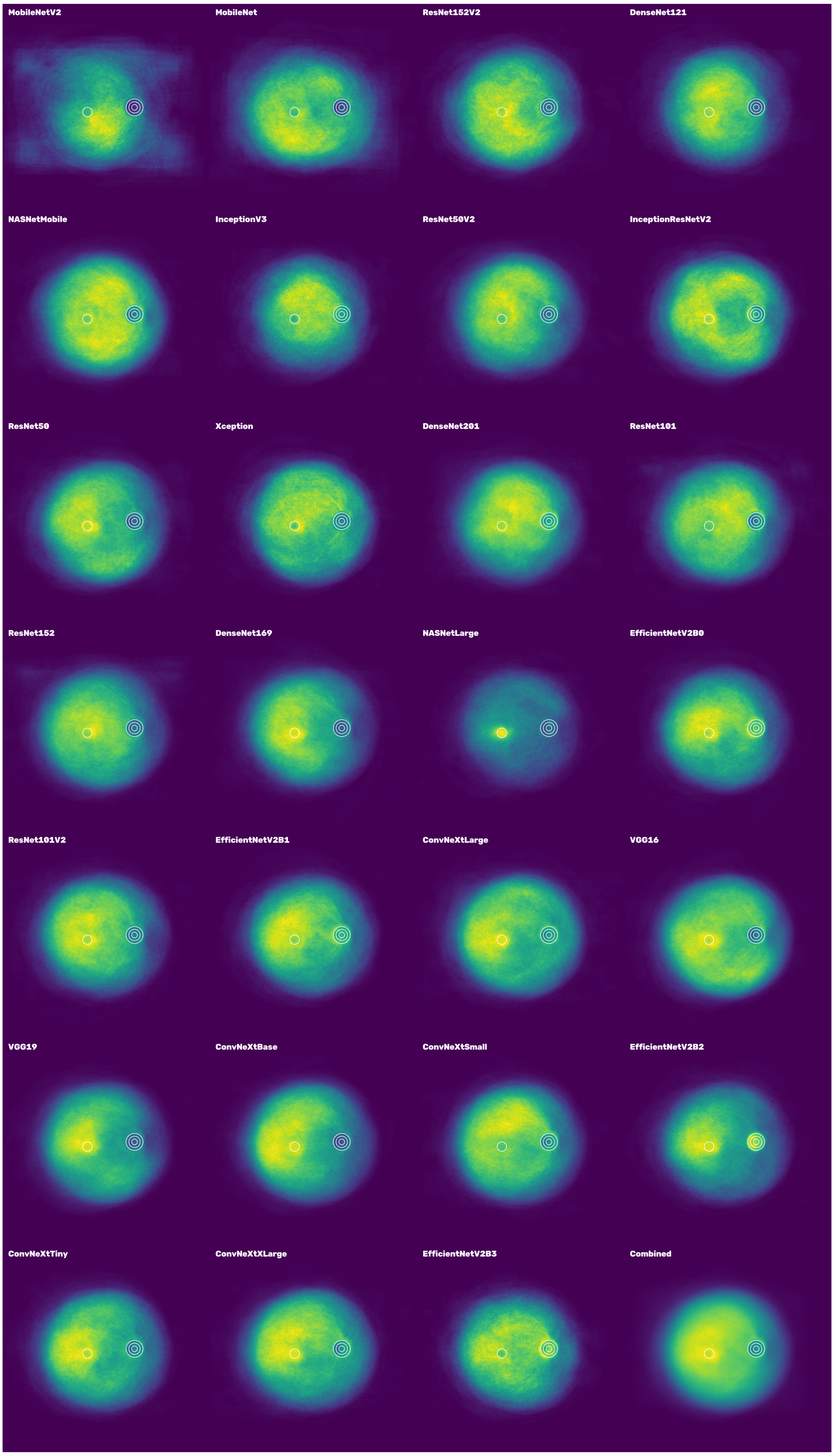

Figure 52: Collated LIME heatmaps for "grade" models for images predicted as having gradeable diabetic retinopathy. Heatmaps are ordered by Macro-F1 score in ascending order.

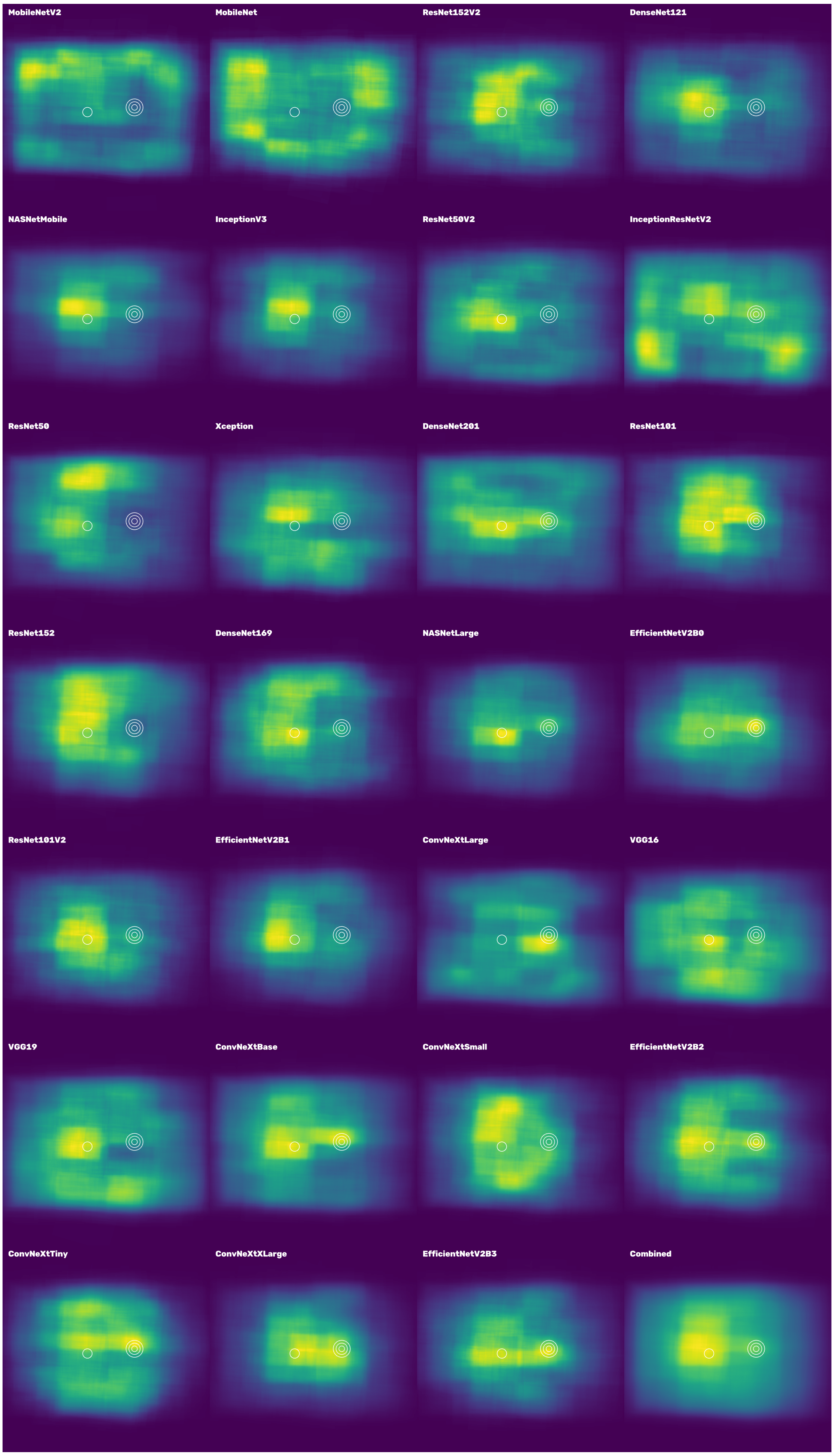

Figure 53: Collated SHAP heatmaps for "grade" models for images predicted as having gradeable diabetic retinopathy. Heatmaps are ordered by Macro-F1 score in ascending order.

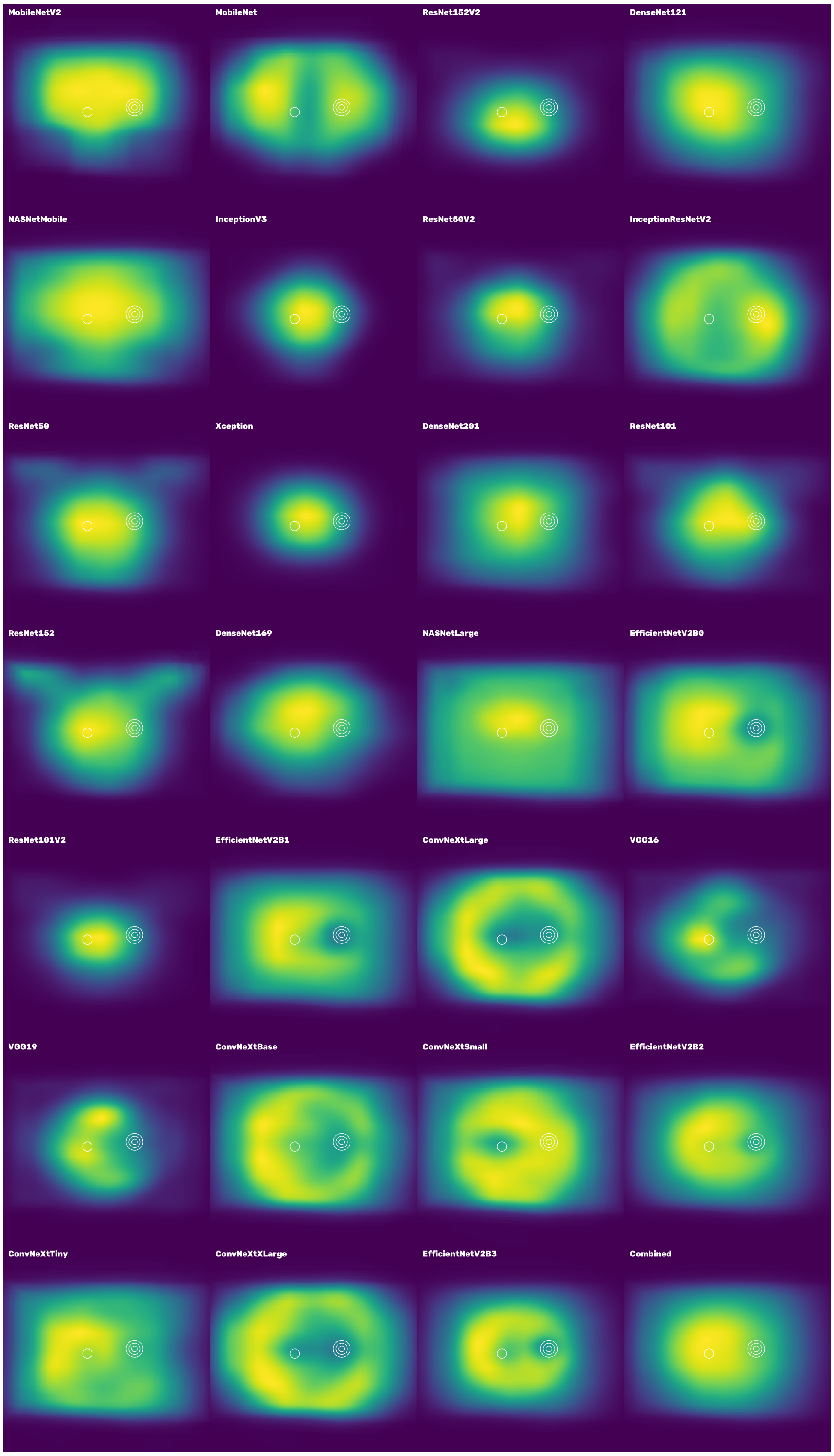

Figure 54: Collated Grad-CAM heatmaps for "grade" models for images predicted as having gradeable diabetic retinopathy. Heatmaps are ordered by Macro-F1 score in ascending order.

#### 2.3 Diabetic retinopathy feature coverage

##### 2.3.1 Grad-CAM

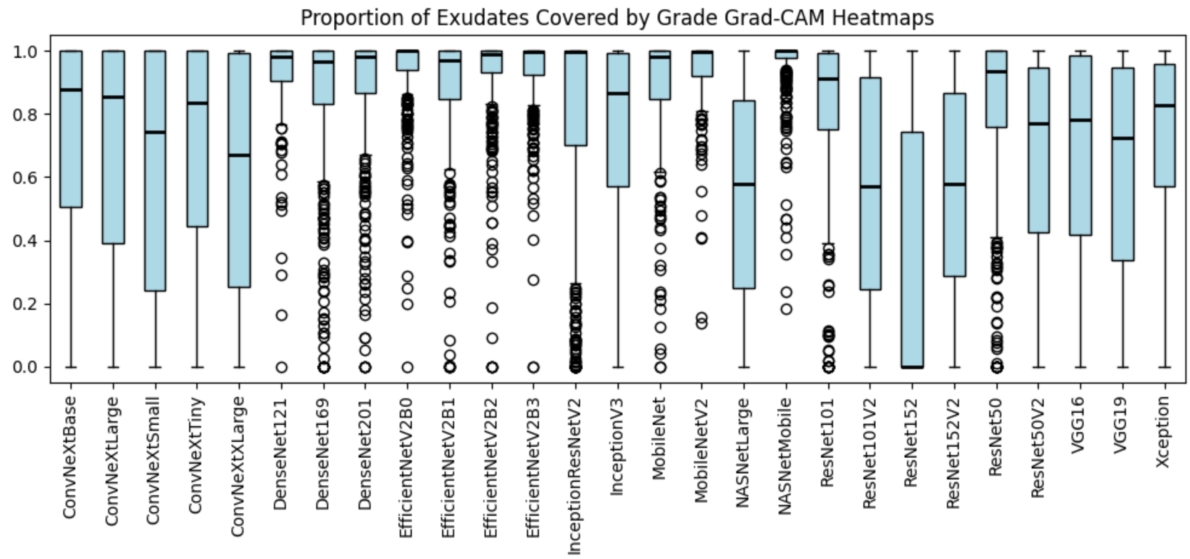

Figure 55: Proportion of exudates covered by “grade” Grad-CAM heatmaps.

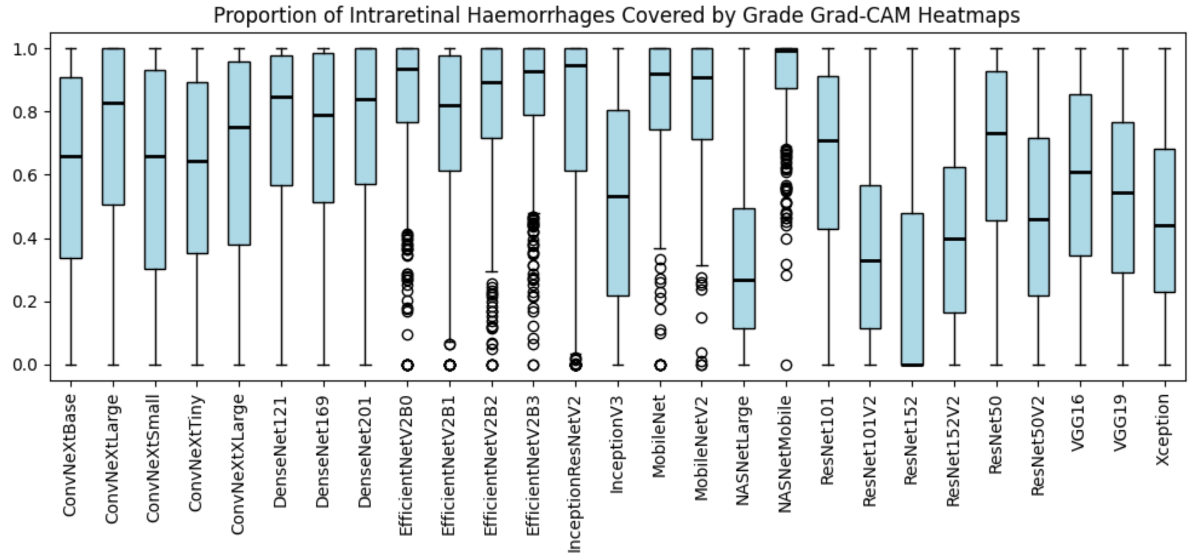

Figure 56: Proportion of intraretinal haemorrhages covered by “grade” Grad-CAM heatmaps.

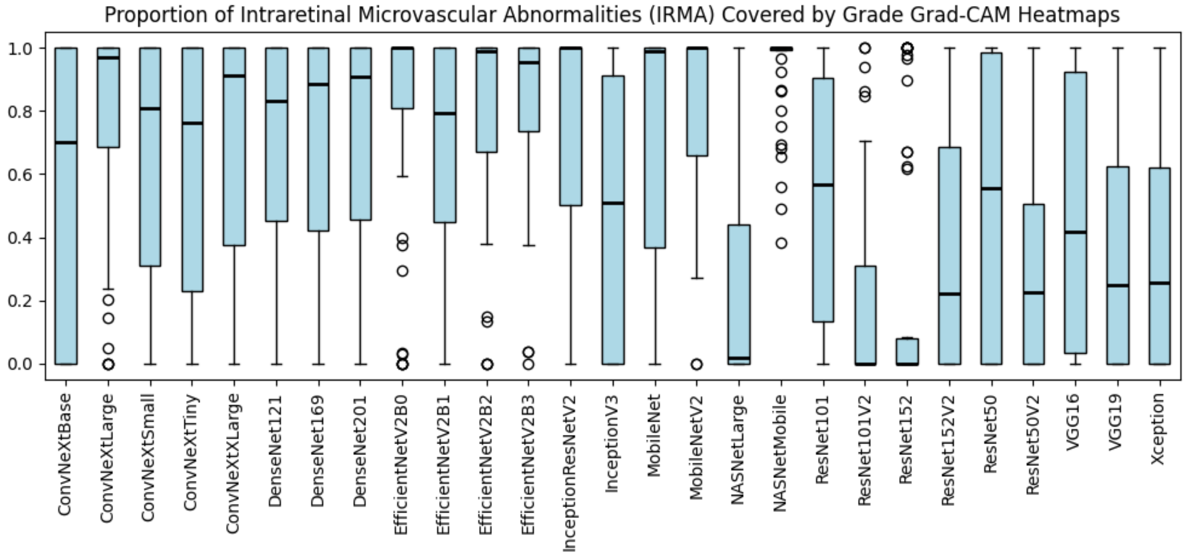

Figure 57: Proportion of intraretinal microvascular abnormalities (IRMA) covered by "grade" Grad-CAM heatmaps.

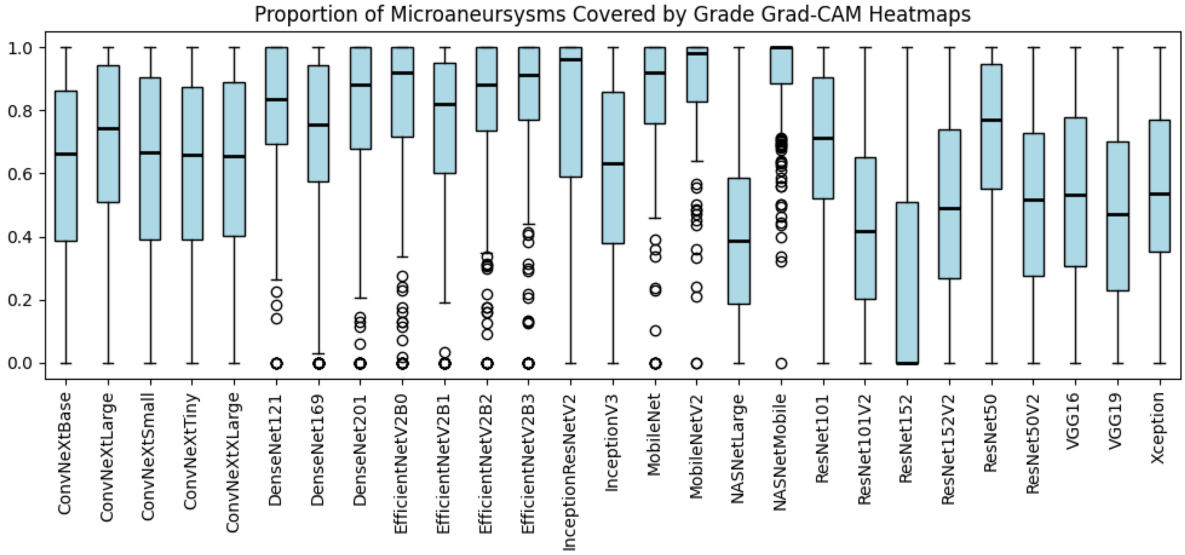

Figure 58: Proportion of microaneurysms covered by "grade" Grad-CAM heatmaps.

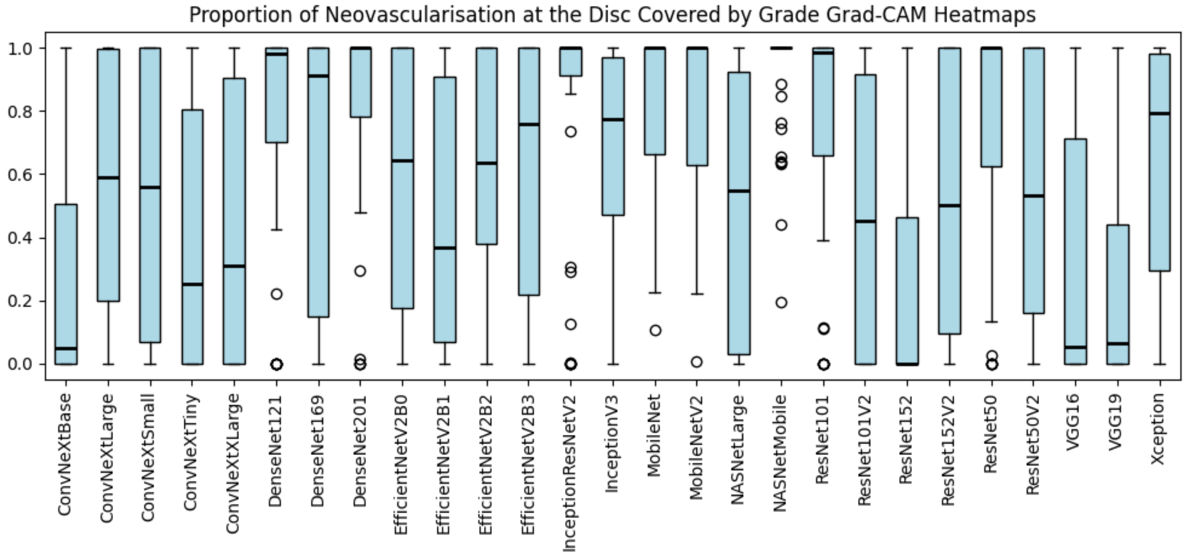

Figure 59: Proportion of neovascularisation at the disc covered by “grade” Grad-CAM heatmaps.

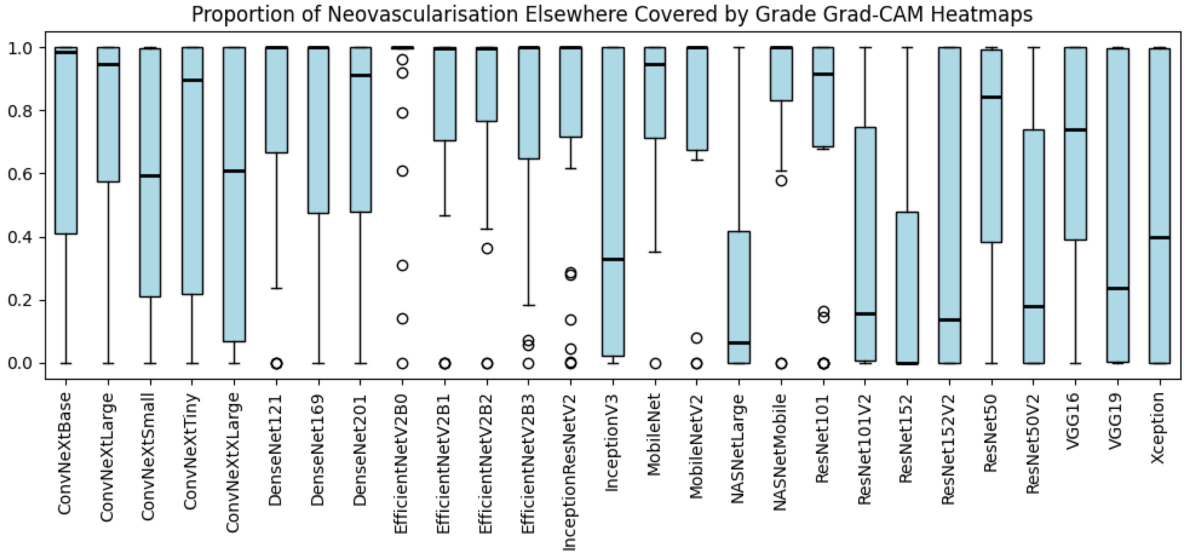

Figure 60: Proportion of neovascularisation elsewhere covered by “grade” Grad-CAM heatmaps.

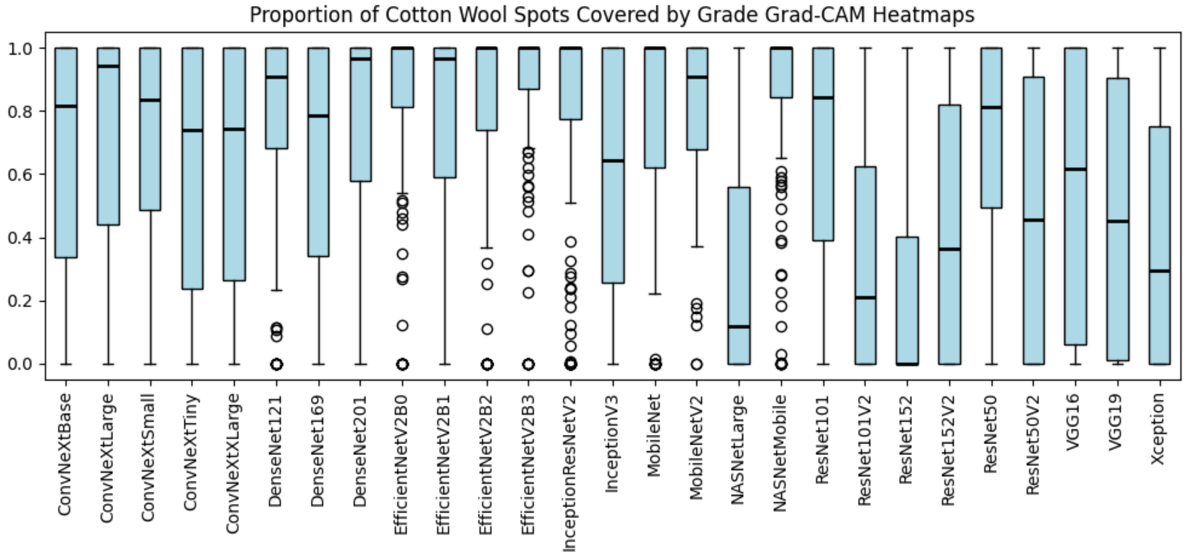

Figure 61: Proportion of cotton wool spots covered by “grade” Grad-CAM heatmaps.

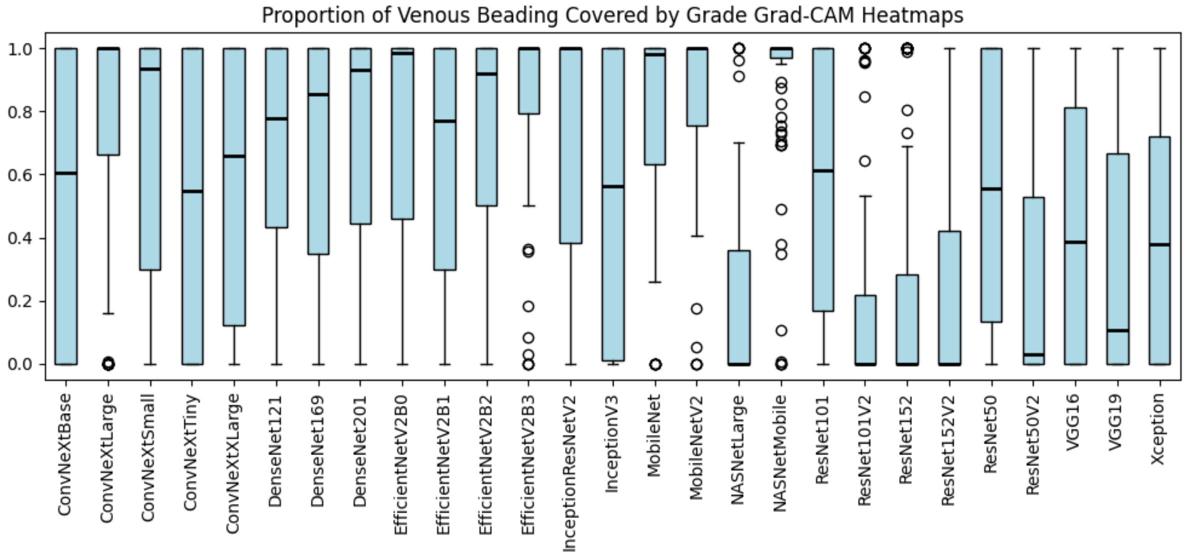

Figure 62: Proportion of venous beading covered by “grade” Grad-CAM heatmaps.

##### 2.3.2 Integrated Gradients

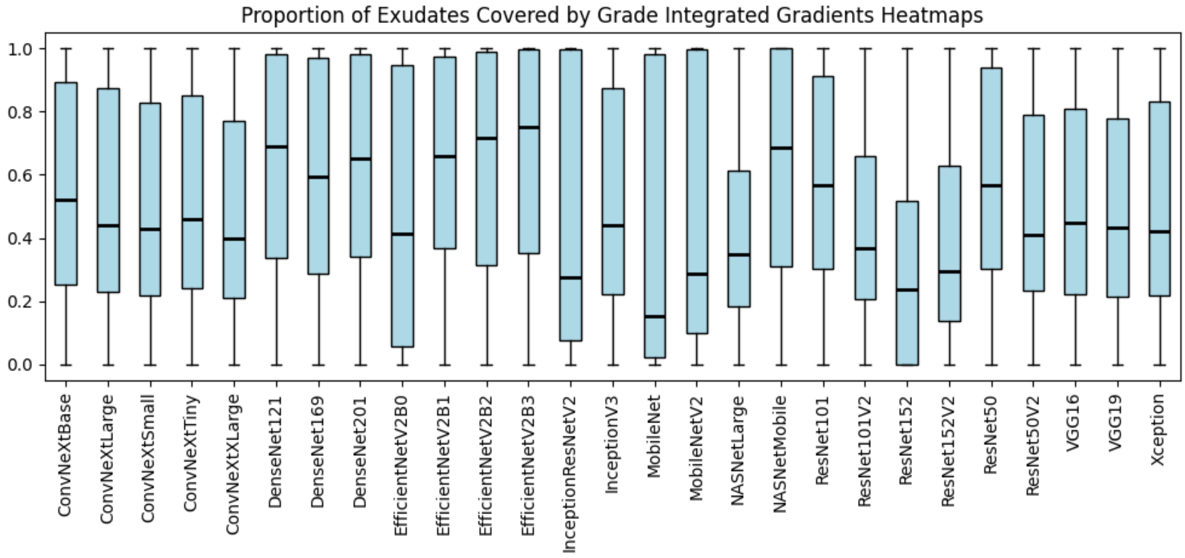

Figure 63: Proportion of exudates covered by “grade” Integrated Gradient heatmaps.

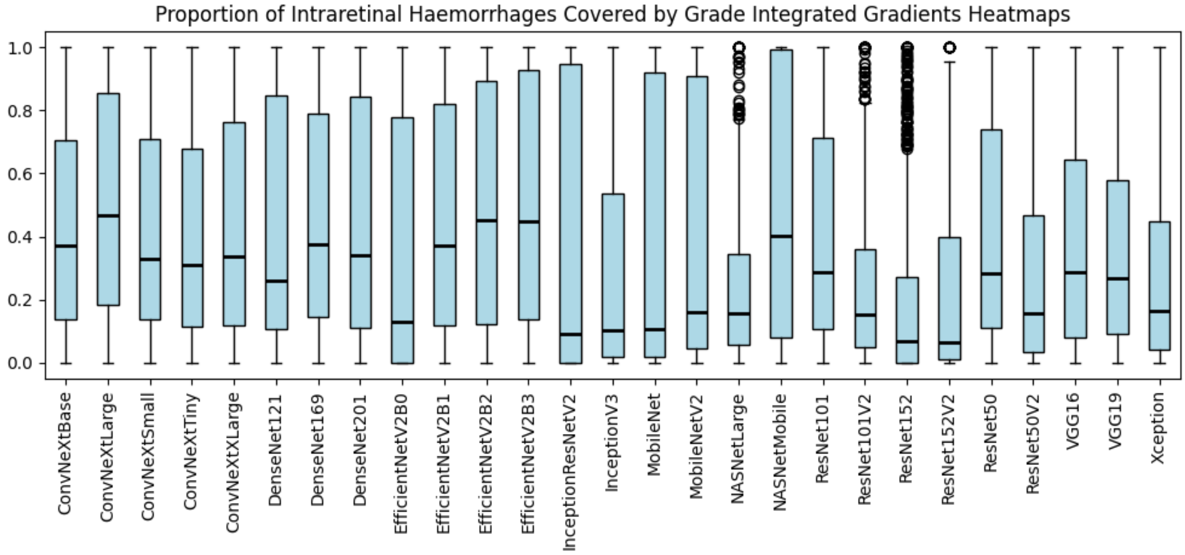

Figure 64: Proportion of intraretinal haemorrhages covered by “grade” Integrated Gradient heatmaps.

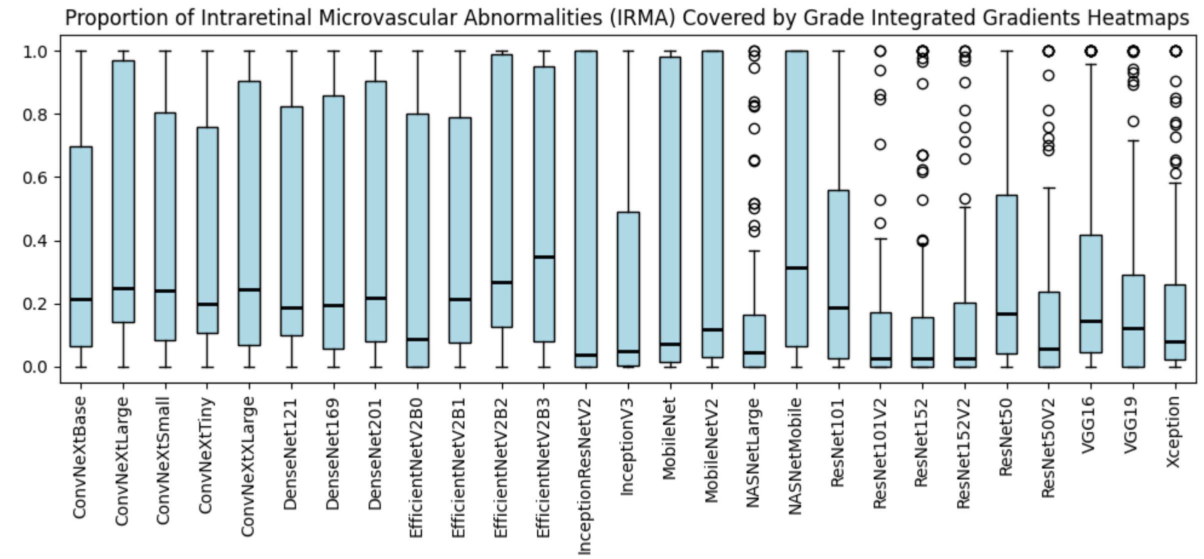

Figure 65: Proportion of intraretinal microvascular abnormalities (IRMA) covered by “grade” Integrated Gradient heatmaps.

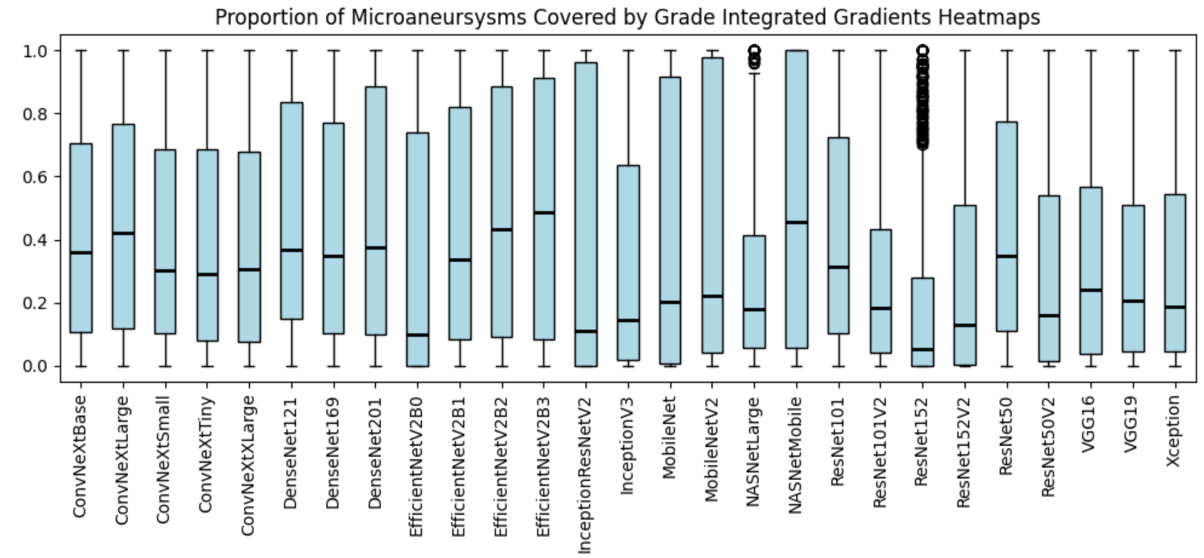

Figure 66: Proportion of microaneurysms covered by “grade” Integrated Gradient heatmaps.

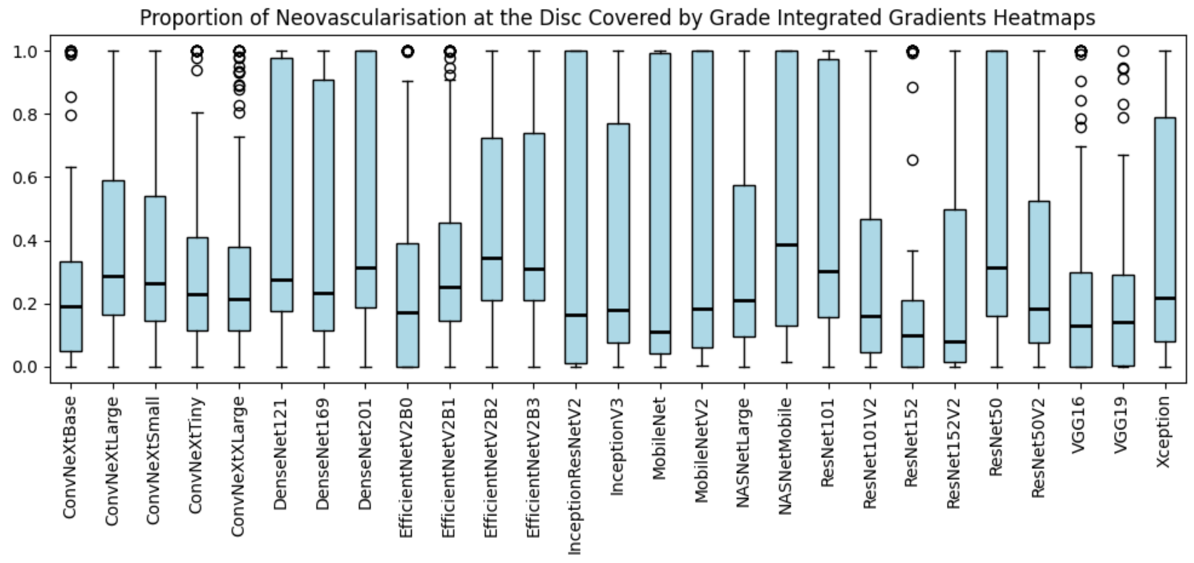

Figure 67: Proportion of neovascularisation at the disc covered by “grade” Integrated Gradient heatmaps.

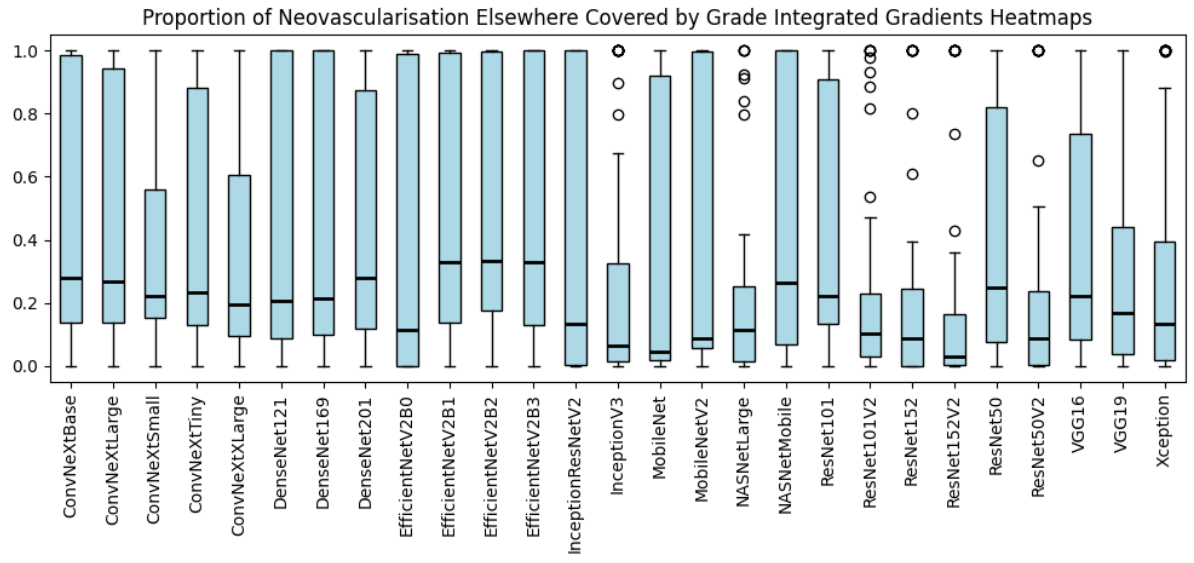

Figure 68: Proportion of neovascularisation elsewhere covered by “grade” Integrated Gradient heatmaps.

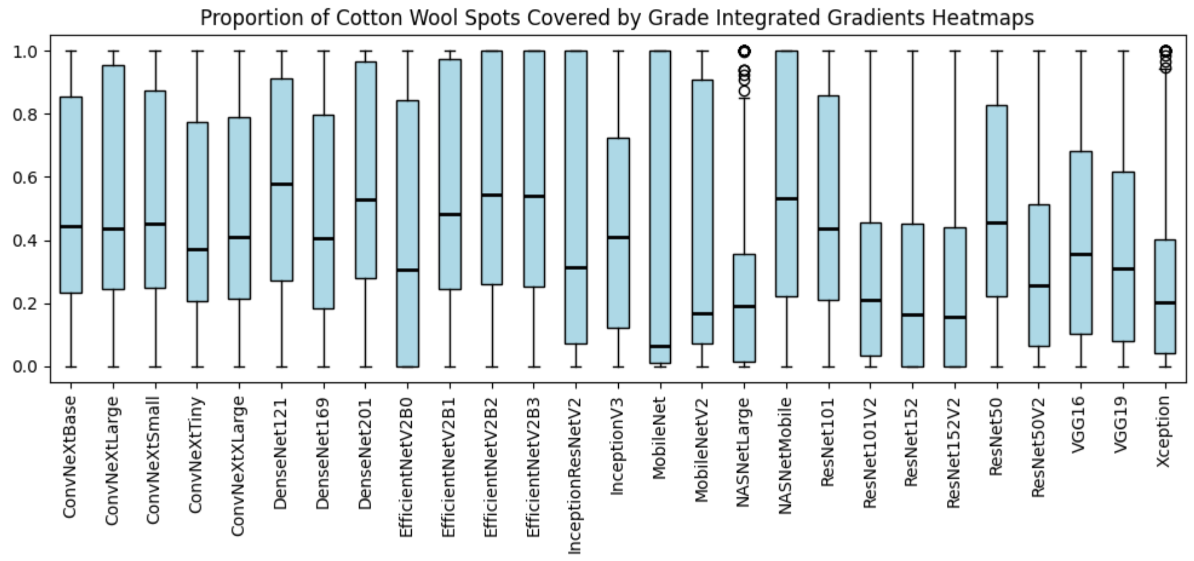

Figure 69: Proportion of cotton wool spots covered by “grade” Integrated Gradient heatmaps.

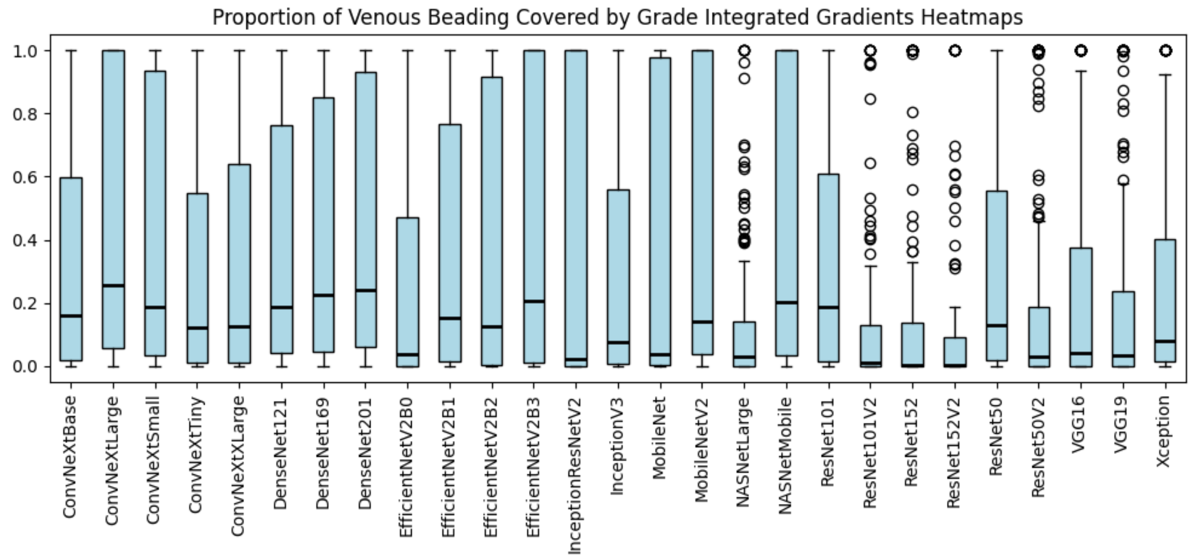

Figure 70: Proportion of venous beading covered by “grade” Integrated Gradient heatmaps.

##### 2.3.3 LIME

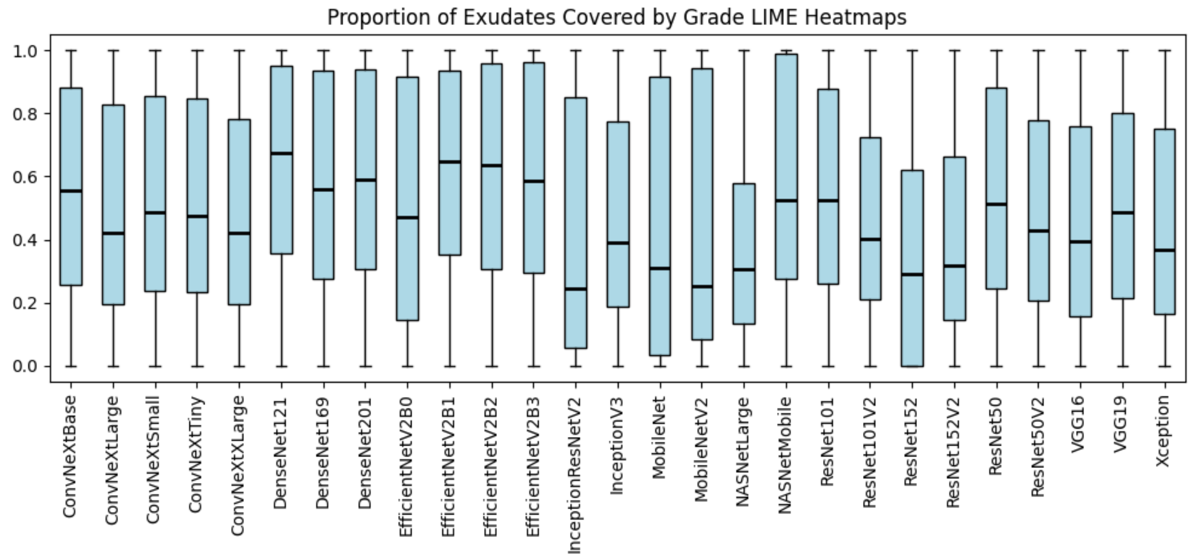

Figure 71: Proportion of exudates covered by “grade” LIME heatmaps.

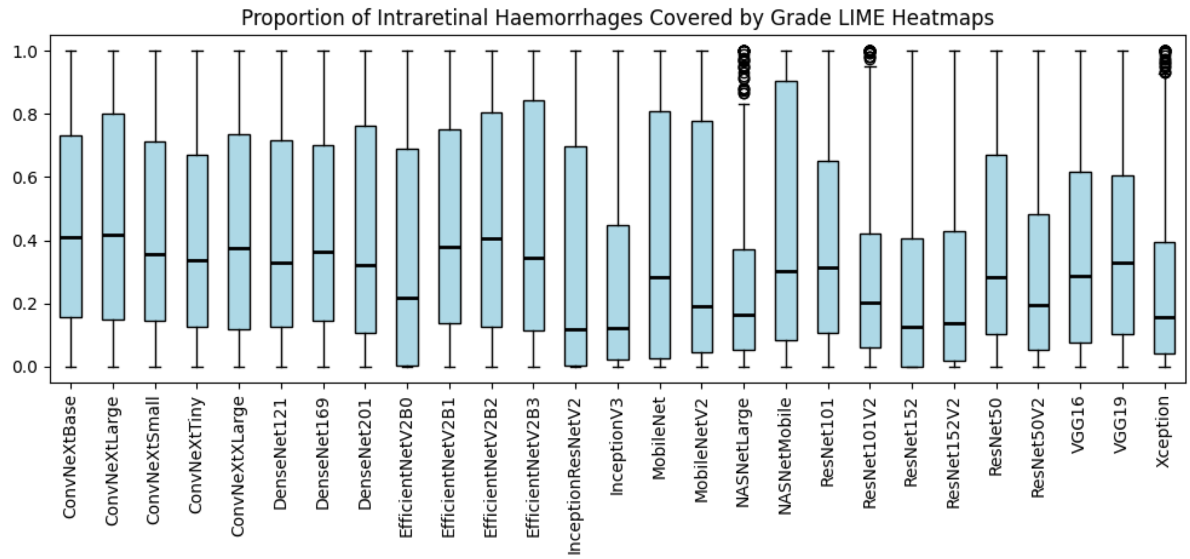

Figure 72: Proportion of intraretinal haemorrhages covered by “grade” LIME heatmaps.

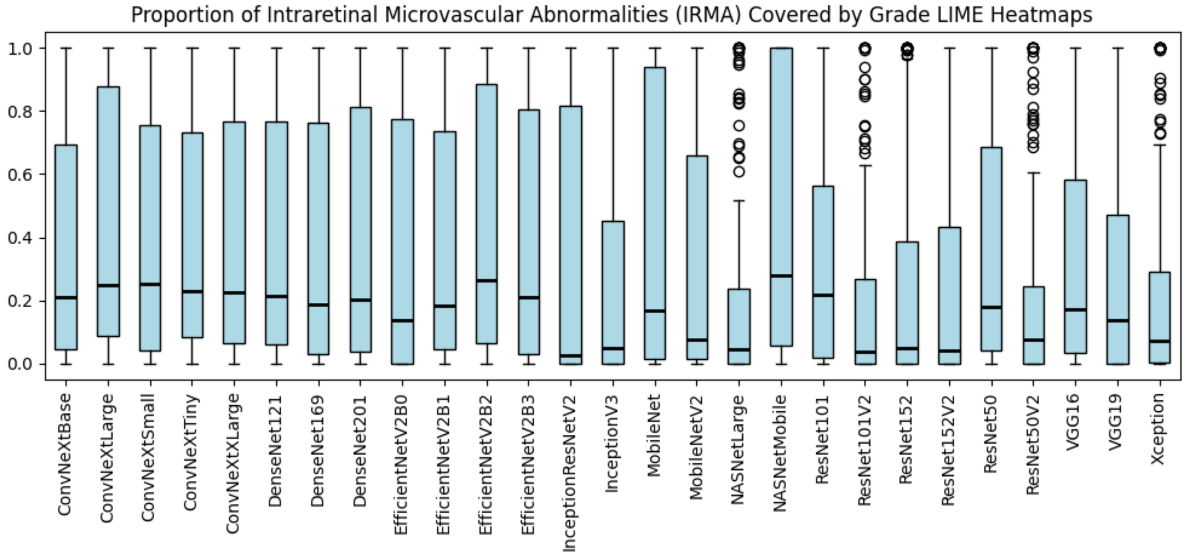

Figure 73: Proportion of intraretinal microvascular abnormalities (IRMA) covered by “grade” LIME heatmaps.

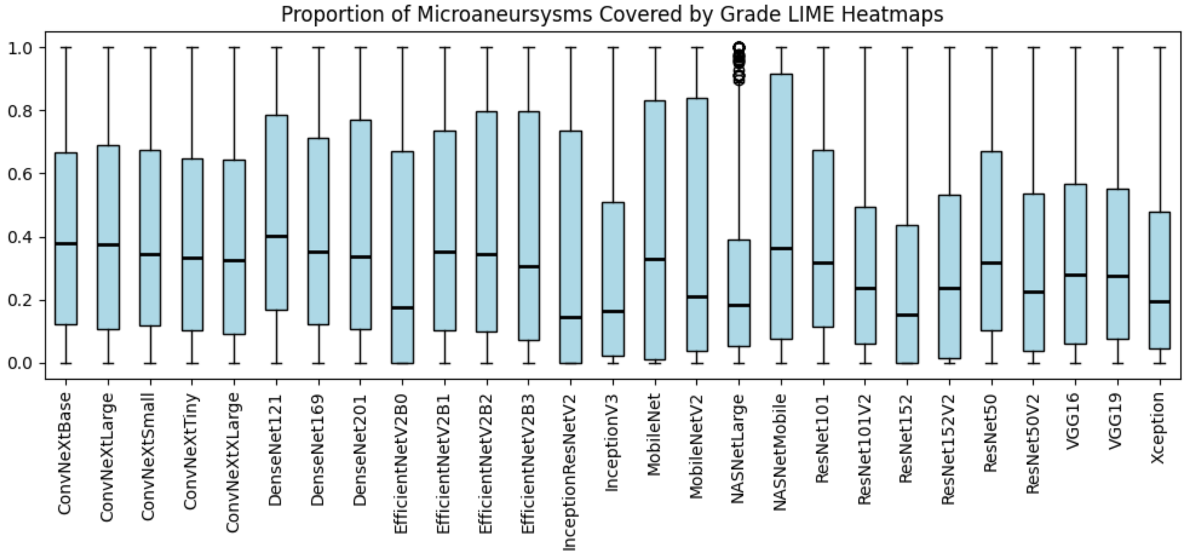

Figure 74: Proportion of microaneurysms covered by “grade” LIME heatmaps.

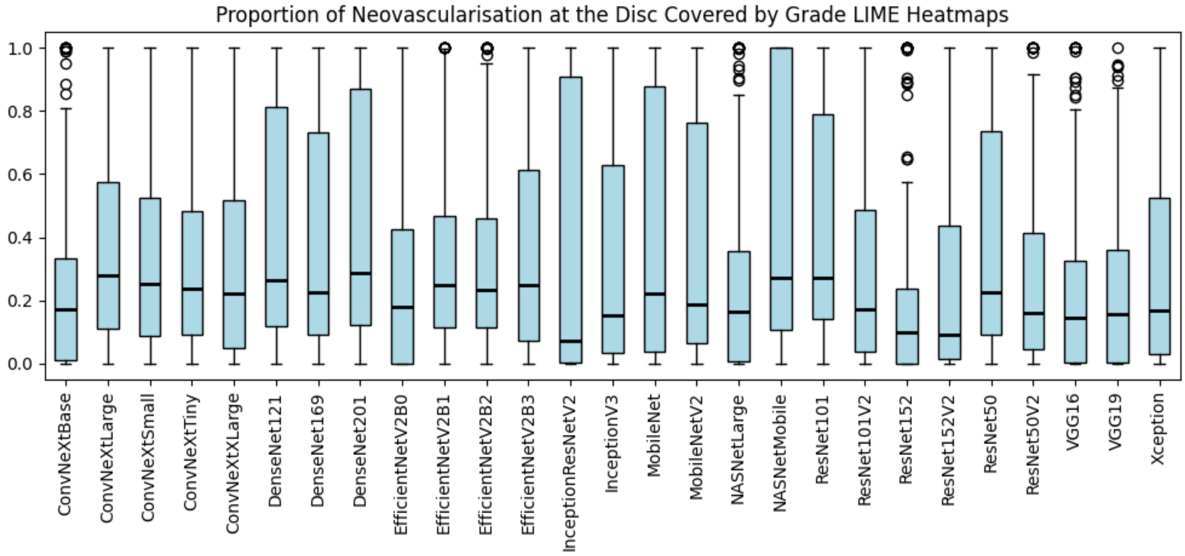

Figure 75: Proportion of neovascularisation at the disc covered by "grade" LIME heatmaps.

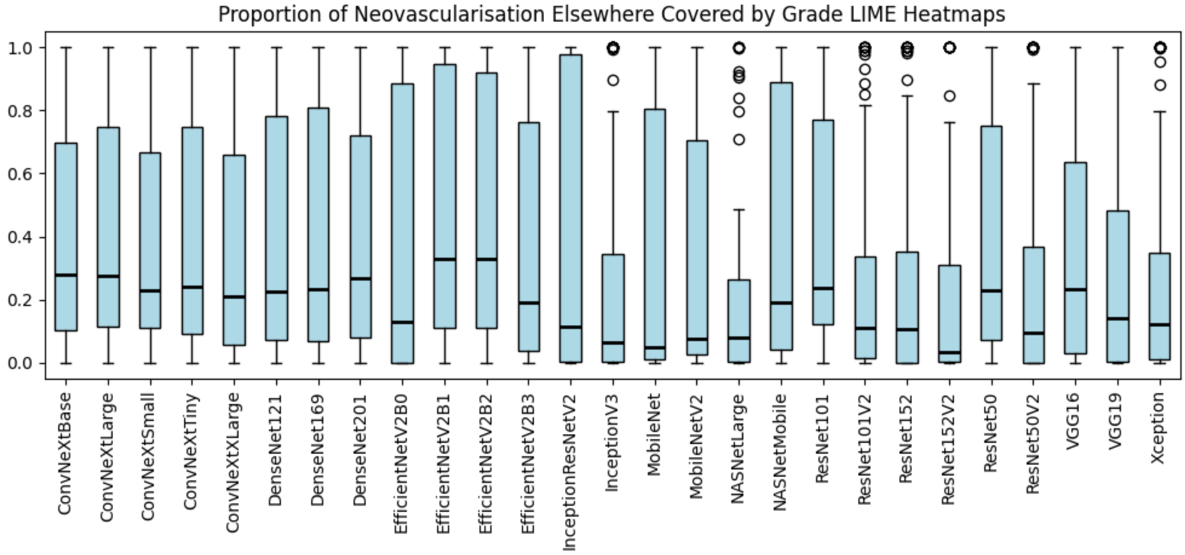

Figure 76: Proportion of neovascularisation elsewhere covered by "grade" LIME heatmaps.

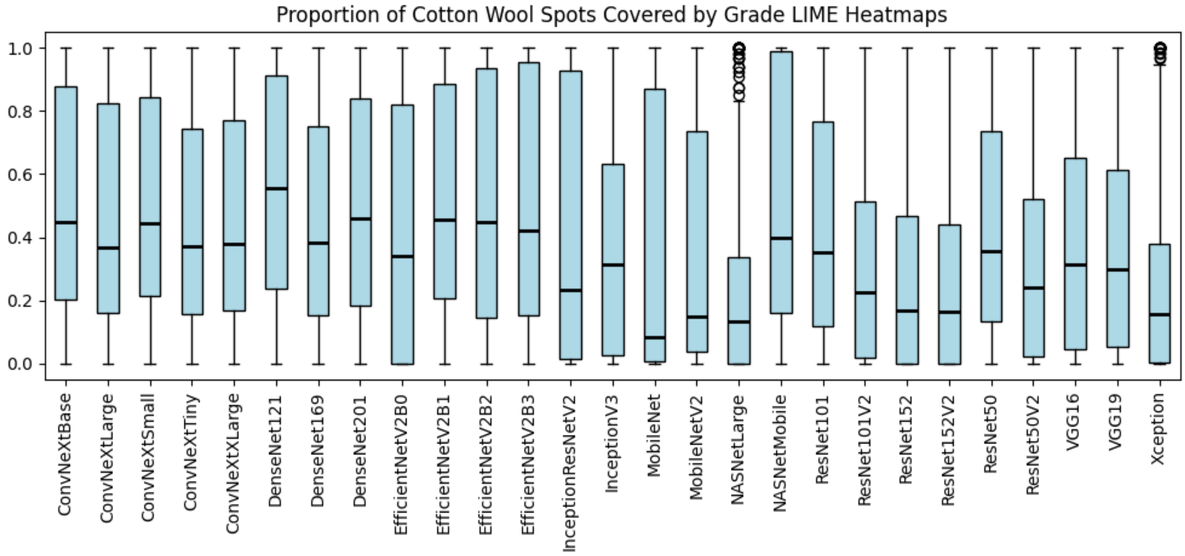

Figure 77: Proportion of cotton wool spots covered by “grade” LIME heatmaps.

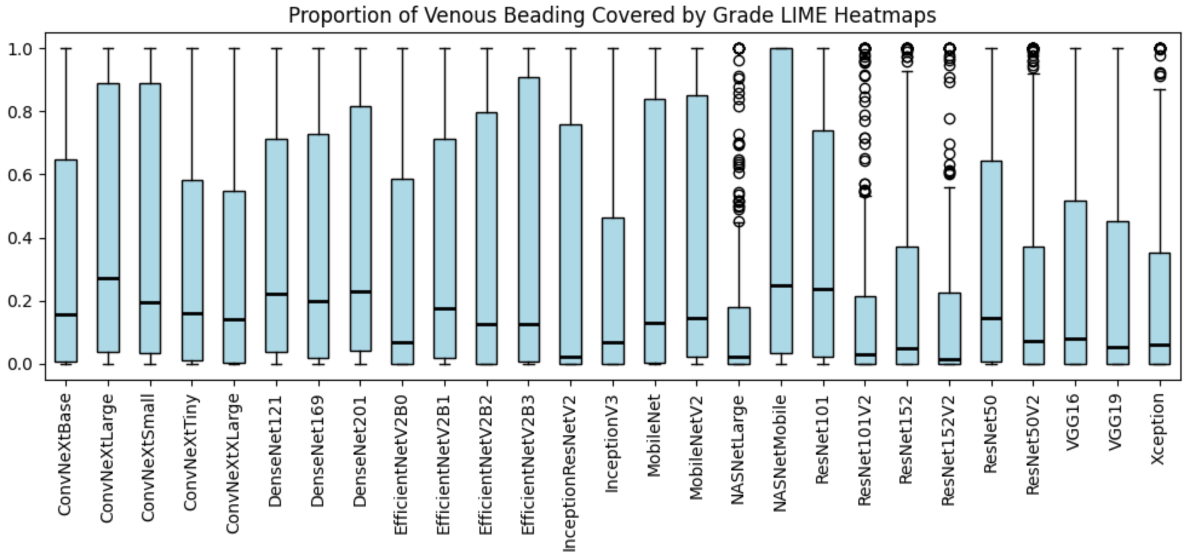

Figure 78: Proportion of venous beading covered by “grade” LIME heatmaps.

##### 2.3.4 SHAP

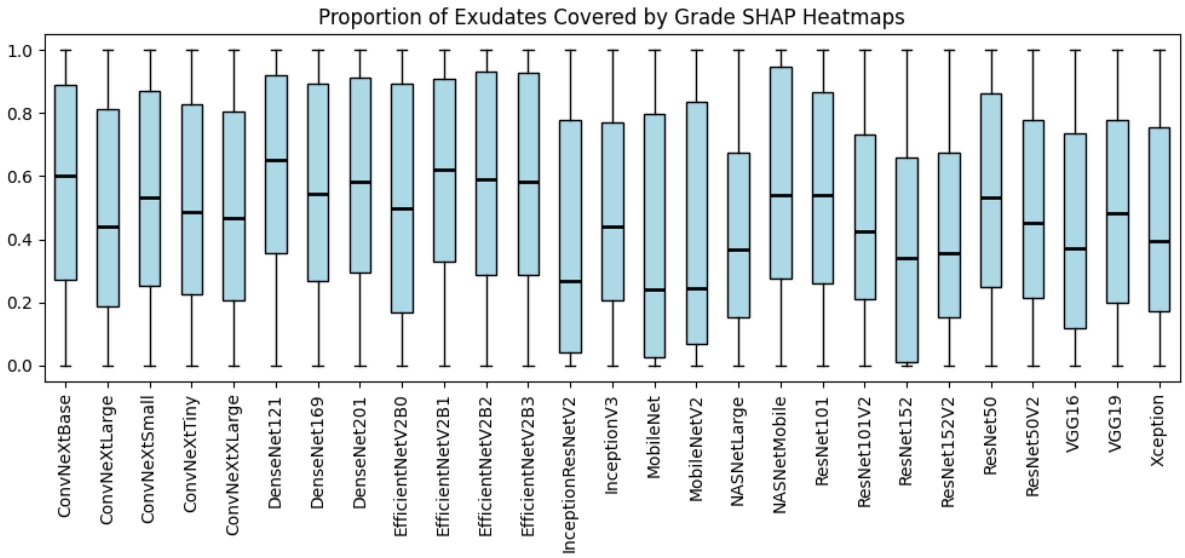

Figure 79: Proportion of exudates covered by "grade" SHAP heatmaps.

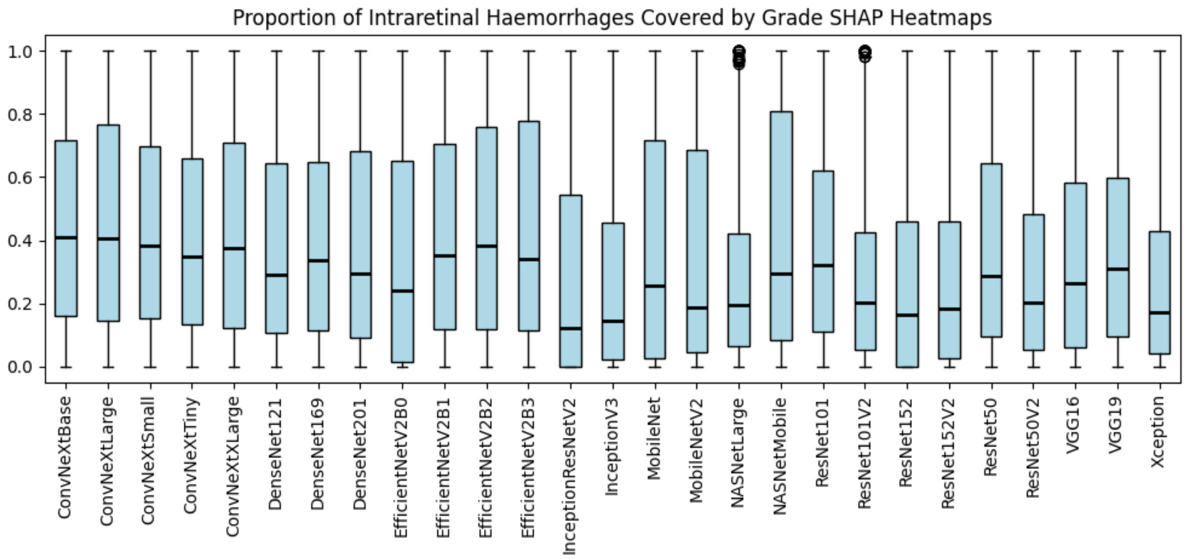

Figure 80: Proportion of intraretinal haemorrhages covered by "grade" SHAP heatmaps.

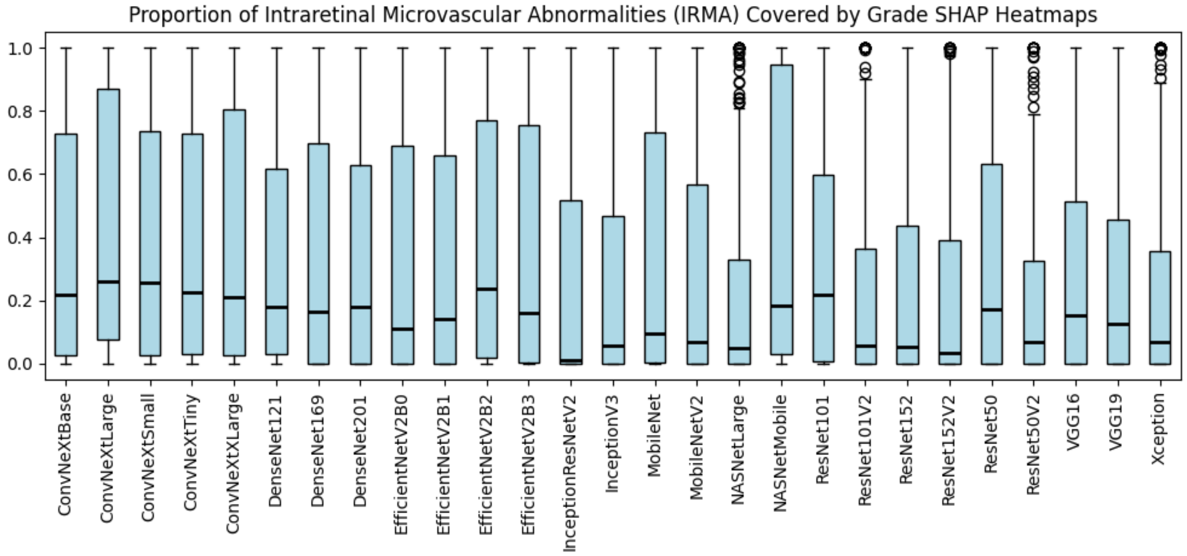

Figure 81: Proportion of intraretinal microvascular abnormalities (IRMA) covered by “grade” SHAP heatmaps.

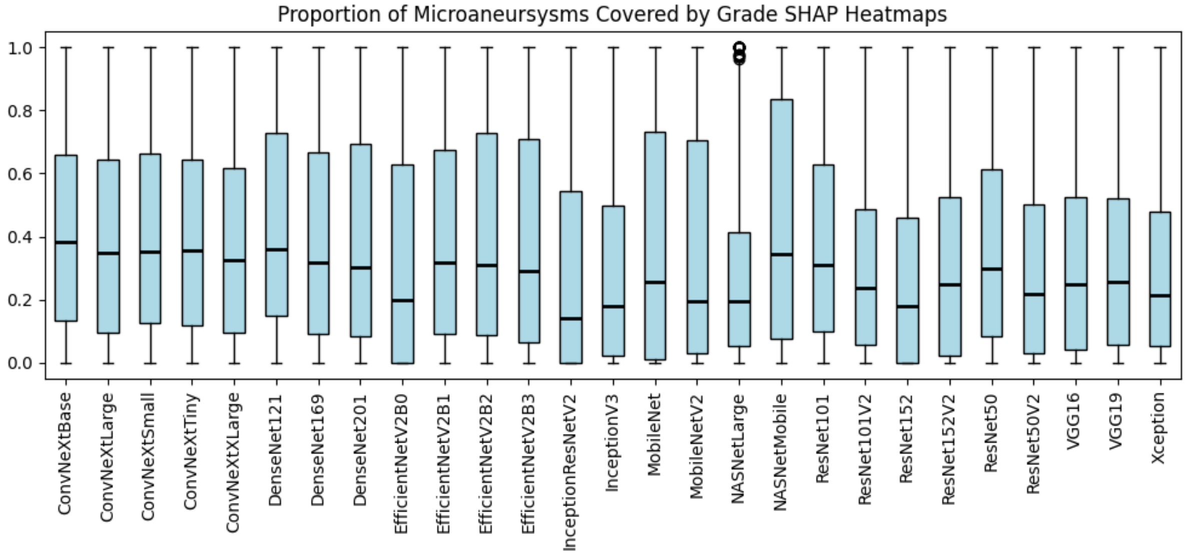

Figure 82: Proportion of microaneurysms covered by “grade” SHAP heatmaps.

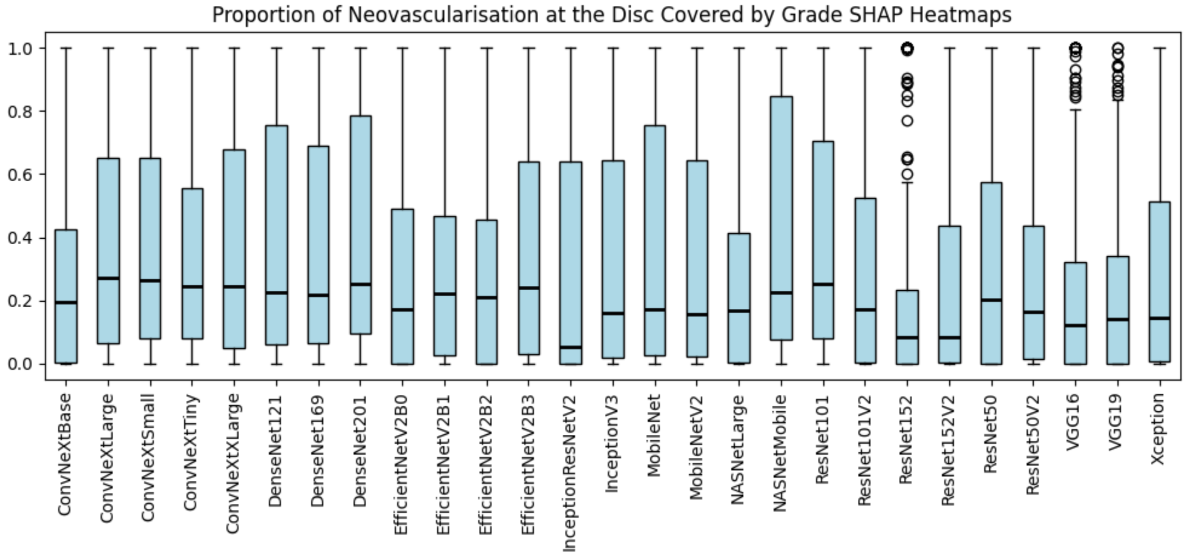

Figure 83: Proportion of neovascularisation at the disc covered by "grade" SHAP heatmaps.

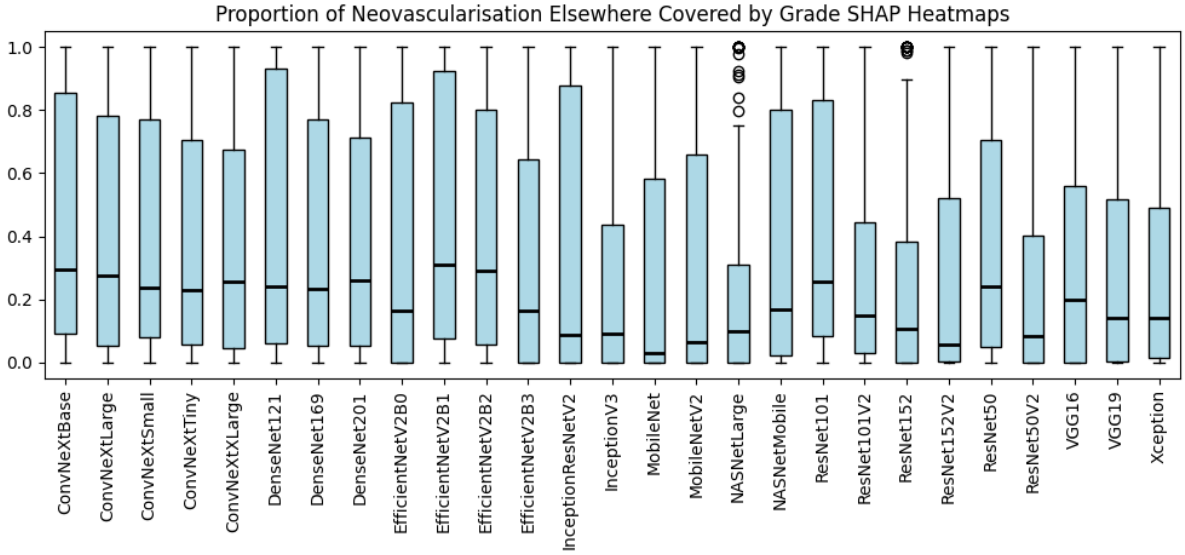

Figure 84: Proportion of neovascularisation elsewhere covered by "grade" SHAP heatmaps.

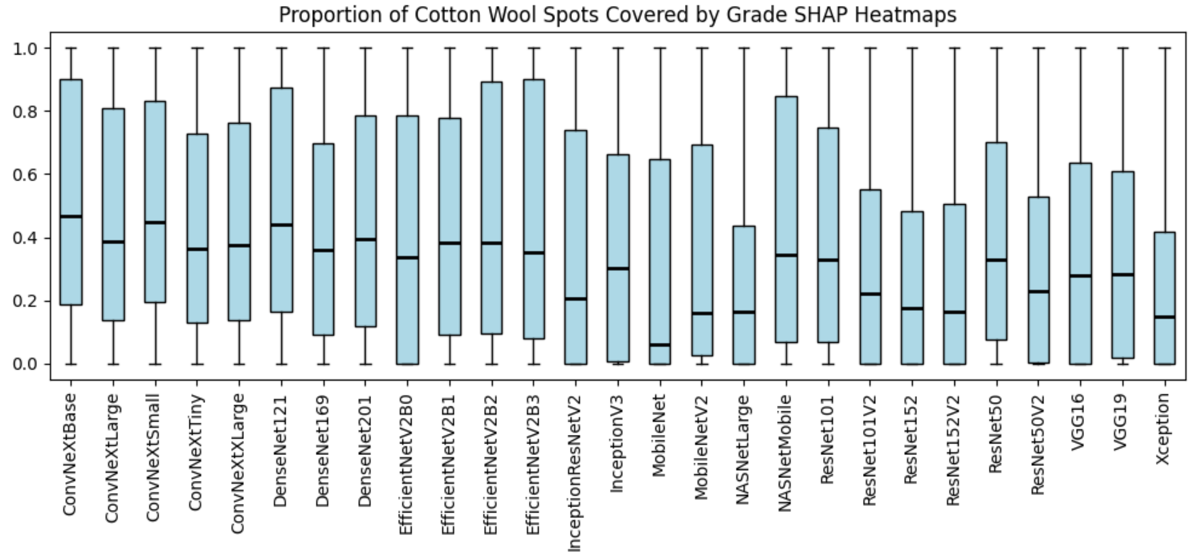

Figure 85: Proportion of cotton wool spots covered by "grade" SHAP heatmaps.

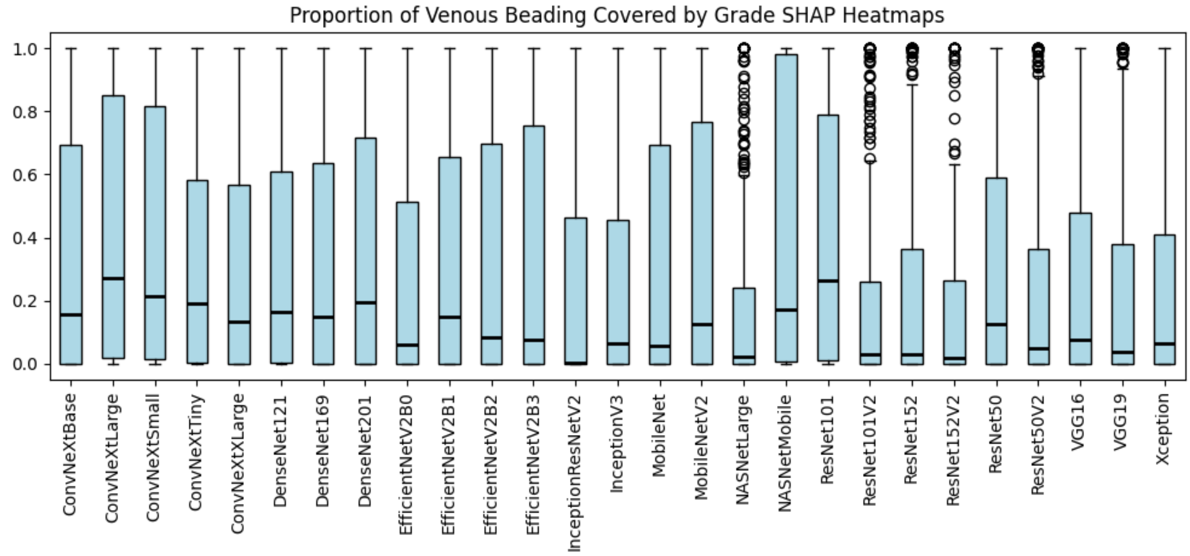

Figure 86: Proportion of venous beading covered by "grade" SHAP heatmaps.

#### 2.4 Heatmap variability

The following images represent the standard deviation of pixel values after bootstrapping with 10,000 iterations. Two heatmaps are presented: the standard deviation relative to the full dynamic range (e.g. SD / 255), and a normalised version to make the patterns easier to see.

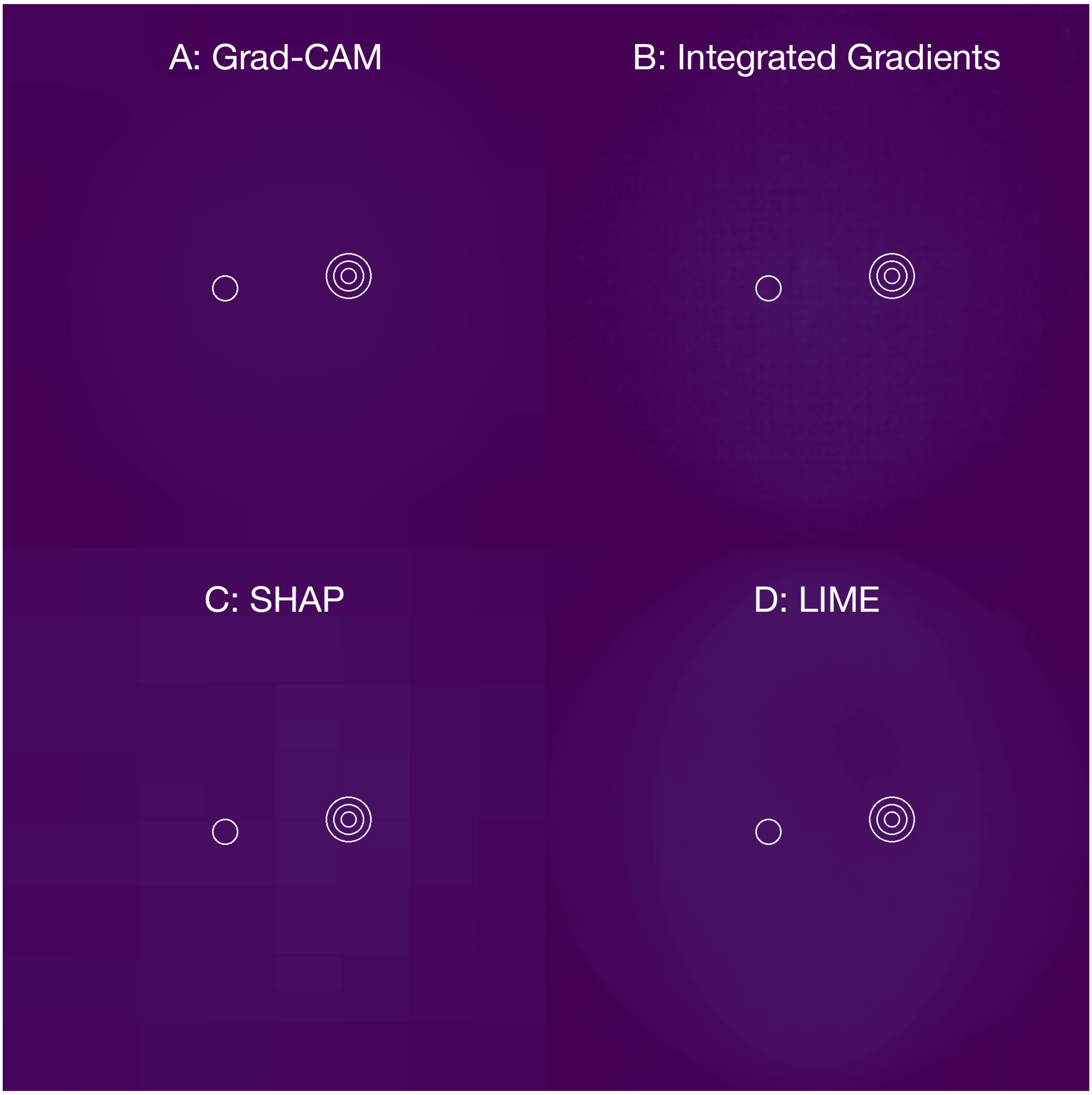

Figure 87: Standard deviation visualisation of “grade” model heatmaps after bootstrapping with 10,000 iterations for Grad-CAM (A), Integrated Gradients (B), SHAP (C) and LIME (D). Values are displayed relative to the maximum pixel intensity.

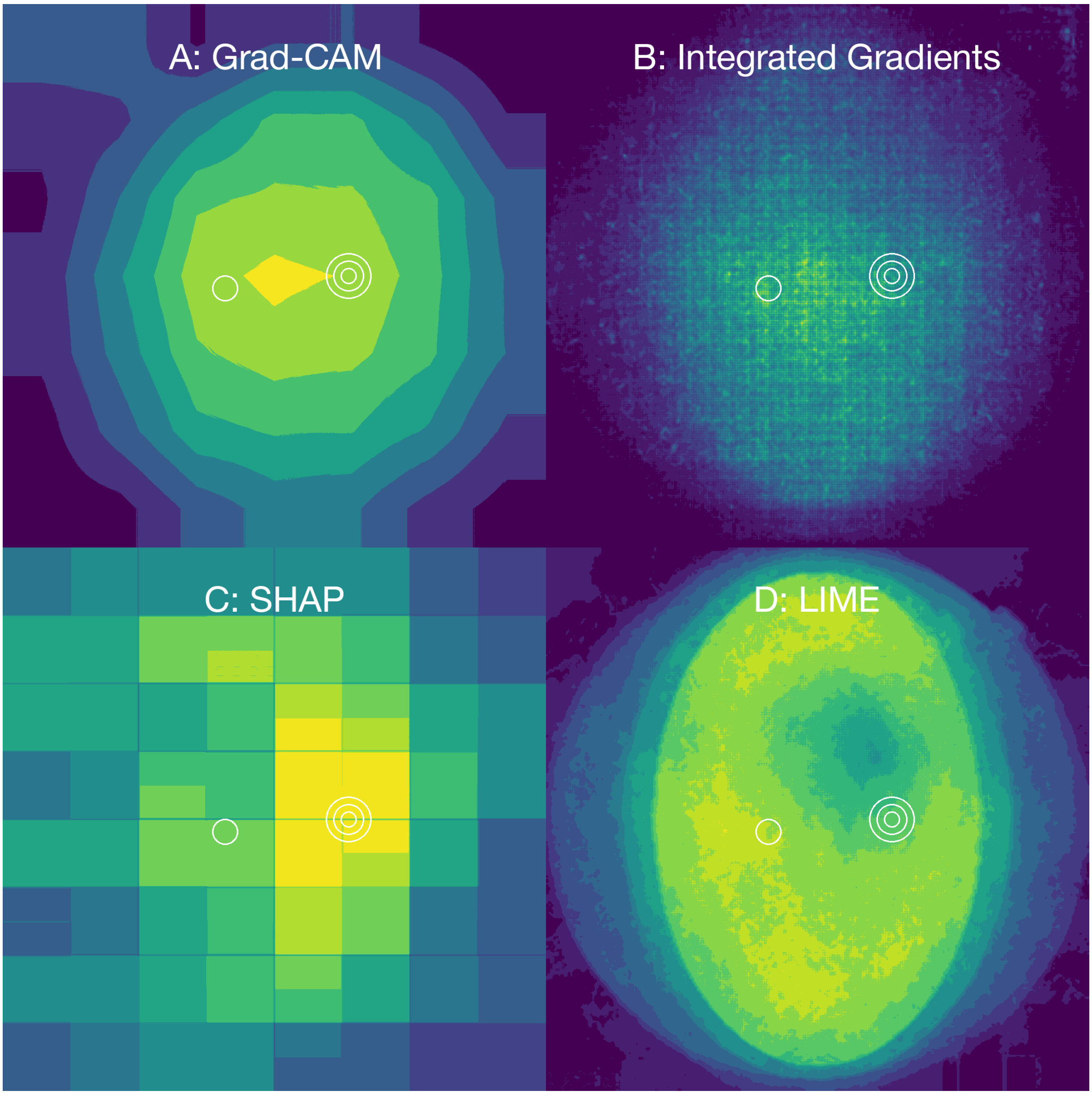

Figure 88: Standard deviation visualisation of “grade” model heatmaps after bootstrapping with 10,000 iterations for Grad-CAM (A), Integrated Gradients (B), SHAP (C) and LIME (D). Values have been normalised for clarity.
